# Trends and country-level variation in age at first sex in sub-Saharan Africa among birth cohorts entering adulthood between 1985 and 2020

**DOI:** 10.1101/2021.12.24.21267822

**Authors:** Van Kính Nguyen, Jeffrey W Eaton

## Abstract

**Background:** Debuting sexual intercourse marks exposure to pregnancy or fatherhood and sexually transmitted infections (STIs), including HIV. In sub-Saharan Africa (SSA), sexual debut varies according to cultural, religious, and economic factors, and encouraging delay has been a longstanding component of behavioural HIV prevention strategies. Age at first sex (AFS) is routinely collected in national household surveys, but data are affected by reporting biases, limiting utility to monitor trends and guide sexual health interventions.

**Methods:** We collated individual-level data from nationally-representative household surveys to analyse timing and national trends in AFS in 42 SSA countries. We used a log-skew-logistic distribution to characterize the time to AFS in a Bayesian spatio-temporal model, providing estimates of the sexual debut rate by sex, age, time, and country. We statistically adjusted for reporting biases by comparing AFS reported by the same birth cohorts in multiple survey rounds, allowing different reporting biases by sex and country.

**Results:** Median AFS in 2015 ranged from 15.8 among Angolan women to 25.3 among men in Niger. AFS was younger for women than men in 37/40 countries. The gap was largest for Sahel region countries and minimal in southern African countries. The distribution of female AFS was asymmetric with half debuting sex in an age range of 3.9 years [IQR 3.4–5.0 across countries]. Median AFS increased slightly between 1985 and 2020, ranging 0.84 years [IQR 0.11–1.55] and 0.79 [IQR -0.23–1.98] for females and males, respectively. The gender gap changed little over time in most countries. Female teens often reported higher AFS compared to when asked in their late twenties while male teens reported lower AFS; both sexes recalled a higher AFS in older ages compared to their thirties.

**Conclusions:** AFS increased slightly in most SSA countries, but changes were modest relative to large and persistent variation between countries and sexes, indicating relatively entrenched socio-cultural practices around sexual debut. Sexual health, family planning, and HIV/STI prevention services should adapt to local practices rather than focusing interventions to change AFS. These estimates for rates of sexual debut provide data to guide programmatic prioritization and implementation of sexual health services.

## Introduction

Debuting sexual intercourse is an important life course event for myriad developmental, social, and health reasons. Initiation of sexual activity marks the time when an individual becomes at risk for pregnancy or fatherhood and of acquiring and transmitting sexually transmitted infections (STIs) [1]. Younger age at first sex (AFS) has been associated with a higher rate of STIs [2–5] and increased risk of engaging risky sexual behaviours that facilitate STIs [6].

In sub-Saharan Africa (SSA), patterns and determinants of sexual debut have been of particular interest because of risk of exposure to HIV infection [7] and high rates of adolescent pregnancy [8], which is associated with increased risk of adverse maternal and child health outcomes. Age of first sex (AFS) varies between populations across SSA countries [9,10] according to cultural factors, religious practices, education, socio-economic status [11,12], and has changed over time along with these determinants. Secondary education and wealth quintile have been associated with later sexual debut among young women, though among men evidence is more mixed or in the opposite direction [13–15].

Delayed sexual debut has been associated with declines in HIV incidence [4,16], and encouraging young people to delay sexual debut was considered a priority for curtailing HIV epidemics in sub-Saharan Africa (SSA) during the 2000s as part of the “Abstinence, Be faithful, and use a Condom (ABC)” approach to HIV prevention [17]. Consequently, AFS in a population has become a key sexual risk indicator for determining and monitoring HIV and STI prevention activities [18,19].

Data on AFS are routinely collected in nationally-representative household surveys [20,21] conducted roughly every five years to monitor key health and development indicators. These data, reported retrospectively through face-to-face interviews, are susceptible to systematic response biases related to difficulty recalling dates of life events that happened long ago or desirable standards of society, culture, and health or political campaigns [22–25]. Studies in which the same individuals have been surveyed longitudinally have estimated that the percentage of inconsistent AFS reports ranged from 30% to 56% between survey rounds [26,27] and documented a tendency to report older AFS as respondents age [10,27,28]. Discrepant reports were in both directions and mostly a few years in magnitude but up to 10 years [21,27]. As the cohort aged from teenager to adulthood, men tended to report higher AFS while women reported lower AFS [10,29,30]. Including inconsistently reported individual observations, however, did not substantially affect estimates of the population median AFS [27,31] or predictors in multivariate modelling [21].

While many studies have documented evidence of reporting biases in AFS, few have attempted to systematically correct for biases to reconstruct how AFS has changed within and between populations in SSA. Moreover, analyses have tended to focus on the median AFS in a population [9,32], but other features of the distribution of AFS are less well characterised. The rate at which young people at each age initiate sexual activity determines level of exposure to adolescent pregnancy, the range of ages at which sexual health services need to be introduced, and sexual mixing dynamics that determine HIV and STI transmission and need for prevention.

Guided by previous literature on systematic differences in reported AFS as respondents aged [9,10,27], we previously developed and validated a survival analysis model to estimate the rate of sexual debut from population survey data, adjusting for reporting biases [33]. In this analysis, we extended and applied the model to incorporate all available AFS data in the SSA region. Through this analysis, we aimed to (1) characterise variation in AFS across countries and between sexes in SSA, (2) assess temporal changes in AFS over the last four decades, including during the period of intensive behavioural HIV interventions in eastern and southern Africa, and (3) provide data on the rate and full distribution of age at first sex as a resource for programmatic prioritisation and implementation of sexual health services and re-evaluation of strategies to identify the most at risk adolescent and young populations across SSA.

## Material and Methods

We used a skew logistic distribution to model the distribution of the log of reported age of first sex in a time to event modelling framework (Supplemental Text S1). In methodological comparisons, the log-skew-logistic distribution more accurately represented the sexual debut hazard than other common survival distributions [33]. To characterise temporal trends and geographic variation in the AFS distribution, we allowed the skew logistic distribution parameters to vary by country and birth cohort (Supplemental Text S1). Changes in the AFS over time for successive birth cohorts were represented by a second-order auto-regressive (AR2) process separately for males and females in each country. Since members of each birth cohort reported data about their AFS distribution in multiple household surveys spaced, typically, approximately five years apart, differences in the AFS reported among the same cohort in successive surveys quantified and permitted adjustment for systematic reporting bias in the AFS according to the age of the cohort at the time of the survey as described elsewhere [33]. The AFS reporting bias as function of age at report was smoothed with a first-order random walk (RW1) term for each sex and country. To accommodate countries with only one survey (making identification of country-specific bias from successive surveys impossible), the effect was estimated borrowing from other countries with a hierarchical model (Supplemental Text S1).

### Household survey data

Nationally representative surveys which collected AFS were collated, including Demographic and Health Surveys (DHS) [34], Multiple Indicators Cluster Surveys (MICS) [35], AIDS Indicators Survey (AIS) [34], Performance Monitoring and Accountability (PMA) [36], Population and Health Survey (PHS) [37], Population-based HIV Impact Assessment (PHIA) [38], Sexual Behavior Survey (SBS) [39], and South Africa HIV Prevalence, HIV Incidence, Behaviour and Communication Survey (SABSSM) [40]. Supplementary S1 Table lists the analysed surveys. We used only surveys in which individual records dataset were available for analysis. In these surveys, respondents were asked “How old were you (at which age) when you had sex (sexual intercourse/vaginal sex) for the very first time (if ever)?” for which valid responses were either never, integer age in years, or at first marriage/union which was replaced by the reported age at first marriage. Respondents’ age was calculated from the century-month-code records of data of birth and date of interview; if those variables were not presented, we calculated the century-month-code from the month and year of birth; if only the year was available, we set month of birth to June; otherwise, the age of respondent variable in the questionnaires were used.

### Model for age at first sex

AFS was reported as integer ages. We modelled the integer AFS as an interval censored event in which the exact event occurring between the reported AFS and AFS + 1. Individuals who reported not yet sexually debuting were right censored at their age at the interview. The likelihood for the reported AFS by each survey respondent was modelled using a time to event, interval censored model. A weighted pseudo-likelihood approached was used to account for unequal survey weights, which were normalized to the total sample size of each survey [41]. The three-parameter skew log-logistic distribution was used to describe the AFS distribution. This distribution permits a non-monotonic and asymmetric functional form of the sexual debut hazard by age which other commonly used distributions, e.g., log-normal, gamma, were not able to capture. It was also shown to have a better predictive value than the simpler two-parameter log-logistic distribution, which has a symmetric hazard function [42]. Illustrations of the distribution and its parameters are in Supplemental Text S1.

The model was fitted separately by sex but simultaneously for all countries, with independent shape and skewness parameters for each country. The linear predictor was modelled on the distribution’s scale parameter as an addictive effect of the spatial correlation, birth cohort, country-birth cohort interaction, age at report effect, and country-age interaction. Birth cohort and age at report effects were structured as second-order autoregressive (AR2) and first-order random walk (RW1) models, respectively; an intrinsic conditional auto-regressive (ICAR) model [43] was used to model the potential correlation between neighbouring countries. We assumed interactions that allowed the temporal trend and age effect to vary independently between countries. Model parameters were estimated with empirical Bayes using Template Model Builder [44]. Further technical specifications of the model constraints, priors, and implementations are in Supplemental Text S1. We computed posterior medians and 95% credible intervals of the outcomes of interests, including median AFS and proportion ever had sex by specified age thresholds, using posterior simulations of 3000 parameters samples.

## Results

Survey data were collated for 42 SSA countries. Seven SSA countries were not included in our analysis because they did not have AFS data (Cape Verde, Djibouti, Equatorial Guinea, Mauritania, Mauritius, Seychelles, and Somalia). Among 253 surveys considered (consisting of 251 female and 172 male datasets), 32 datasets had more than 10% missing the AFS variables (AFS and ever had sex indicator variable), mainly from PHIA and MICS surveys; 6 datasets had more than 10% responded as has ever had sex but did not answer a specific age (do not know or refused responses). These observations were removed from the analyses. In addition, 136 datasets had reports of AFS under seven years, ranging 0.01%– 1.67% of all observations. We considered those observations irregularities and removed from the analyses. We included only respondents aged 15–55 years at the time of interview and born after 1950. The final dataset contained 3,088,315 individual survey respondents for 56 birth cohorts from 1950 through 2005 (entering adulthood at age 15 between 1965 and 2020). The sample size per dataset ranged from 1364 (male, Eswatini MICS 2014) to 41,821 (female, Nigeria DHS 2018). Further details are in Supplemental Table S1.

### Spatial patterns of median AFS and proportion sexually active by age 18

For the birth cohort entering adulthood (turning age 15) in 2015, the median AFS ranged from 15.8 [IQR: 15.5–16.2] in Angola to 25.3 [IQR: 24.5–26.1] in Niger (Figure 1). Females had lower median AFS than males in most countries, but across countries the AFS between the two sexes were correlated such that countries where females had low median AFS also had males with low median AFS and vice versa. This was substantiated by a high correlation between male and female’s estimates of the skew-logistic distribution parameters (Supplemental Figure S1). The median AFS for men was less than women only in Lesotho, South Africa, and Comoros. Only females in Rwanda, Burundi, Comoros, and Eritrea had median AFS above twenty.

**Figure 1.**
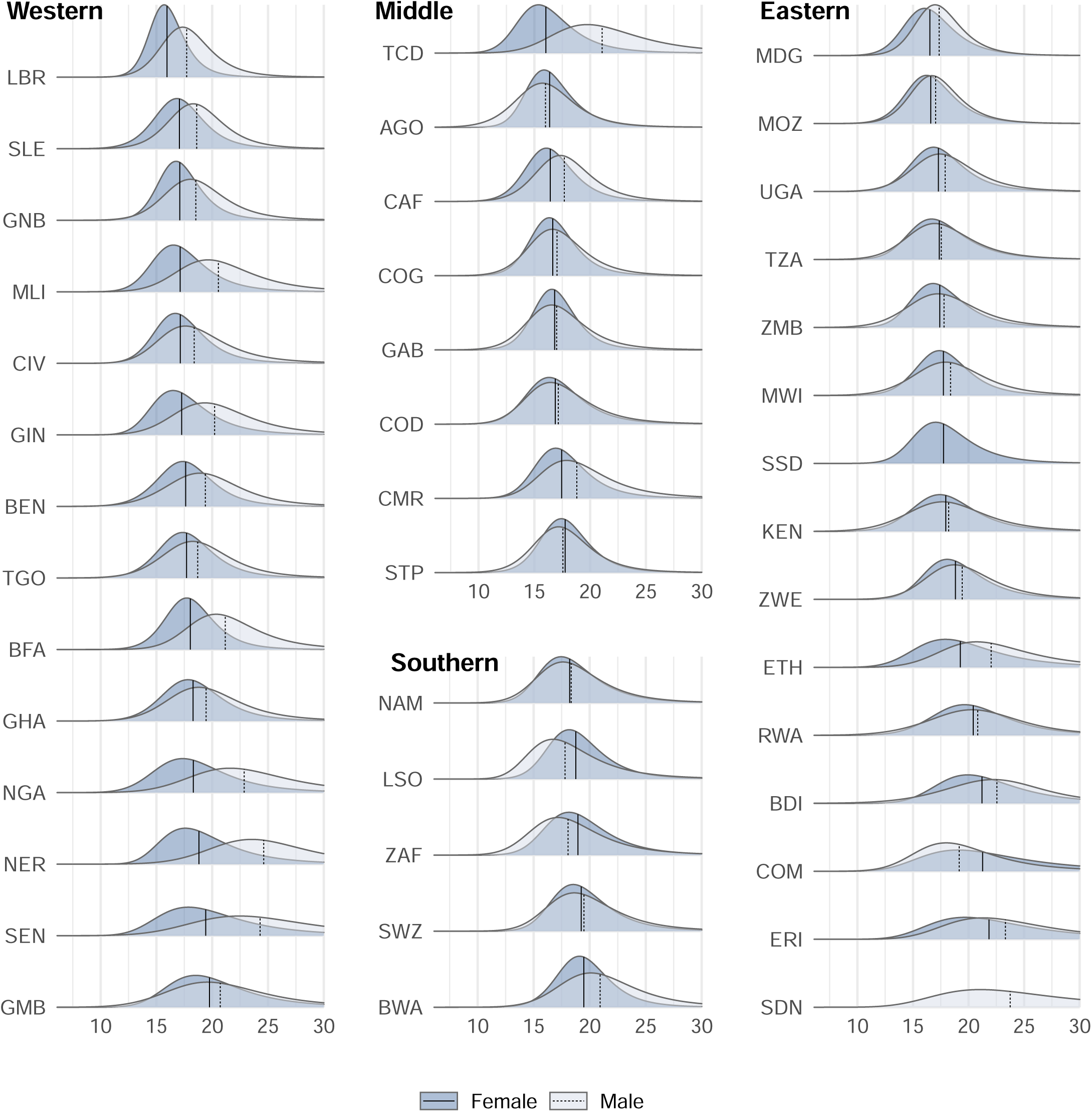
Distribution of the age at first sex by region, country, and sex. The median of each distribution is shown with the vertical lines. Estimates are for the birth cohort turning fifteen in 2015 and assumed age 23 years at survey report. Within each region, countries sequenced by increase median female AFS. Numerical values and 95% uncertainty intervals for estimates are reported in Supplemental Table S3. Country names and region allocations corresponding to the three-letter are ISO 3166-1 alpha-3 code labels were as follows: ***Western* Africa:** BEN – Benin; BFA – Burkina Faso; CIV – Cote d’Ivoire; GHA – Ghana; GIN *– Guinea; GMB – Gambia; GNB – Guinea-Bissau; LBR – Liberia; MLI – Mali; NER – Niger; NGA – Nigeria; SEN – Senegal; SLE – Sierra Leone; TGO – Togo;* ***Middle* Africa:** AGO - Angola; CAF - Cent. Afr. Republic; CMR - Cameroon; COD - Dem. Rep. Congo; COG - Congo; GAB - Gabon; STP - Sao Tome and Principe; TCD - Chad; ***Eastern* Africa:** BDI - Burundi; COM - Comoros; ERI - Eritrea; ETH - Ethiopia; KEN - Kenya; MDG - Madagascar; MOZ - Mozambique; MWI - Malawi; RWA - Rwanda; SDN - Sudan; SSD - South Sudan; TZA - Tanzania; UGA - Uganda; ZMB - Zambia; ZWE - Zimbabwe; ***Southern* Africa:** BWA - Botswana; LSO - Lesotho; NAM - Namibia; SWZ - Eswatini; ZAF - South Africa *(also in* Supplemental Table S4).

The spatial variation in sexual debut is further evident in the percentage who ever had sex before age 18 years (Figure 2). A very low percentage of men in the Sahel region of Africa had debuted sex by age 18, with the lowest 4.4% [3.2–6.0%] percent in Niger. Among women, the percentage sexually active by age 18 was lowest, below 45%, in the southern countries, and as low as 18.4% [16.5–20.6%] in Burundi. In both sexes, Botswana, Burundi, and Rwanda all had older AFS than the surrounding region whereas Mozambique had notably younger AFS in the southeast Africa. At the other extreme, women in Liberia and men in Angola were most likely to debut sex under eighteen with 86.1% [82.8–88.9%] and 77.5% [73.2–80.8%], respectively. AFS were spatially homogenous in the remained locations. Similar patterns were observed for the percentage ever had sex under fifteen (Supplemental Figure S2) where the largest estimate was 37.7% [32.9–42.7%] in Angola male following by Chad female at 33.3% [31.5–35.1]. In nearly half of the cases (37/82) less than 10% had initiated sexual activity by age 15. (Supplemental Figure S2, Supplemental Table S2).

**Figure 2.**
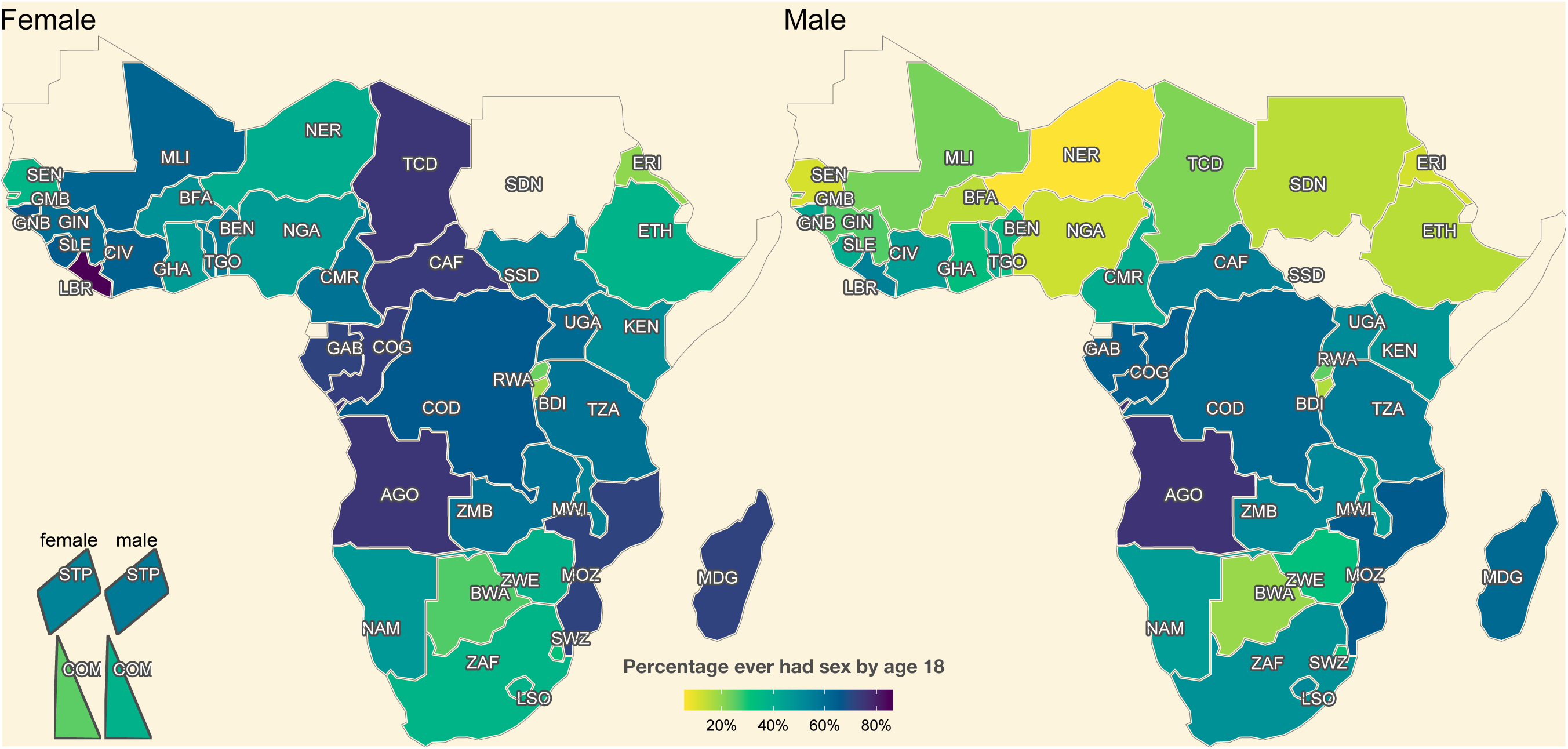
Percentage ever had sex by age 18 among the birth cohort turning 15 in the year 2015. The island countries Comoros and São Tomé & Príncipe are represented in the bottom left. Countries without data are coloured the same as the background. Numerical estimates and 95% uncertainty interval are in Supplemental Table S2. Country names corresponding to the three-letter ISO 3166-1 alpha-3 code labels are in Figure 1 caption and Supplemental Table S4.

### Time trends in AFS

The median AFS increased between birth cohorts entering adulthood in 1985 and 2020 in 60 of 82 time-series estimated by country and sex (Figure 3), and in 40 increased more than one year. During this period, the average change in median AFS across all countries was 0.84 years [IQR: 0.11-1.55] for females and 0.79 [IQR: -0.23 -1.97] for males. The changes, however, have not been monotonic but fluctuated over time (such as in Gambia, Guinea Bissau, Uganda, and Rwanda) and varied between countries. The estimated trends were similar for male and female in most countries with few exceptions of Comoros, South Africa, and Lesotho in the most recent birth cohorts. Increases in AFS were especially large for men in some West African countries (e.g., Senegal, Niger, Mali, Chad, Nigeria, Burkina Faso, Benin). The largest estimated decreases were also among men (e.g., Tanzania, Sierra Leon, Lesotho, Mozambique). Eastern and Southern Africa exhibited minimal changes in AFS during the time frame with the average increase of 0.31 and 0.12 years, respectively.

**Figure 3.**
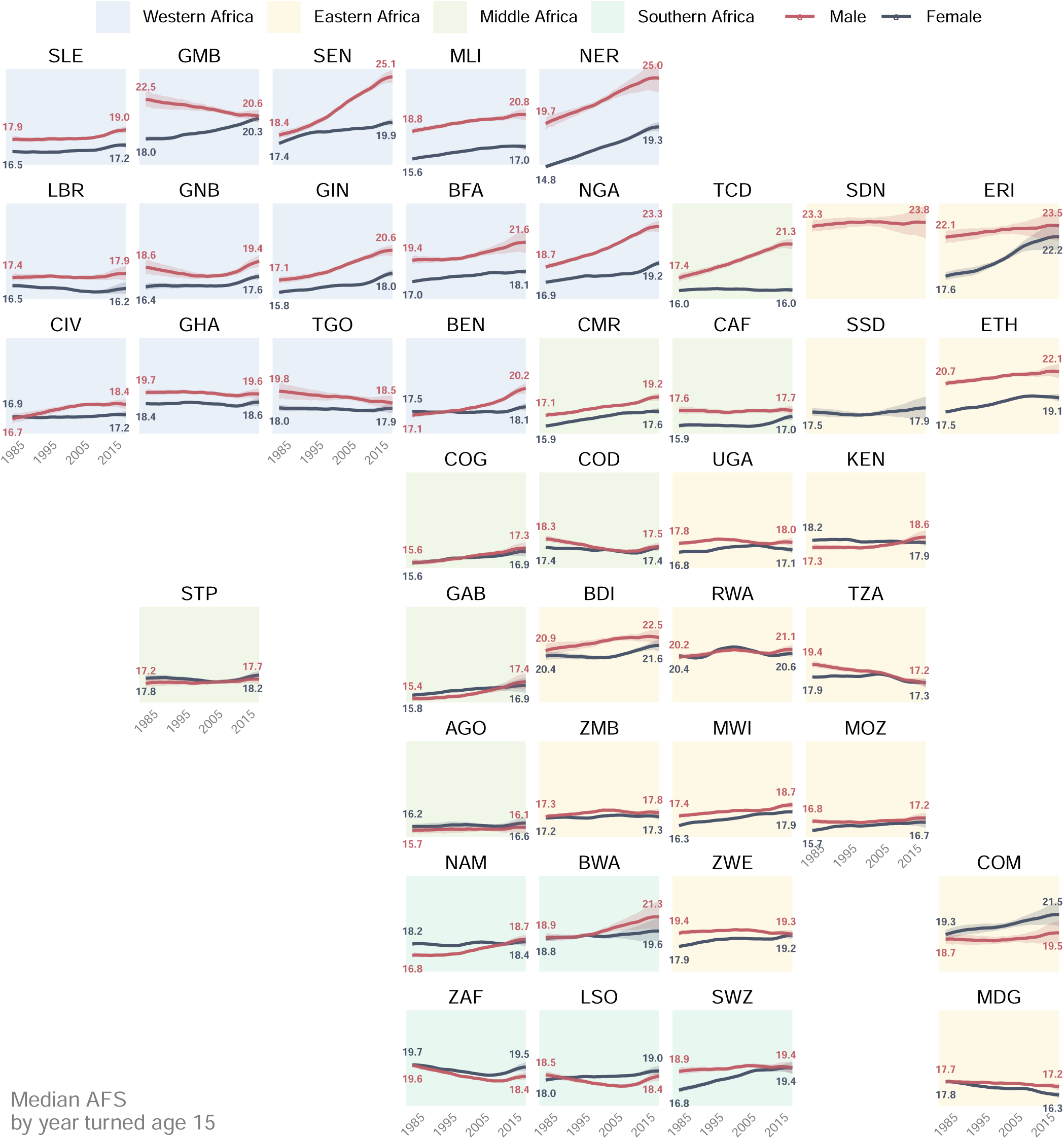
Trend in median age at sexual debut for men and women in Sub Saharan-Africa countries by year turned age 15 (x-axis). For countries where the most recent survey is before 2015, trends are extrapolated for recent birth cohorts based on the overall trend across the region and its neighbours. Countries without AFS data are excluded from this figure. Country names corresponding to the three-letter ISO 3166-1 alpha-3 code labels are in Figure 1 caption and Supplemental Table S4.

In the same birth cohort, the gap in median AFS between women and men was under two years in most countries and remained unchanged in the region. The gap was larger in the West Africa countries, especially those in the Sahel and Nigeria, and in the Horn of Africa (Ethiopia, Eritrea). Median AFS increased most in Western Africa (Figure 4). Middle Africa had the lowest median AFS on average for both men and women. In Southern Africa, which is dominated by the large population of South Africa, there was smaller sex difference in median AFS and median AFS decreased since the 1985 adult cohort, in contrast to the increasing trend in other regions.

**Figure 4.**
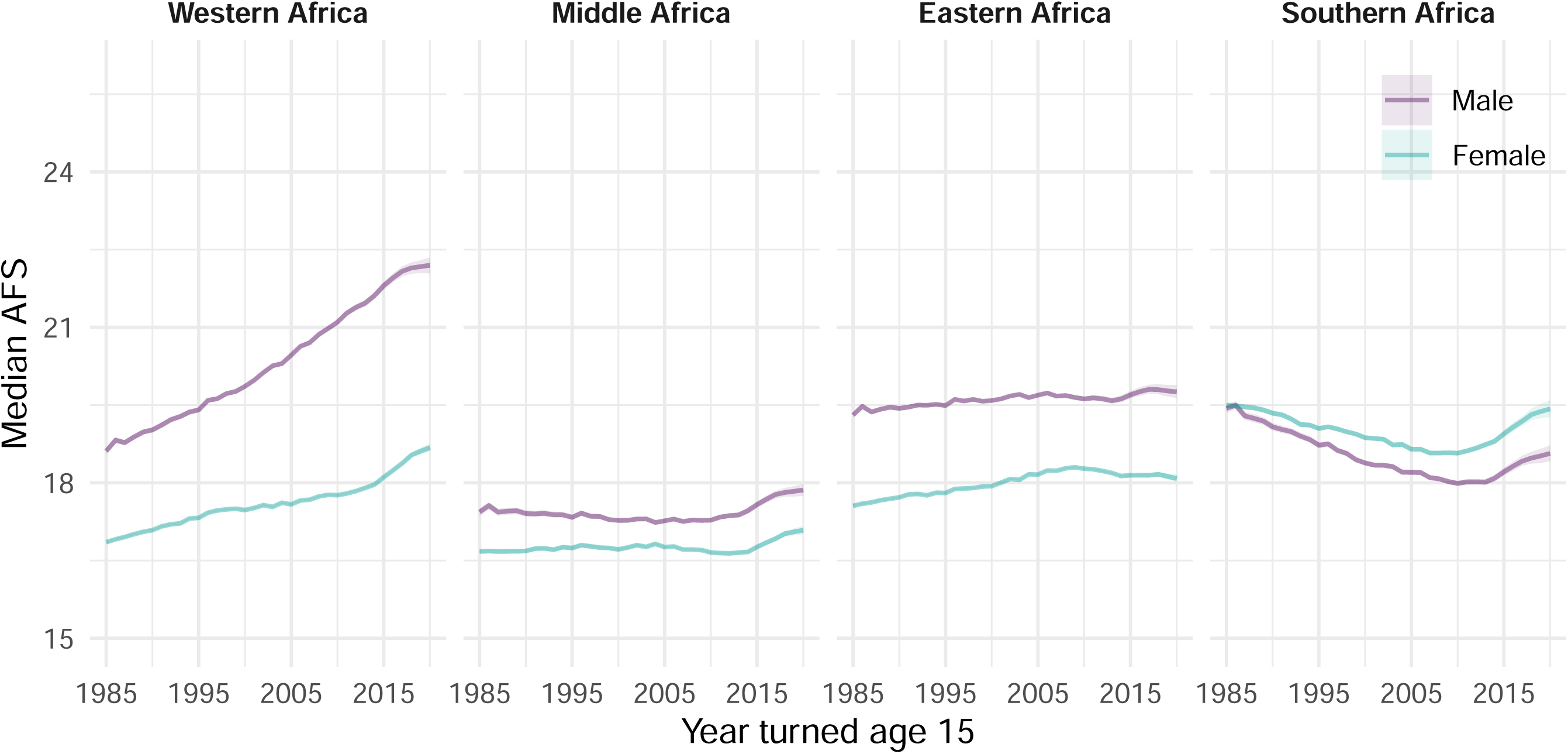
Aggregated trend of median age at first sex by region for men and women. Sudan was included in Eastern Africa. The regional trend is a weighted average of the countries included in the analyses by their respective population size in 2020 sourced from the UN World Population Prospects 2019. Country classifications in each region are defined in the Figure 1 *caption and Supplemental Table S4*.

### Distribution of age at sexual debut by sex

The distribution of AFS was asymmetric in most countries with various degree of skewness (Figure 1). The inter-quartile range (IQR) of the AFS distribution represent the age range with the highest sexual debut activity and its magnitude reflects how fast the rate of most of the population becomes sexually active. In most countries the IQR was under five years with an average of 4.0 [2.8–8.2] years. The duration for 50% of a birth cohort to initiate sexual activity was as short as 2.2 years (Liberia - female) or up to 9.6 years (Sudan - male).

Between sexes, in most countries (37/40, excluding two countries with only one sex data) the IQR was shorter for females than for males; among these, 18 countries had a difference of more than a year. The distribution of AFS spanned a longer range for countries of the Sahel and Horn of Africa.

### Biases in reported AFS by age at interview

The estimated pattern for the effect of age at the time of interview on the reported AFS for each country are shown in Figure 5. Data in different countries yielded similar information on the effect of age at report bias. Among females, there was a tendency to report higher AFS in their teens compared to when they reached their late twenties, indicating under-reporting of sexual activity among the youngest survey respondents. The opposite pattern was observed among men—young men tended to report a younger AFS distribution. For both sexes, the reported distribution of AFS increased as the cohort aged. The direction of both bias’s directions varied across countries. The coefficient estimates of the biases, however, only show the relative difference between ages, but do not identify at which age people would be both truthful and accurately recall their AFS. To arrive at a marginal estimate for the AFS of each birth cohort, we chose age 23 as the reference age in both female and male and report estimates using this reference age throughout the results presented.

**Figure 5.**
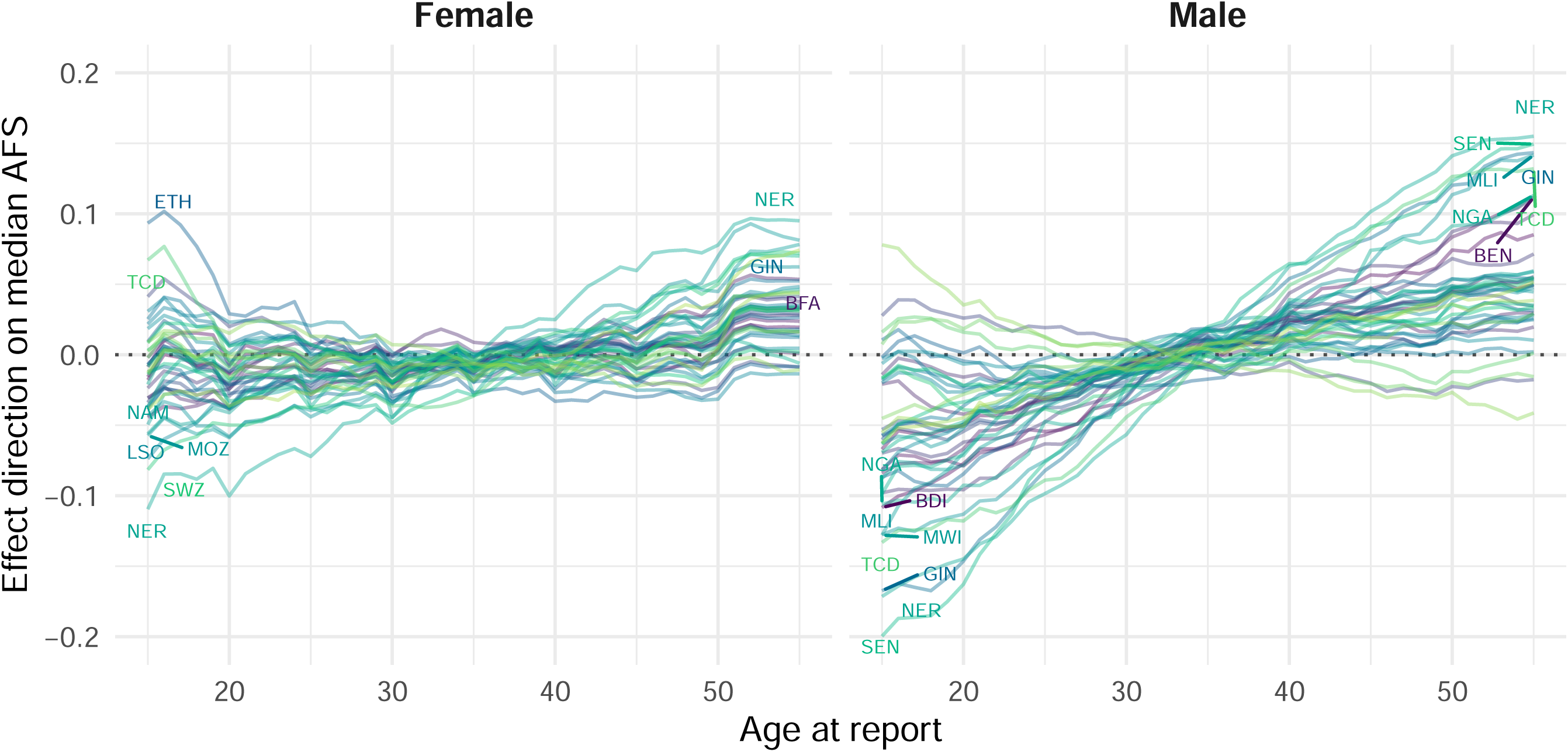
Direction of the effect of age at report bias on the median age at first sex estimate relative to other ages. A positive coefficient in the figure corresponds to an increase in median AFS estimate reported by the respective age compared to a median AFS reported by the age with that coefficient equals zero.

## Discussion

In this study, we systematically analysed trends and patterns in debuting sexual activity across sub-Saharan Africa among cohorts entering adulthood from 1985 to 2020, a period during which the continent combatted the HIV/AIDS pandemic and experienced rapid social change, urbanisation, and economic development. Assessment of past sexual debut trends and current status in each country are important to help evaluate the impact of the past interventions, respond to the increasing sexual health needs for young people, and identify the key ages at which to provide sexual health services. We applied a statistical model to account for reporting biases that have previously hampered consistent interpretation of AFS recorded in household surveys and used a parsimonious but flexible model to characterise the distribution of AFS over ages.

The gap between AFS for men and women differed greatly between the southern and northern countries with little evidence of changes over time in most countries. The gap in AFS remained large and widened over time in the north while remained small and narrowed over time in the south. In the Western Africa region, the gap of more than two years in median AFS between male and female in many countries implies more disparity in sexual partner mixing compared to countries in the south. Large gender gaps could make adolescent girls more vulnerable to adverse sexual health outcomes [45].

The median AFS has increased in many parts of sub-Saharan Africa over the past three decades, especially in the northern-most countries, but the timing and magnitude of changes varied. In countries heavily affected by HIV/AIDS epidemic in southern and eastern Africa, in which there has been a multitude of intervention programmes focused on reducing sexual risk, median AFS only slightly increased or even decreased. Other analyses have reported that trends towards earlier sexual debut are not pronounced [9] and median AFS in SSA increased over time [32]. Previous findings from individual countries also found similar results such as a small increase in men’s AFS in Zambia [28] or both sexes in Uganda [46]. The only modest or decreasing AFS may indicate that efforts to encourage delayed sexual initiation via expanding access to education [47] may not have had large effect on AFS, though access to education may have improved knowledge and accessibility to safe sex practices and sexual health services. In contrast to the Southern African countries, AFS has increased more steeply in Western Africa where the HIV/AIDS epidemic is not as severe [48]. Stricter cultural and religious norms about abstaining sex until marriage in this region may have meant that gains in female education [49] leading to later marriage in for women [50] resulted more directly in delayed sexual debut, while increasing economic uncertainty has pressured men to delay marriage [51]. Taken together, health policies aimed at effecting changes have had limited or inconsistent effects. Strategies adapt sexual health, family planning, and HIV/STI prevention promotion to changes in AFS to appropriately target the timing of the current and upcoming adult birth cohorts entering sexually active life are likely to be more effective.

Reproductive health and HIV monitoring indicators have been focused on single measure including median AFS in a population or the proportion initiating sexual activity by specified ages, resulting a loss of age-specific sexual debut behaviour. Our analysis calculated the full distribution of debuting sexual activity, reflecting the range of ages at which adolescents and young adults may need to become engaged in sexual health, family planning, and HIV/STI prevention services. The rate at which populations become sexually active varied across countries, but typically was slightly more peaked for young women. For young women, the percentage sexually experienced increased from 25% to 75% in around four years compared to five years for men. This varied across countries, however. In some, sexual debut of a population occurred in a very short span of age while in others in a gradual manner (Figure 1). Two populations with the same median AFS, but different shapes parameters may have different demand for reproductive health and STIs services where one may need to cover a larger target age than the other. Programming prevention services efforts should be directed accordingly to deliver services to the crucial period of a sexually active life using our detailed estimate of the AFS distribution.

Previous research using longitudinal data or sexual activity biomarkers has identified reporting biases in AFS that can obfuscate trends assessed from cross-sectional surveys [27,32,45,52], yet it was difficult to address them in isolated analyses of specific surveys or single countries considering wide varieties of pattern and magnitude in reporting biases. This study addressed this problem by comparing reports among the same cohort in successive surveys and learning from all the available data in SSA to extract the pattern of the bias. Biases existed in most countries and we identified a few general patterns in the age at report bias. Young men tended to report earlier AFS than when asked again in later surveys, while young women tended to report older AFS. Above age 30, the reported AFS tended to increase as the cohort aged, especially for men, indicative of recall biases for exact age for events that occurred long ago. These results are consistent with previous findings documenting biased self-reporting of sexual activity by norms that boys are expected to engage in sexual activity and girls are expected to abstain until marriage [24][10,29,30]. Other country-specific analyses have also documented that respondents were less likely to report a young AFS in later surveys [28]. While adjusting for these biases to reconstruct consistent time trends is a strength of our model, the variation in magnitude and direction of biases across countries implies caution about confidently distinguishing actual trends from changing reporting biases among the youngest cohorts in new surveys. This will be especially challenging in settings with fewer surveys from which to triangulate across different time points in life of a birth cohort [24]. The results from our study still highlight that age- and sex-specific reporting bias is prevalent and could lead to misjudgement of the needs for sexual health services in adolescent and young population – especially for young girls where more might be needed than the data suggest.

This study has several limitations. First, while we were able to adjust for relative reporting differences by age at report, there was no ‘gold standard’ measure to identify at which age reporting was most accurate. The chosen reference age of twenty-three is at most ten years difference from the AFS and above the average age of marriage of women in SSA, thus it is expected that reports at this age are less influenced by social desirability compared to a younger age and recall more accurate compared to older reporting age. In addition, as digit preference for age reporting ending in zero or five is a common issue [53], using ages around 25 or 30 might introduce biases to the reported estimates. Second, country- and sex-specific differences of potential confounders to median AFS such as religion [54] were not modelled. Third, our model allowed the median AFS to change over time, and for the shape of the distribution and reporting bias to vary by country but did not allow the shape or reporting bias to change over time within each country. Fourth, survey series varied slightly in the eligibility of respondents asked about AFS, the way AFS was recorded, and data processing procedures. For example, respondents older than 24 were not asked AFS in some in MICS or FHS. In DHS it is standard practice to recode internally inconsistent data such as reported AFS that is older than the age at the time of survey, after the conception of the first child, or after the first marriage (45), but this is not done in all surveys, as evidenced in larger proportions of excluded observations (S1 Table). Fifth, we accounted for survey design by incorporating survey weights into the model pseudo likelihood [41], but did not model stratified clustered sampling designs. Finally, results in this paper are limited to SSA at national level. Similar methods to ours could be applied for subnational estimates, which may provide more granular and actionable characterisation of sexual risk dynamics. Subnational analyses should consider urban-rural migration trajectories and implications of respondents no longer residing in the location where retrospectively reported sexual debut occurred.

## Conclusions

AFS has increased slightly in most, but not all, SSA countries. However, in most cases changes over-time within a country were small compared to large and relatively persistent variation between countries and genders, reflecting deeply rooted cultural norms around sexual debut. AFS trend remained relatively stagnant in the countries most affected by the HIV epidemic suggests behavioural intervention programs encouraging delayed sexual debut [55] have had relatively modest effects on AFS. Instead, adapting sexual health, sex education, family planning, and STI and HIV prevention services to local norms around sexual debut should be prioritised over seeking to intervene to change AFS. Our estimates of age- and sex-specific sexual debut rates provides data to support locally-adapted health programming and more detailed inputs to epidemic models of HIV and STIs that may also help to shed light on national and regional variation in epidemic burdens and transmission dynamics, for example the role of the larger age gaps in west and middle African countries compared eastern and southern regions.

## Data Availability

All data produced are available online at dhsprogram.com/data/Dataset-Types.cfm and https://mics.unicef.org/surveys and https://phia.icap.columbia.edu

## Declarations

### Ethics approval and consent to participate

The study involved secondary analysis of anonymised publicly available data following project approval from the Demographic and Health Surveys Program, the Performance Monitoring for Action project, the Population-based HIV Impact Assessment project, and South African National HIV Prevalence, Incidence, Behaviour and Communication Survey. Primary survey protocols were reviewed and approved by the relevant national ethics committee in each country.

### Consent for publication

Not applicable

### Availability of data and materials

The datasets analysed during the current study are available in the Demographic and Health Surveys Program (https://dhsprogram.com/Data/), the Performance Monitoring for Action project (https://www.pmadata.org/data), the Population-based HIV Impact Assessment project (https://phia-data.icap.columbia.edu), and South African National HIV Prevalence, Incidence, Behaviour and Communication Survey (http://www.hsrc.ac.za/).

### Competing interests

The Authors declare that there is no conflict of interest.

### Funding

This research was supported the Bill and Melinda Gates Foundation (OPP1190661), National Institute of Allergy and Infectious Disease of the National Institutes of Health under award numbers R01AI136664 and R01AI152721, and the MRC Centre for Global Infectious Disease Analysis (reference MR/R015600/1), jointly funded by the UK Medical Research Council (MRC) and the UK Foreign, Commonwealth & Development Office (FCDO), under the MRC/FCDO Concordat agreement and is also part of the EDCTP2 programme supported by the European Union. The funders have no roles in the study design or manuscript preparation.

### Authors’ contributions

VKN and JWE conceived the work. VKN and JWE designed the work. VKN implemented the models. Both authors critically reviewed model results throughout the model development process. VKN wrote the first draft of the manuscript. Both authors critically edited the manuscript for intellectual content.

## Acknowledgements

This research was supported the Bill and Melinda Gates Foundation (OPP1190661). We acknowledge joint MRC Centre funding from the UK Medical Research Council and Department for International Development via MRC MR/R015600/1.

## Supplemental Materials

Text S1. Skew log-logistic distribution & specifications of the survival model

Figure S1. Parameter distribution correlation

Figure S2. Spatial distribution of the percentage ever had sex under fifteen.

Figure S3. Proportion ever had sex and fitted value by country and sex

Table S1. List of surveys used in the analyses

Table S2. Parameter estimates by country and sex

Table S3. Proportion ever had sex under 15 &18 by country and sex

## Notes

### Competing Interest Statement

The authors have declared no competing interest.

### Author Declarations

This study involves only openly available human data, which can be obtained from: dhsprogram.com/data/Dataset-Types.cfm and https://mics.unicef.org/surveys and https://phia.icap.columbia.edu

### Summary of Updates

We revised to add details on research context, contributions, and aims of the research. In addition, we add 7 new surveys which have become available since the last submission.

## References

[1] WHO. Report on global sexually transmitted infection surveillance 2018. 2018.

[2] Pettifor AE, van der Straten A, Dunbar MS, Shiboski SC, Padian NS. Early age of first sex. AIDS 2004. https://doi.org/10.1097/01.aids.0000131338.61042.b8.

[3] Kaestle CE, Halpern CT, Miller WC, Ford CA. Young age at first sexual intercourse and sexually transmitted infections in adolescents and young adults. Am J Epidemiol 2005;161:774–80. https://doi.org/10.1093/aje/kwi095.

[4] Hallett TB, Gregson S, Lewis JJC, Lopman BA, Garnett GP. Behaviour change in generalised HIV epidemics: Impact of reducing cross-generational sex and delaying age at sexual debut. Sex Transm Infect 2007;83:i50–54. https://doi.org/10.1136/sti.2006.023606.

[5] Wand H, Ramjee G. The relationship between age of coital debut and HIV seroprevalence among women in Durban, South Africa: a cohort study. BMJ Open 2012;2. https://doi.org/10.1136/bmjopen-2011-000285.

[6] Kaplan DL, Jones EJ, Olson EC, Yunzal-Butler CB. Early Age of First Sex and Health Risk in an Urban Adolescent Population. J Sch Health 2013. https://doi.org/10.1111/josh.12038.

[7] Howard AL, Pals S, Walker B, Benevides R, Massetti GM, Oluoch RP, et al. Forced Sexual Initiation and Early Sexual Debut and Associated Risk Factors and Health Problems Among Adolescent Girls and Young Women — Violence Against Children and Youth Surveys, Nine PEPFAR Countries, 2007–2018. Morb Mortal Wkly Rep 2021;70:1629–34. https://doi.org/10.15585/mmwr.mm7047a2.

[8] Mbabazi C, Kintu A, Asiimwe JB, Ssekamatte JS, Shah I, Canning D. Proximate and distal factors associated with the stall in the decline of adolescent pregnancy in Uganda. BMC Public Health 2021;21:1875. https://doi.org/10.1186/s12889-021-11403-6.

[9] Wellings K, Collumbien M, Slaymaker E, Singh S, Hodges Z, Patel D, et al. Sexual behaviour in context: a global perspective. Lancet Lond Engl 2006;368:1706–28. https://doi.org/10.1016/s0140-6736(06)69479-8.

[10] Zaba B, Pisani E, Slaymaker E, Boerma JT. Age at first sex: Understanding recent trends in African demographic surveys. Sex Transm Infect 2004;80:28–35. https://doi.org/10.1136/sti.2004.012674.

[11] Upchurch DM, Levy-Storms L, Sucoff CA, Aneshensel CS. Gender and ethnic differences in the timing of first sexual intercourse. Fam Plann Perspect 1998;30:121– 7.

[12] Zaba B, Pisani E, Slaymaker E, Boerma JT. Age at first sex: Understanding recent trends in African demographic surveys. Sex Transm Infect 2004;80:28–35. https://doi.org/10.1136/sti.2004.012674.

[13] Gupta N, Mahy M. Sexual initiation among adolescent girls and boys: trends and differentials in sub-Saharan Africa. Arch Sex Behav 2003;32:41–53. https://doi.org/10.1023/a:1021841312539.

[14] McGrath N, Nyirenda M, Hosegood V, Newell M-L. Age at first sex in rural South Africa. Sex Transm Infect 2009;85:i49–55. https://doi.org/10.1136/sti.2008.033324.

[15] Madise N, Zulu E, Ciera J. Is poverty a driver for risky sexual behaviour? Evidence from national surveys of adolescents in four African countries. Afr J Reprod Health 2007;11:83–98.

[16] Gregson S, Garnett GP, Nyamukapa CA, Hallett TB, Lewis JJC, Mason PR, et al. HIV decline associated with behavior change in eastern Zimbabwe. Science 2006;311:664– 6. https://doi.org/10.1126/science.1121054.

[17] Halperin DT, Steiner MJ, Cassell MM, Green EC, Hearst N, Kirby D, et al. The time has come for common ground on preventing sexual transmission of HIV. The Lancet 2004;364:1913–5. https://doi.org/10.1016/S0140-6736(04)17487-4.

[18] UNAIDS. Monitoring the Declaration of Commitment on HIV/AIDS: guidelines on construction of core indicators n.d.

[19] Dehne, K. L., Gabriele, Riedner. Sexually transmitted infections among adolescents : the need for adequate health services. n.d.

[20] Aisha NZD, Philipp U, Vladimíra K. Sexual Activity by Marital Status and Age: A Comparative Perspective, Technical Paper No. 11 2017.

[21] Assche SB Van. Are we measuring what we want to measure? Individual consistency in survey response in rural Malawi. Demogr Res 2003;9:77–108. https://doi.org/10.4054/DemRes.2003.S1.3.

[22] Catania JA. A framework for conceptualizing reporting bias and its antecedents in interviews assessing human sexuality. J Sex Res 1999;36:25–38. https://doi.org/10.1080/00224499909551964.

[23] Wellings K, Collumbien M, Slaymaker E, Singh S, Hodges Z, Patel D, et al. Sexual behaviour in context: a global perspective. Lancet Lond Engl 2006;368:1706–28. https://doi.org/10.1016/S0140-6736(06)69479-8.

[24] Gersovitz M. HIV, ABC and DHS: age at first sex in Uganda. Sex Transm Infect 2007;83:165–8. https://doi.org/10.1136/sti.2006.021576.

[25] United Nations. World population monitoring: adolescents and youth. U N 2012:1.

[26] Eggleston E, Leitch J, Jackson J. Consistency of Self-Reports of Sexual Activity among Young Adolescents in Jamaica. Int Fam Plan Perspect 2000;26:79–83. https://doi.org/10.2307/2648271.

[27] Wringe A, Cremin I, Todd J, McGrath N, Kasamba I, Herbst K, et al. Comparative assessment of the quality of age-atevent reporting in three HIV cohort studies in sub-Saharan Africa. Sex Transm Infect 2009;85. https://doi.org/10.1136/sti.2008.033423.

[28] Slaymaker E. Monitoring trends in sexual behaviour in Zambia, 1996-2003. Sex Transm Infect 2004;80:ii85–90. https://doi.org/10.1136/sti.2004.012054.

[29] Mensch BS, Hewett PC, Erulkar AS. The reporting of sensitive behavior by adolescents: A methodological experiment in Kenya. Demography 2003;40:247–68. https://doi.org/10.2307/3180800.

[30] Luke N, Clark S, Zulu EM. The relationship history calendar: improving the scope and quality of data on youth sexual behavior. Demography 2011;48:1151–76. https://doi.org/10.1007/s13524-011-0051-2.

[31] Cremin I, Mushati P, Hallett T, Mupambireyi Z, Nyamukapa C, Garnett GP, et al. Measuring trends in age at first sex and age at marriage in Manicaland, Zimbabwe. Sex Transm Infect 2009;85:i34–40. https://doi.org/10.1136/sti.2008.033431.

[32] Slaymaker E, Scott RH, Palmer MJ, Palla L, Marston M, Gonsalves L, et al. Trends in sexual activity and demand for and use of modern contraceptive methods in 74 countries: a retrospective analysis of nationally representative surveys. Lancet Glob Health 2020;8:e567–79. https://doi.org/10.1016/S2214-109X(20)30060-7.

[33] Nguyen VK, Eaton JW. A model for reconstructing trends and distribution in age at first sex from multiple household surveys with reporting biases. 2021. https://doi.org/10.1101/2021.12.14.21267770.

[34] ICF. Demographic and Health Surveys (various). Funded USAID Rockv Md ICF Distrib 2020.

[35] UNICEF. Multiple Indicator Cluster Surveys (MICS). 2020.

[36] Elizabeth Heger Boyle, Devon Kristianse, Matthew Sobek. IPUMS PMA: Version 4.1 [dataset]. Minneapolis, MN: IPUMS. 2020. https://doi.org/10.18128/D081.V4.1.

[37] UNICEF. Eritrea Population and Health Survey. 2020.

[38] PHIA Collaborating Institutions. Population-based HIV Impact Assessment (PHIA) 2020. https://doi.org/10.18128/D081.V4.1.

[39] MEASURE Evaluation Project. Zambia sexual behaviour surveys. 2020. https://doi.org/10.18128/D081.V4.1.

[40] The Human Sciences Research Council. South African National HIV Prevalence, Incidence and Behaviour Survey. 2020. https://doi.org/10.18128/D081.V4.1.

[41] Asparouhov T. General Multi-Level Modeling with Sampling Weights. Commun Stat -Theory Methods 2006;35:439–60. https://doi.org/10.1080/03610920500476598.

[42] Van Kinh Nguyen, Jeffrey W Eaton. Model the age at first sex distribution and age at reporting bias in household survey data. MedRxiv 2021.

[43] Besag J, Kooperberg C. On Conditional and Intrinsic Autoregression. Biometrika 1995;82:733–46. https://doi.org/10.2307/2337341.

[44] Kristensen K, Nielsen A, Berg CW, Skaug H, Bell BM. TMB: Automatic differentiation and laplace approximation. J Stat Softw 2016;70. https://doi.org/10.18637/jss.v070.i05.

[45] Melesse DY, Mutua MK, Choudhury A, Wado YD, Faye CM, Neal S, et al. Adolescent sexual and reproductive health in sub-Saharan Africa: who is left behind? BMJ Glob Health 2020;5:e002231. https://doi.org/10.1136/bmjgh-2019-002231.

[46] Slaymaker E, Bwanika JB, Kasamba I, Lutalo T, Maher D, Todd J. Trends in age at first sex in Uganda: evidence from Demographic and Health Survey data and longitudinal cohorts in Masaka and Rakai. Sex Transm Infect 2009;85:i12–9. https://doi.org/10.1136/sti.2008.034009.

[47] Shapiro D, Gebreselassie T. Marriage in Sub-Saharan Africa: Trends, Determinants, and Consequences. Popul Res Policy Rev 2014;33:229–55. https://doi.org/10.1007/s11113-013-9287-4.

[48] Dwyer-Lindgren L, Cork MA, Sligar A, Steuben KM, Wilson KF, Provost NR, et al. Mapping HIV prevalence in sub-Saharan Africa between 2000 and 2017. Nature 2019;570:189–93. https://doi.org/10.1038/s41586-019-1200-9.

[49] Barro Robert, Jong-Wha Lee. A New Data Set of Educational Attainment in the World, 1950-2010. J Dev Econ 2013;104:184–98. https://doi.org/10.1016/j.jdeveco.2012.10.001.

[50] Bongaarts J, Mensch BS, Blanc AK. Trends in the age at reproductive transitions in the developing world: The role of education. Popul Stud 2017;71:139–54. https://doi.org/10.1080/00324728.2017.1291986.

[51] Smith DJ. Masculinity, Money, and the Postponement of Parenthood in Nigeria. Popul Dev Rev 2020;46:101–20. https://doi.org/10.1111/padr.12310.

[52] Plummer ML, Ross DA, Wight D, Changalucha J, Mshana G, Wamoyi J, et al. “A bit more truthful”: The validity of adolescent sexual behaviour data collected in rural northern Tanzania using five methods. Sex. Transm. Infect., vol. 80, 2004, p. ii49–56. https://doi.org/10.1136/sti.2004.011924.

[53] Amos M, Stones T. Trends in Demographic and Health Survey data quality: An analysis of age heaping over time in 34 countries in Sub Saharan Africa between 1987 and 2015. BMC Res Notes 2017;10. https://doi.org/10.1186/s13104-017-3091-x.

[54] Westoff, Charles F. K Bietsch. Religion and Reproductive Behavior in Sub Saharan Africa. DHS Anal Stud No 48 2015.

[55] Halperin DT, Steiner MJ, Cassell MM, Green EC, Hearst N, Kirby D, et al. The time has come for common ground on preventing sexual transmission of HIV. The Lancet 2004;364:1913–5. https://doi.org/10.1016/s0140-6736(04)17487-4.

