## Supplementary material for "Trends and country-level variation in age at first sex in sub-Saharan Africa among birth cohorts entering adulthood between 1985 and 2020": Figure_S3_female.pdf

### Angola - female

#### Data and model's prediction

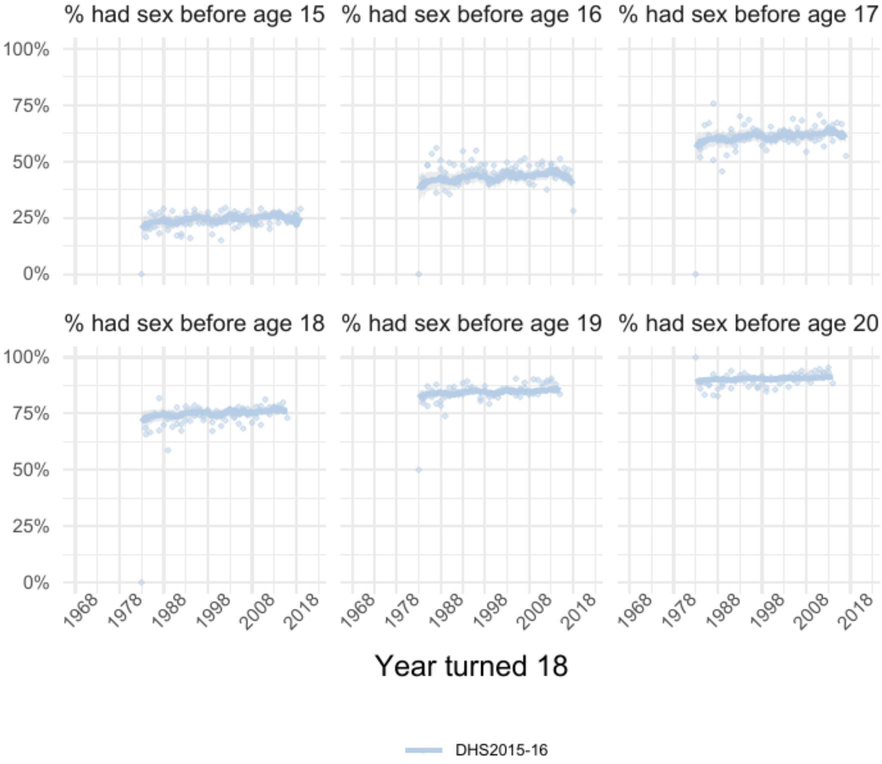


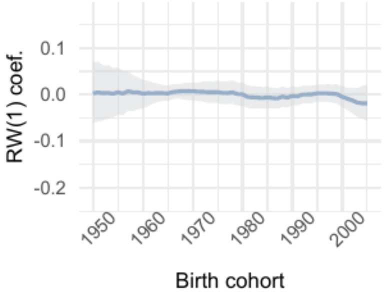


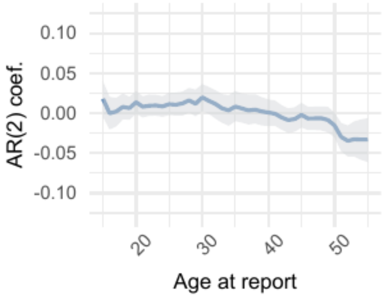


| | Parameter | Estimate |
|---|---|---|
| 1 | intercept | 0.06 [0.04 – 0.08] |
| 2 | skew | 1.94 [1.80 – 2.11] |
| 3 | shape | 10.46 [10.21 – 10.69] |

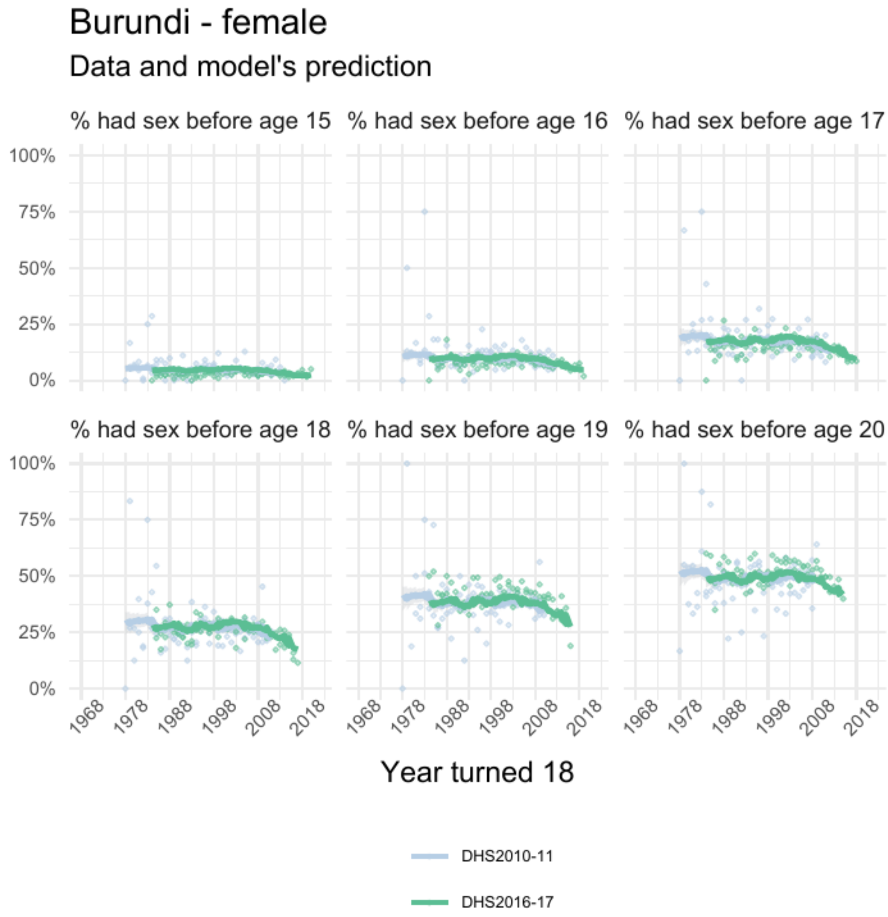


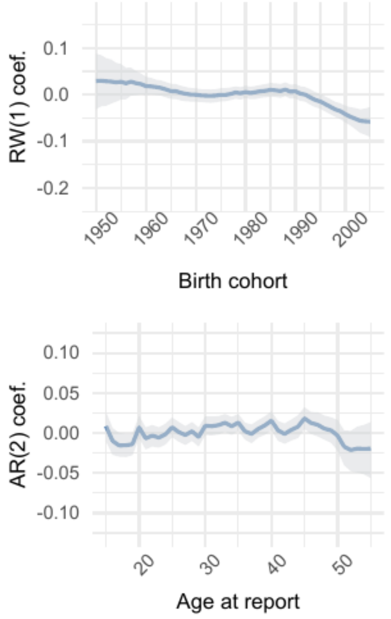


| | Parameter | Estimate |
|---|---|---|
| 1 | intercept | -0.06 [-0.07 – -0.04] |
| 2 | skew | 2.59 [2.46 – 2.73] |
| 3 | shape | 6.83 [6.73 – 6.92] |

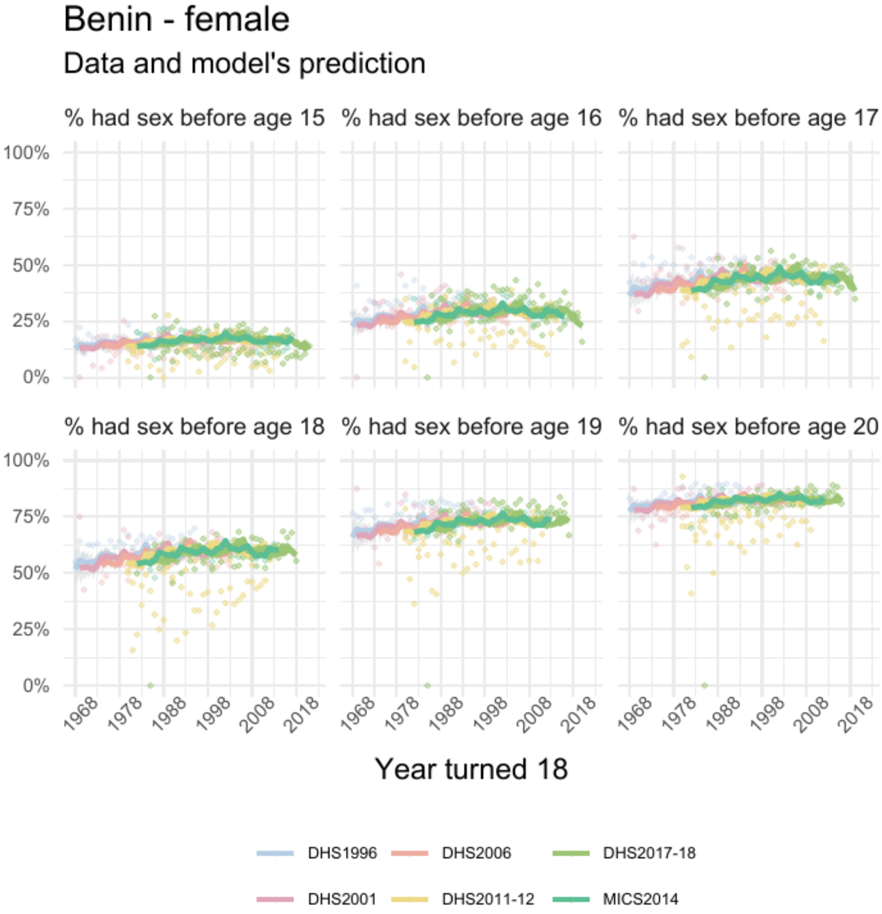


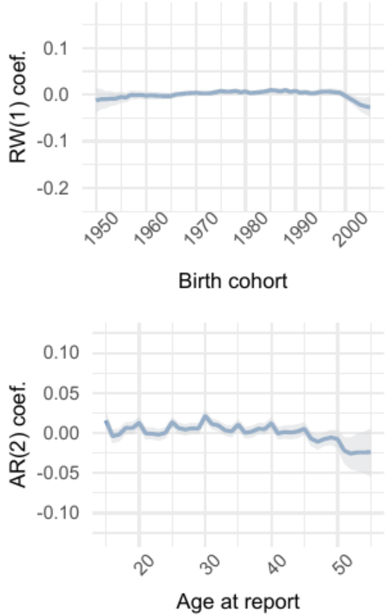


| | Parameter | Estimate |
|---|---|---|
| *1* | intercept | -0.09 [-0.10 – -0.09] |
| *2* | skew | 0.96 [0.93 – 0.99] |
| *3* | shape | 11.15 [11.01 – 11.30] |

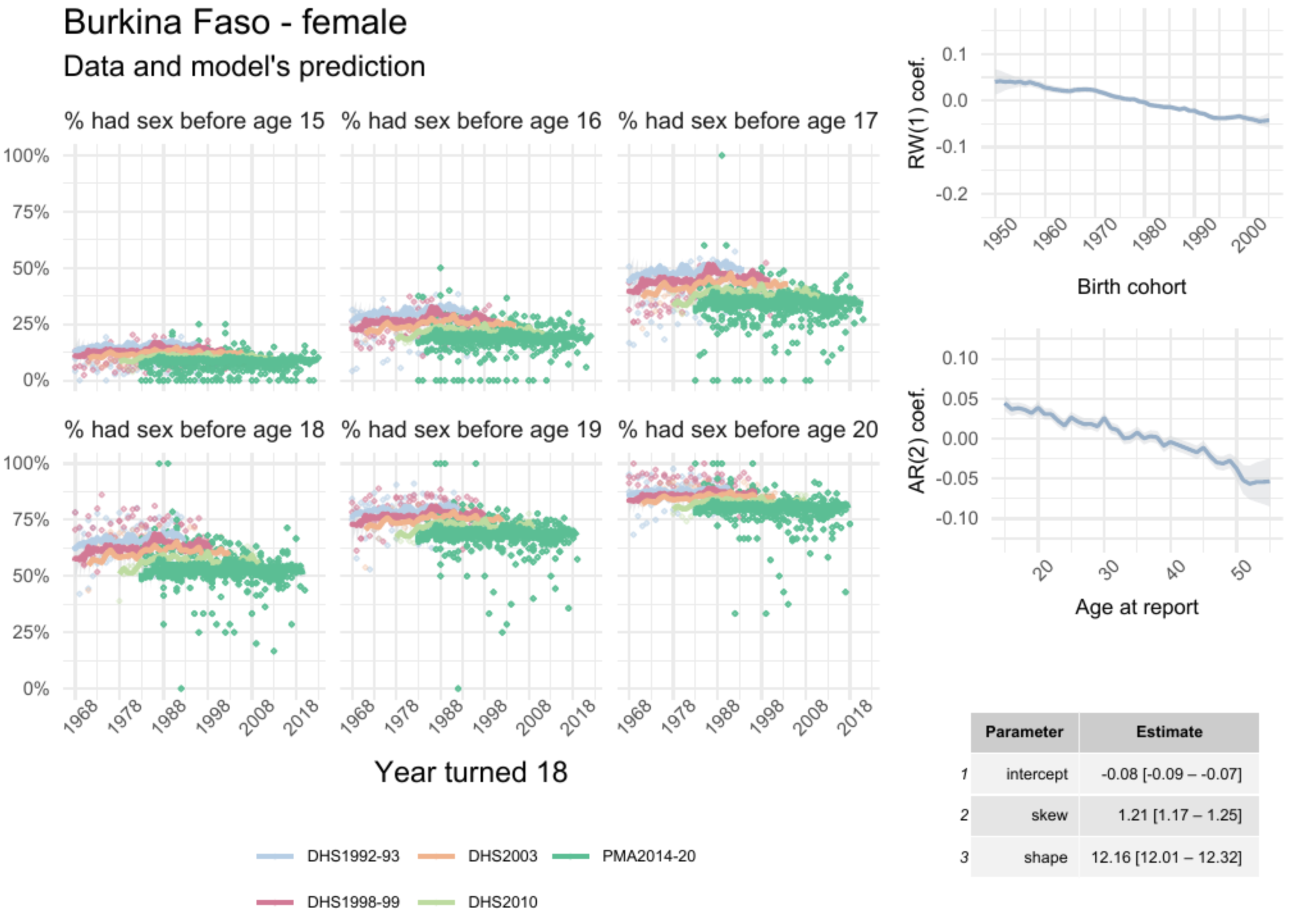


| | Parameter | Estimate |
|---|---|---|
| 1 | intercept | -0.08 [-0.09 – -0.07] |
| 2 | skew | 1.21 [1.17 – 1.25] |
| 3 | shape | 12.16 [12.01 – 12.32] |

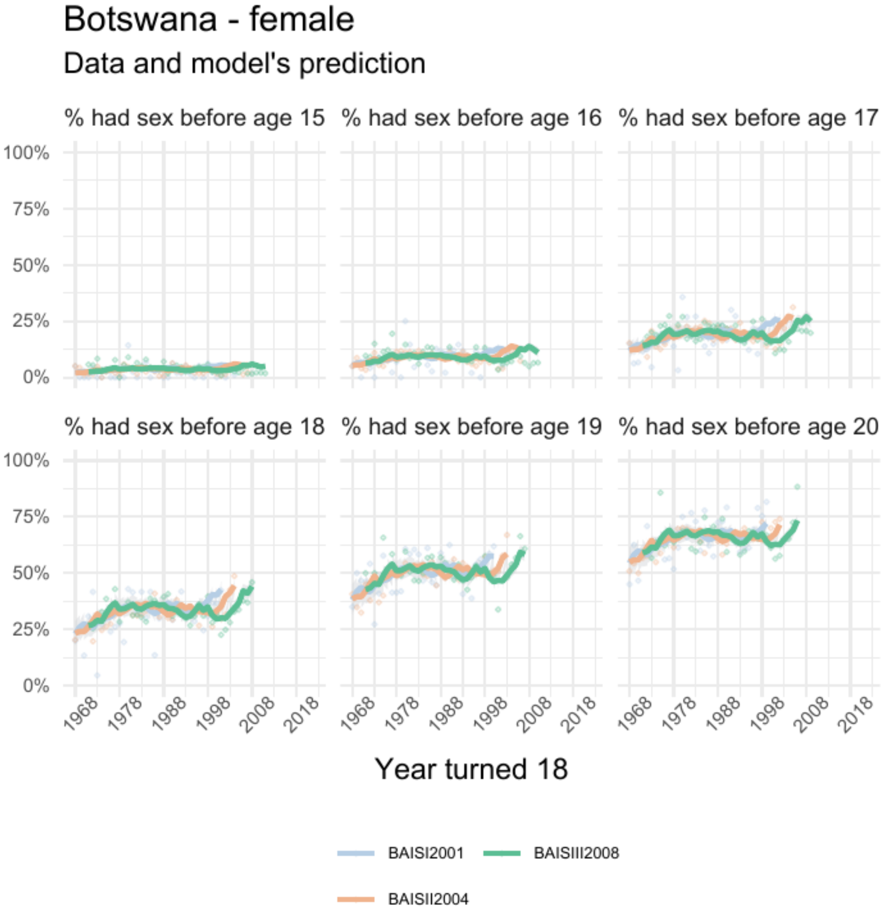


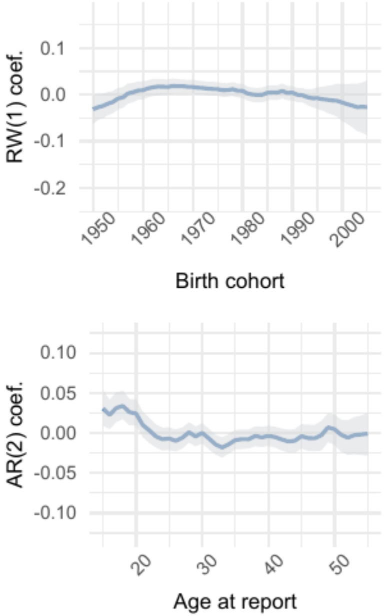


| | Parameter | Estimate |
|---|---|---|
| 1 | intercept | -0.14 [-0.15 – -0.13] |
| 2 | skew | 1.31 [1.23 – 1.40] |
| 3 | shape | 11.92 [11.64 – 12.21] |

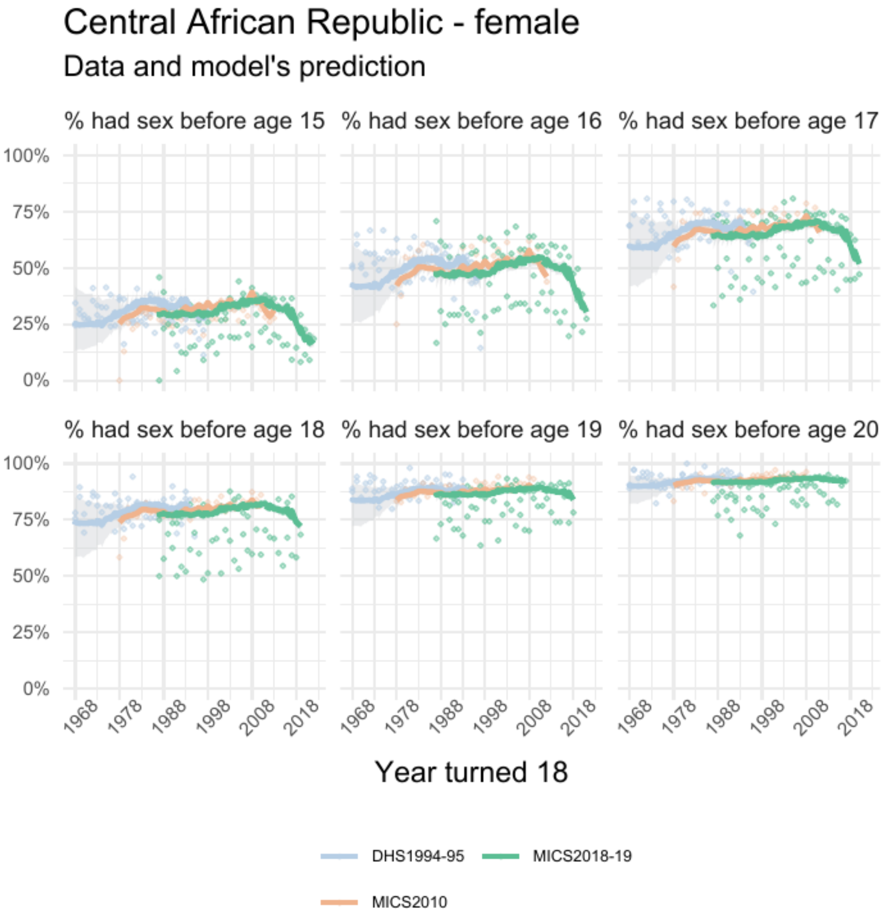


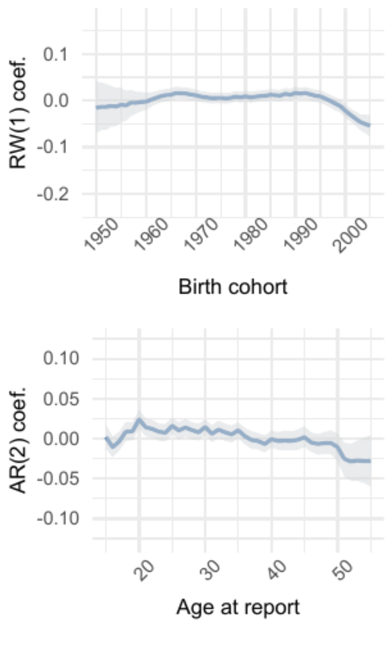


| | Parameter | Estimate |
|---|---|---|
| 1 | intercept | 0.016 [0.004 – 0.028] |
| 2 | skew | 1.24 [1.17 – 1.30] |
| 3 | shape | 10.95 [10.73 – 11.18] |

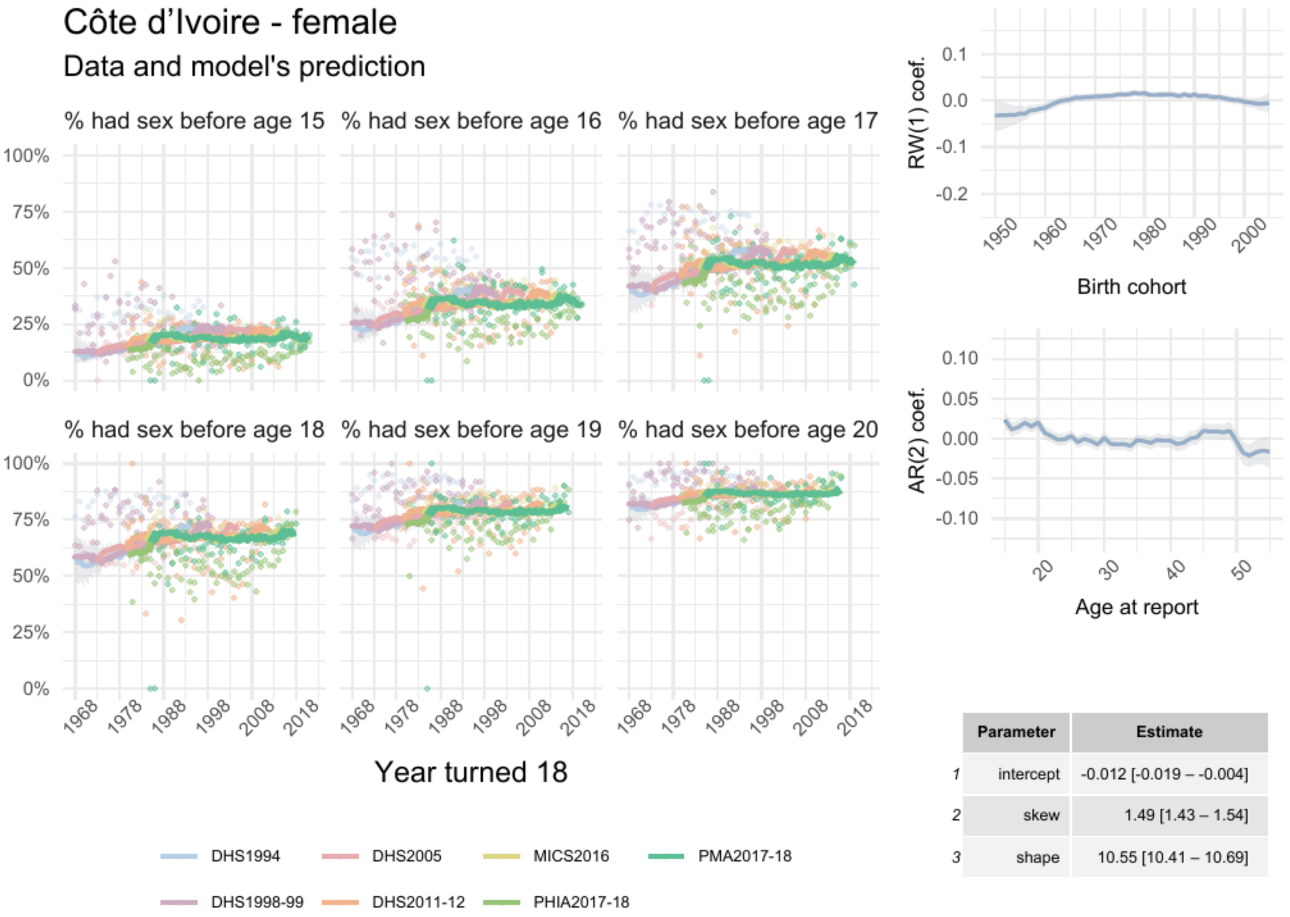


| | Parameter | Estimate |
|---|---|---|
| 1 | intercept | -0.012 [-0.019 – -0.004] |
| 2 | skew | 1.49 [1.43 – 1.54] |
| 3 | shape | 10.55 [10.41 – 10.69] |

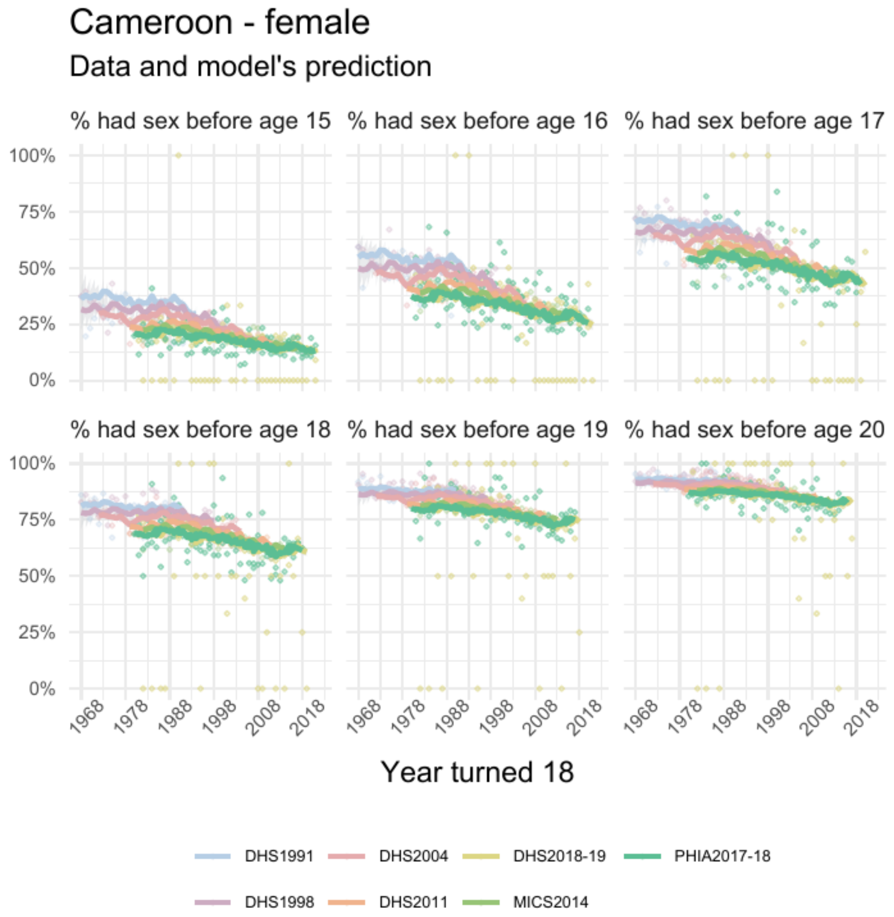


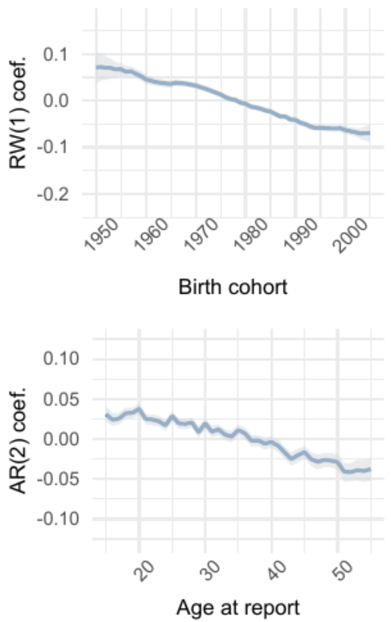


| | Parameter | Estimate |
|---|---|---|
| 1 | intercept | 0.03 [0.02 – 0.03] |
| 2 | skew | 1.70 [1.64 – 1.76] |
| 3 | shape | 10.04 [ 9.93 – 10.17] |

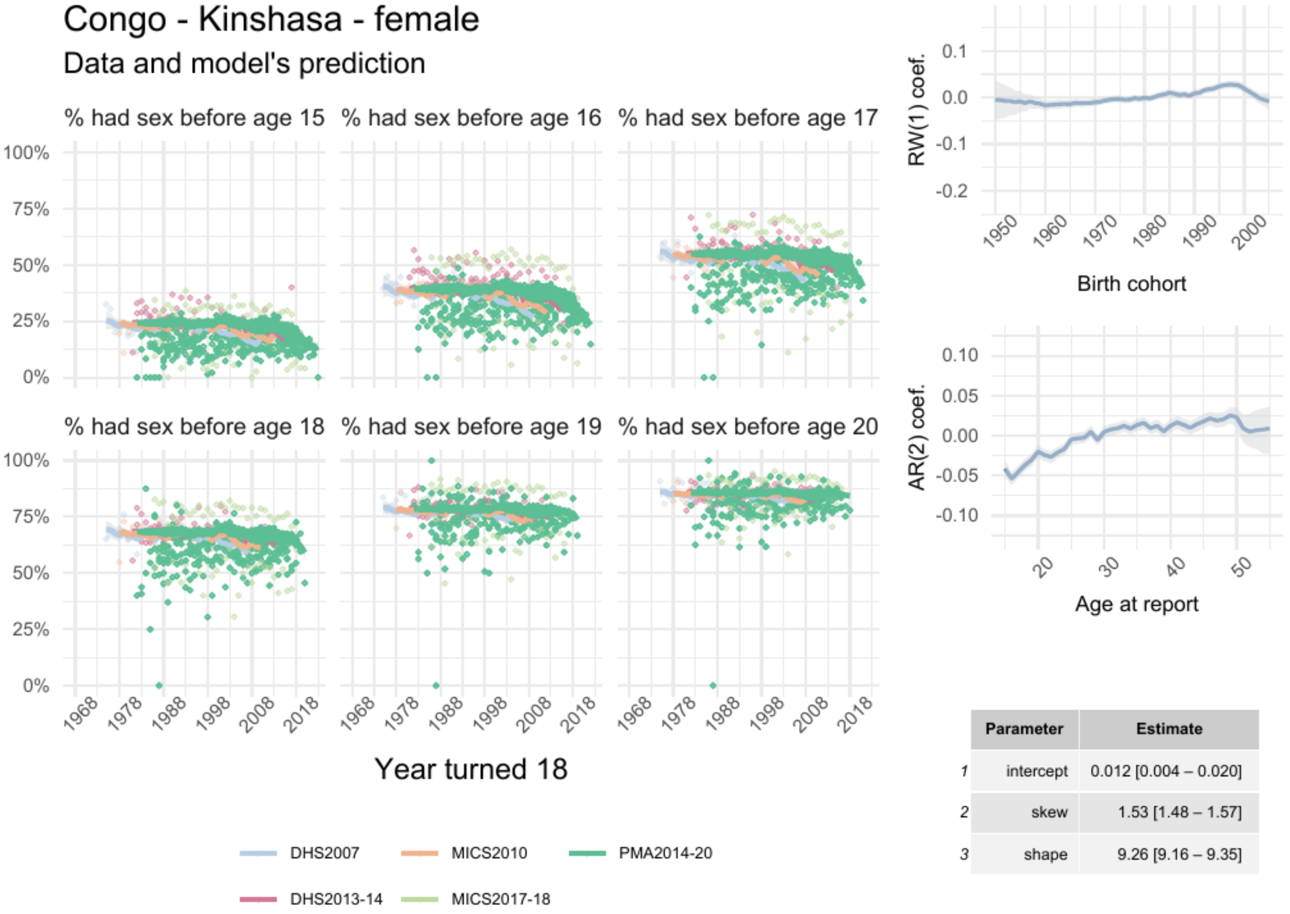


| | Parameter | Estimate |
|---|---|---|
| 1 | intercept | 0.012 [0.004 – 0.020] |
| 2 | skew | 1.53 [1.48 – 1.57] |
| 3 | shape | 9.26 [9.16 – 9.35] |

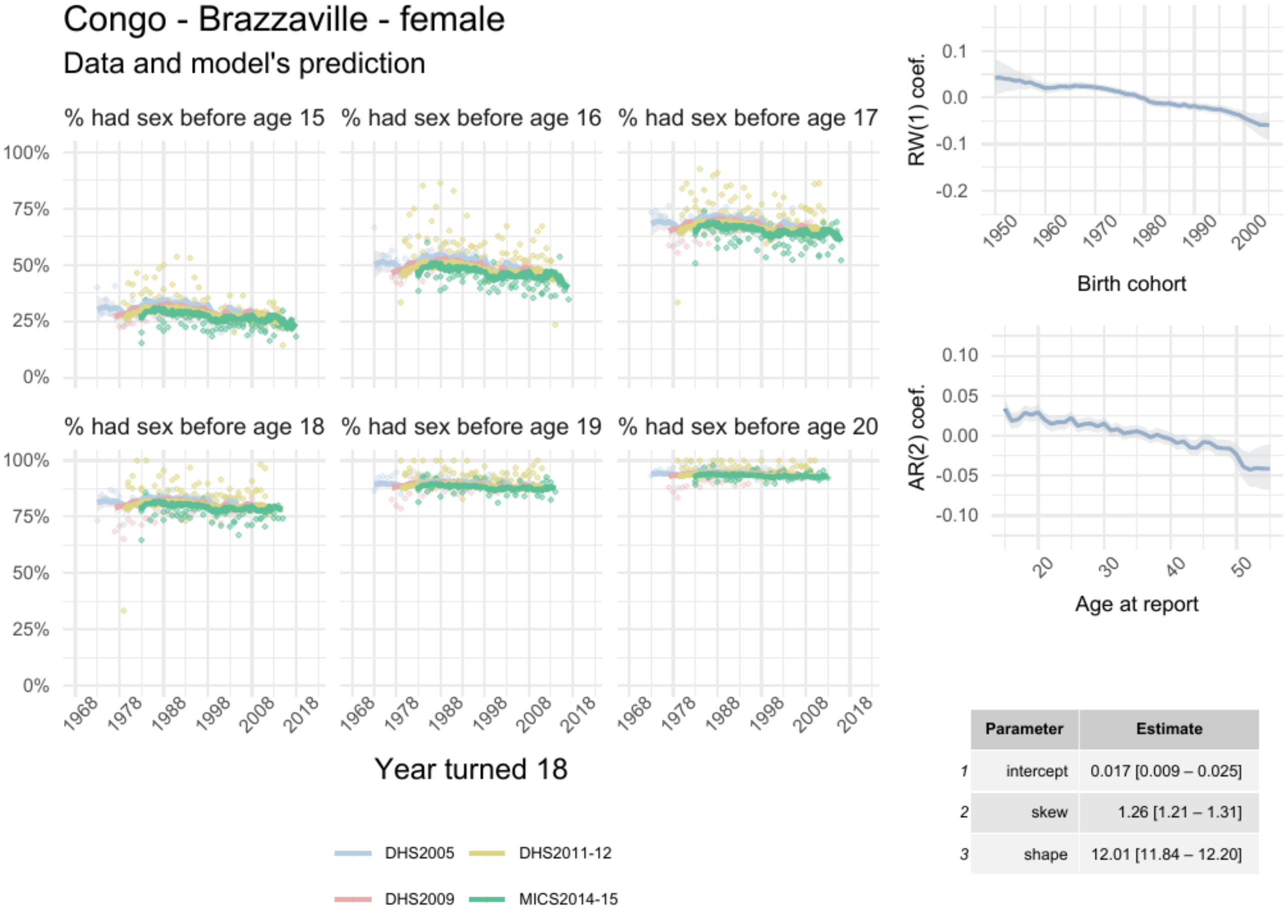


| | Parameter | Estimate |
|---|---|---|
| 1 | intercept | 0.017 [0.009 – 0.025] |
| 2 | skew | 1.26 [1.21 – 1.31] |
| 3 | shape | 12.01 [11.84 – 12.20] |

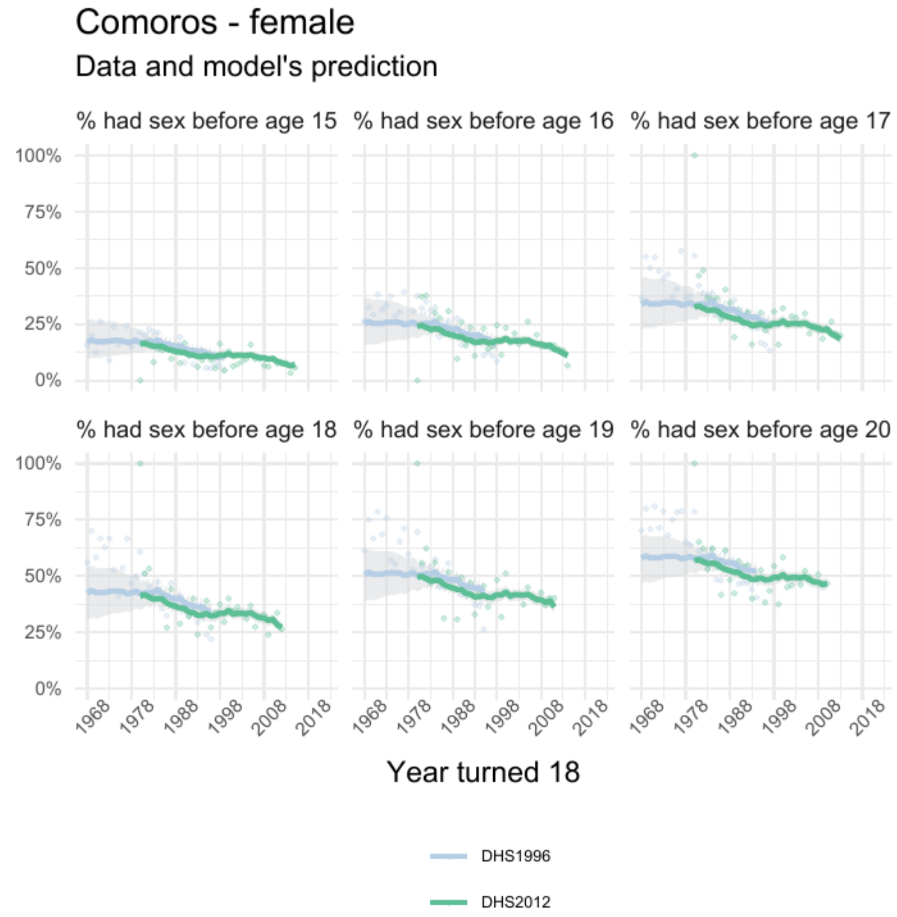


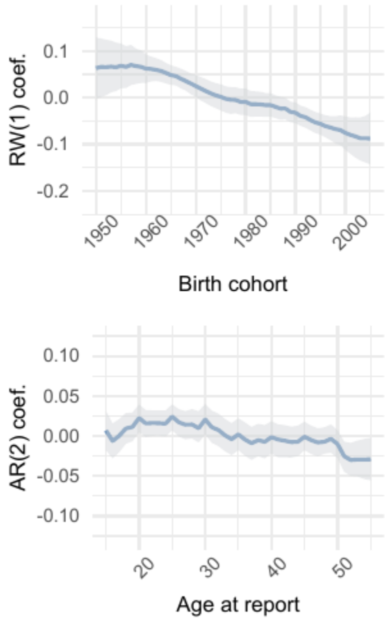


| | Parameter | Estimate |
|---|---|---|
| 1 | intercept | 0.10 [0.05 – 0.15] |
| 2 | skew | 3.52 [2.92 – 4.22] |
| 3 | shape | 4.73 [4.55 – 4.91] |

### Eritrea - female

#### Data and model's prediction

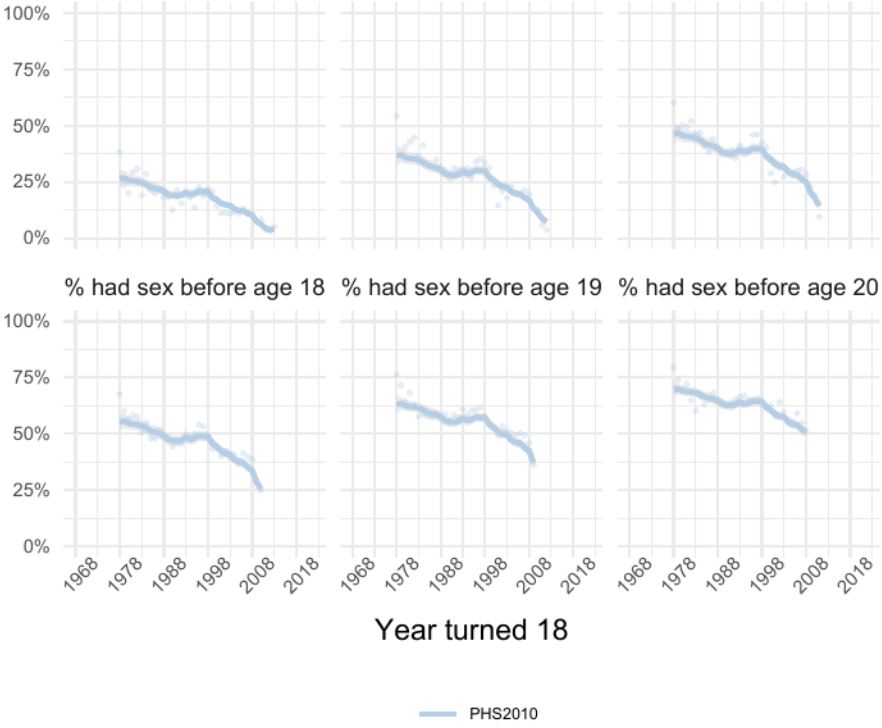


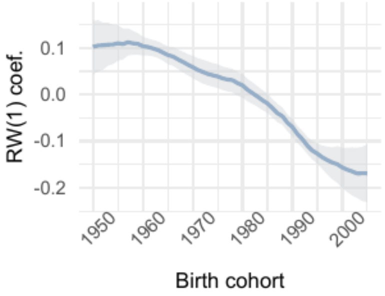


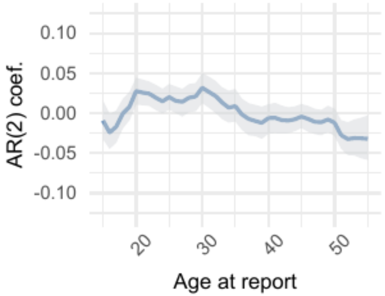


| | Parameter | Estimate |
|---|---|---|
| 1 | intercept | 0.16 [0.12 – 0.21] |
| 2 | skew | 3.98 [3.43 – 4.66] |
| 3 | shape | 4.96 [4.83 – 5.10] |

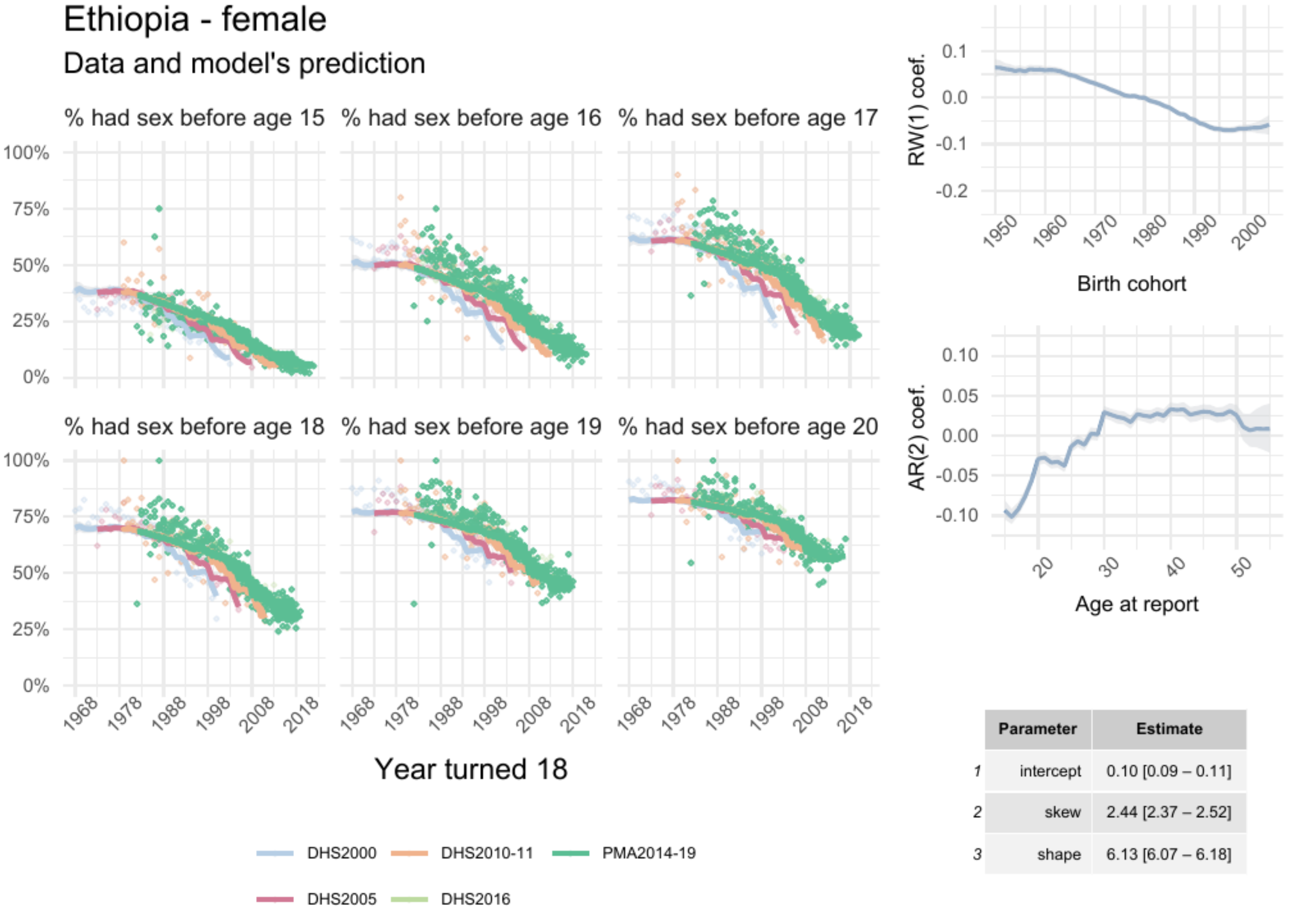


| | Parameter | Estimate |
|---|---|---|
| 1 | intercept | 0.10 [0.09 – 0.11] |
| 2 | skew | 2.44 [2.37 – 2.52] |
| 3 | shape | 6.13 [6.07 – 6.18] |

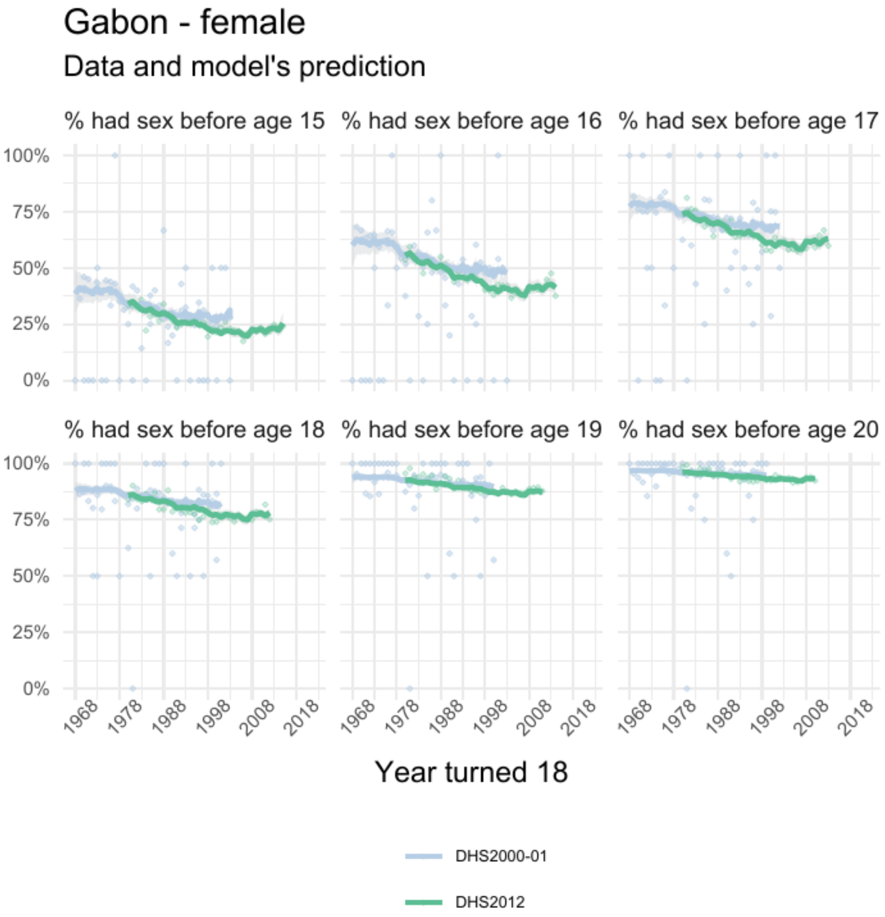


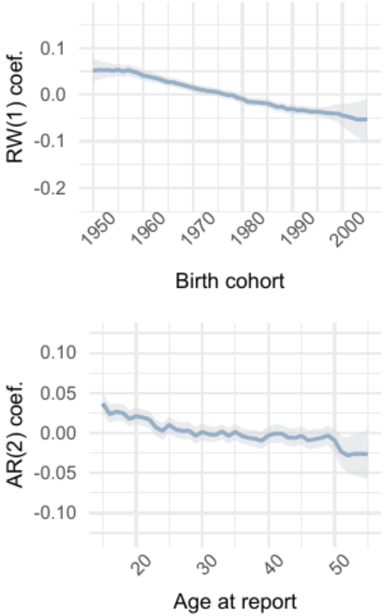


| | Parameter | Estimate |
|---|---|---|
| 1 | intercept | 0.007 [-0.004 – 0.016] |
| 2 | skew | 1.15 [1.08 – 1.22] |
| 3 | shape | 13.07 [12.77 – 13.41] |

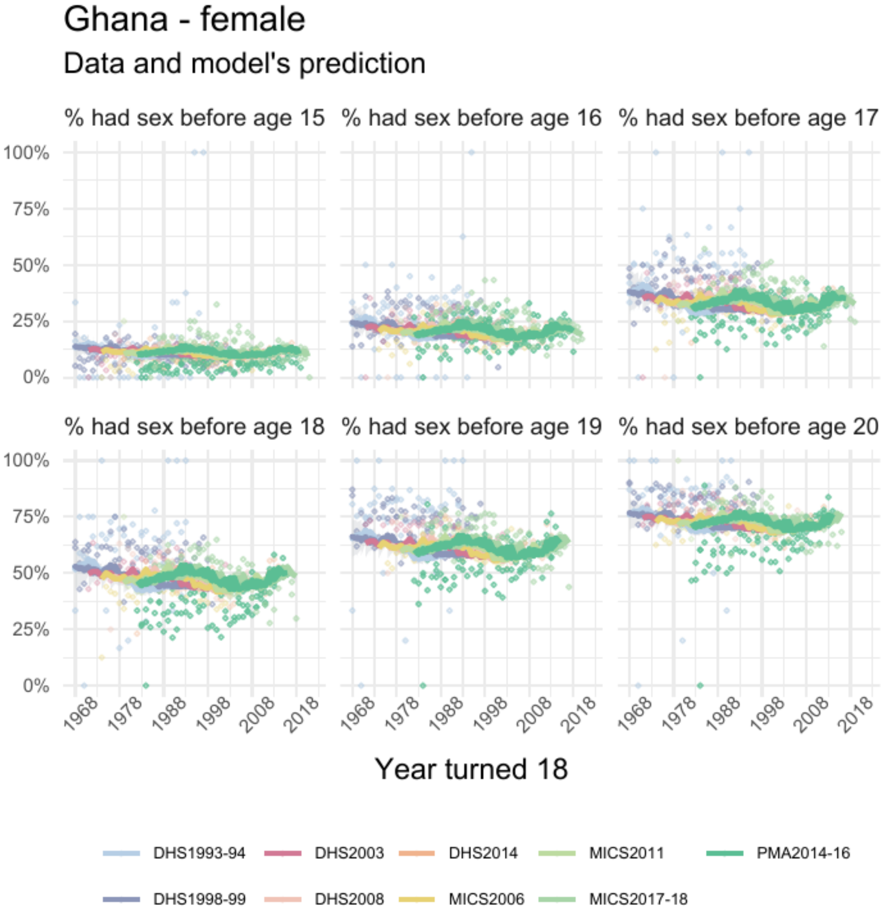


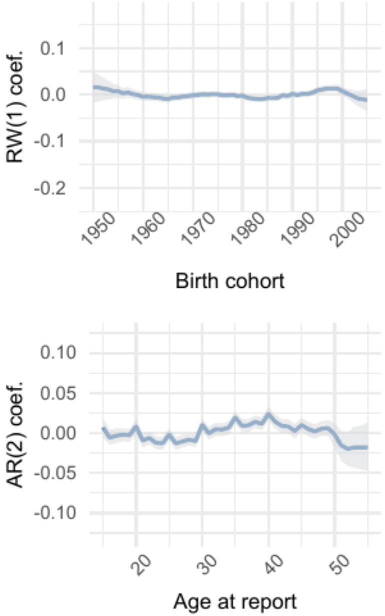


| | Parameter | Estimate |
|---|---|---|
| 1 | intercept | -0.10 [-0.11 – -0.09] |
| 2 | skew | 1.19 [1.16 – 1.23] |
| 3 | shape | 9.88 [ 9.77 – 10.02] |

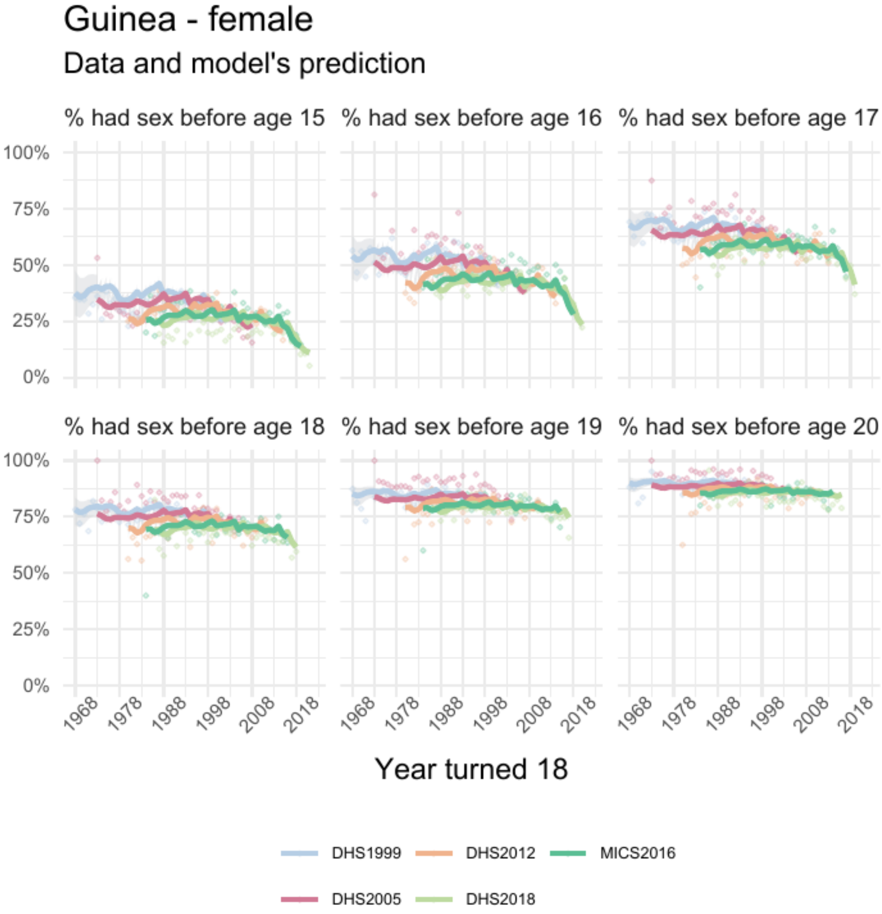


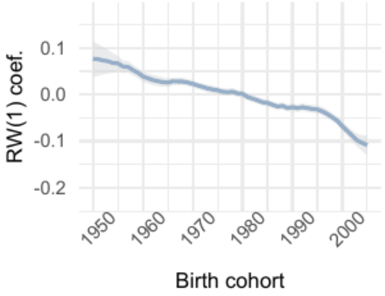


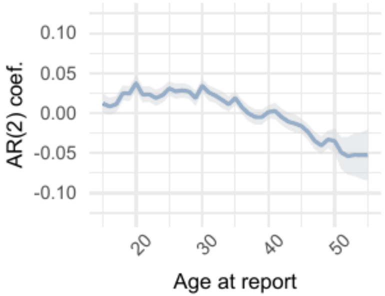


| | Parameter | Estimate |
|---|---|---|
| 1 | intercept | 0.11 [0.10 – 0.12] |
| 2 | skew | 2.45 [2.32 – 2.61] |
| 3 | shape | 8.41 [8.29 – 8.53] |

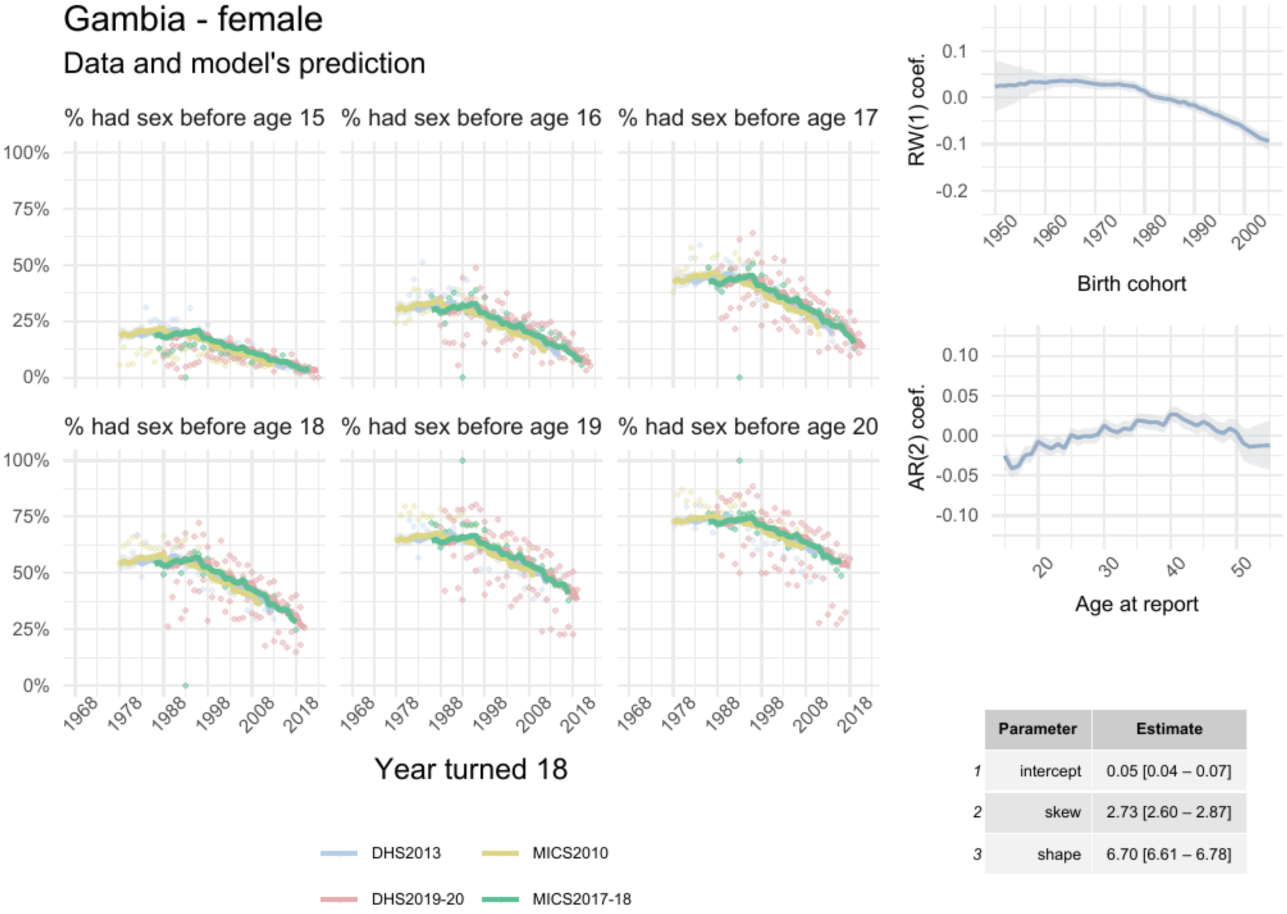


| | Parameter | Estimate |
|---|---|---|
| *1* | intercept | 0.05 [0.04 – 0.07] |
| *2* | skew | 2.73 [2.60 – 2.87] |
| *3* | shape | 6.70 [6.61 – 6.78] |

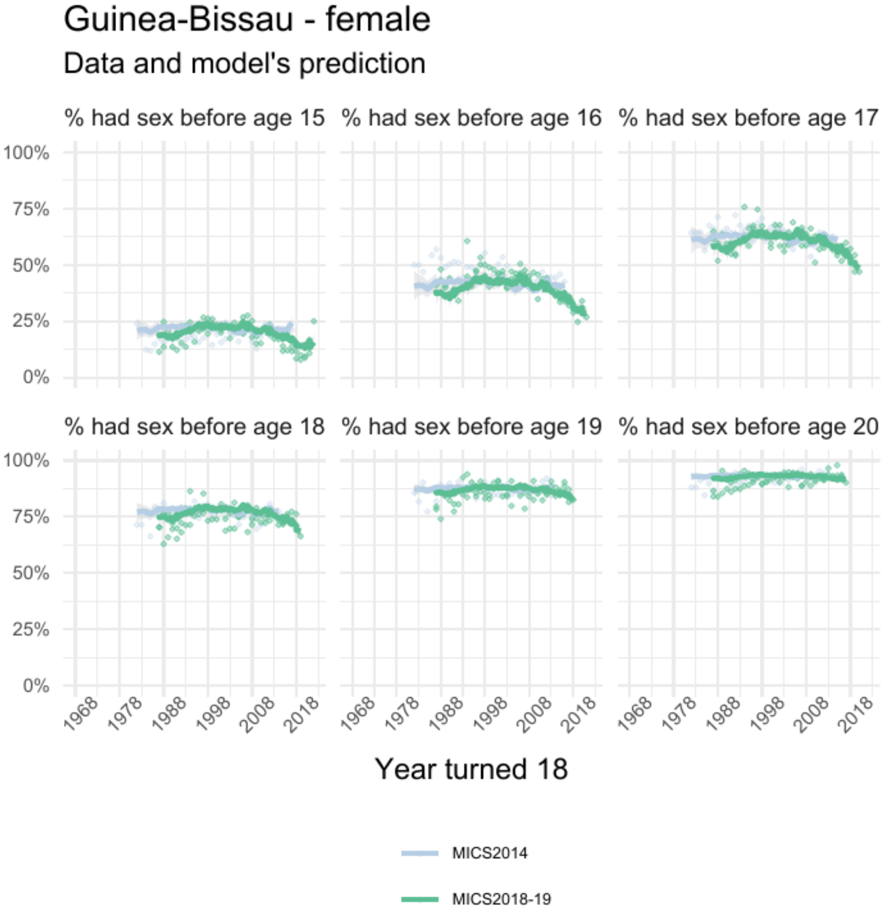


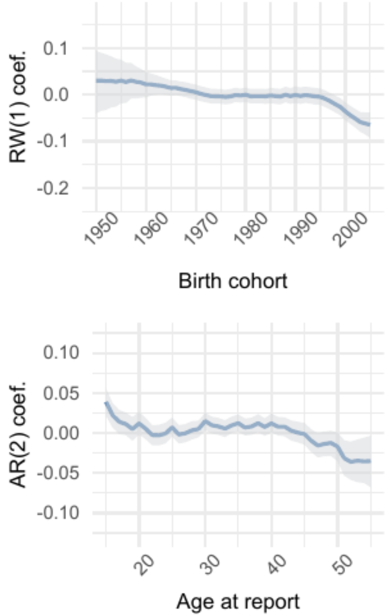


| | Parameter | Estimate |
|---|---|---|
| *1* | intercept | 0.013 [-0.002 – 0.026] |
| *2* | skew | 1.42 [1.35 – 1.49] |
| *3* | shape | 12.62 [12.39 – 12.86] |

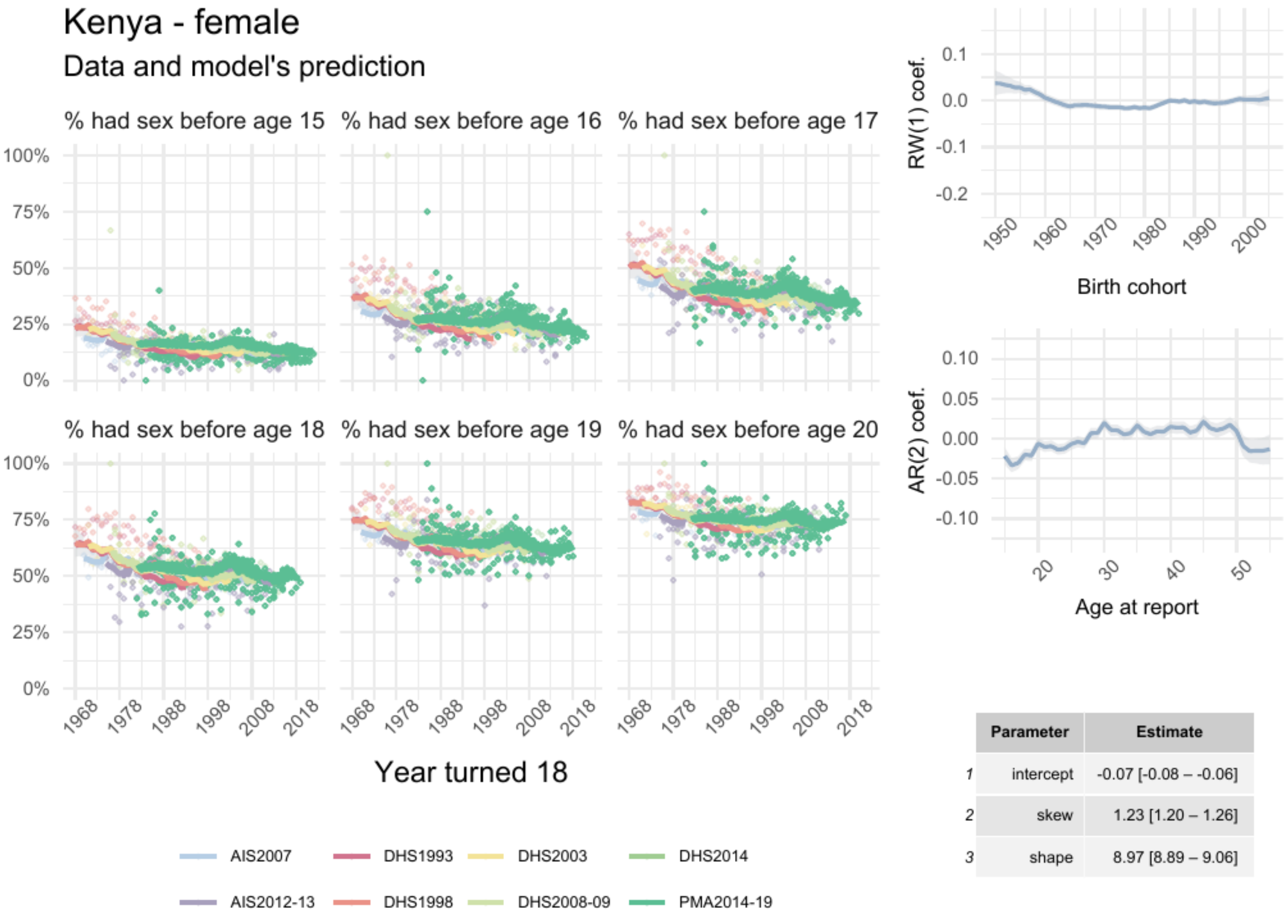


| | Parameter | Estimate |
|---|---|---|
| 1 | intercept | -0.07 [-0.08 – -0.06] |
| 2 | skew | 1.23 [1.20 – 1.26] |
| 3 | shape | 8.97 [8.89 – 9.06] |

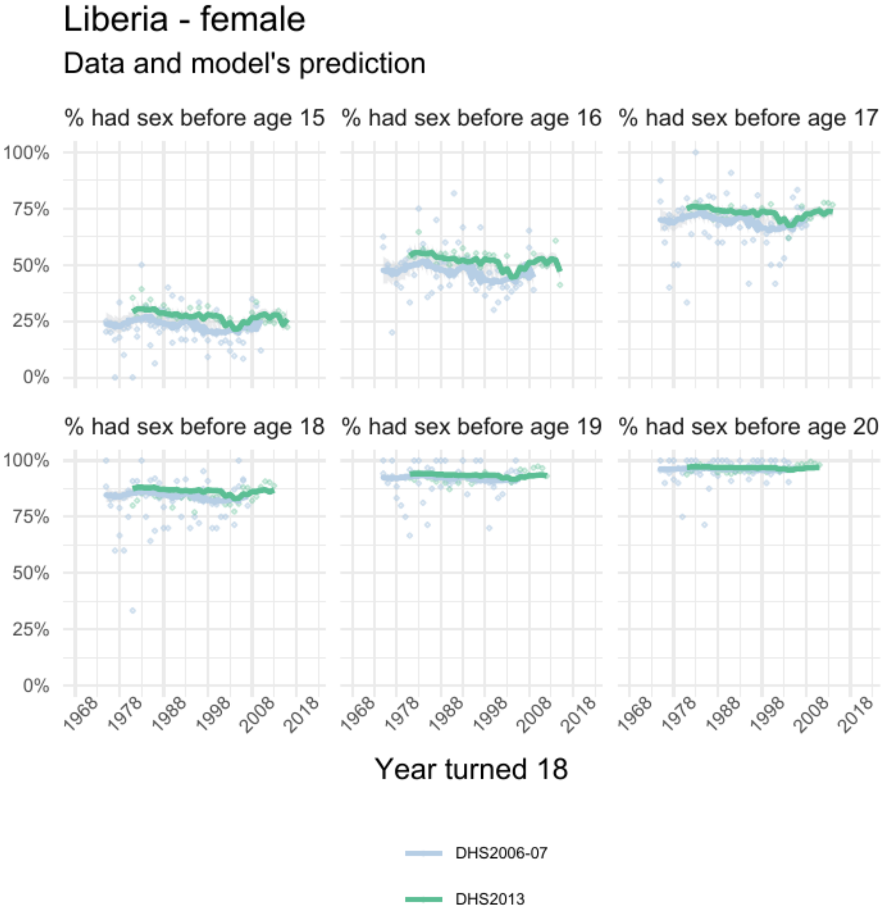


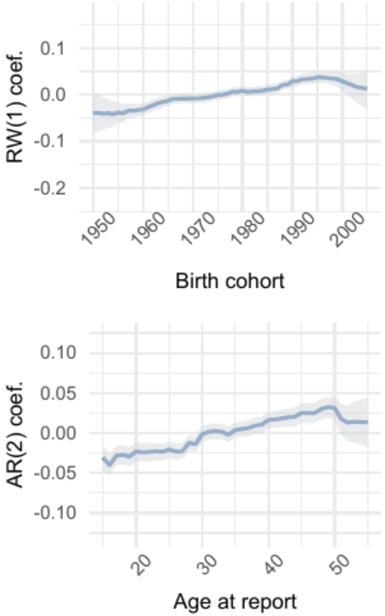


| | Parameter | Estimate |
|---|---|---|
| 1 | intercept | 0.03 [0.02 – 0.04] |
| 2 | skew | 1.42 [1.33 – 1.52] |
| 3 | shape | 14.48 [14.17 – 14.81] |

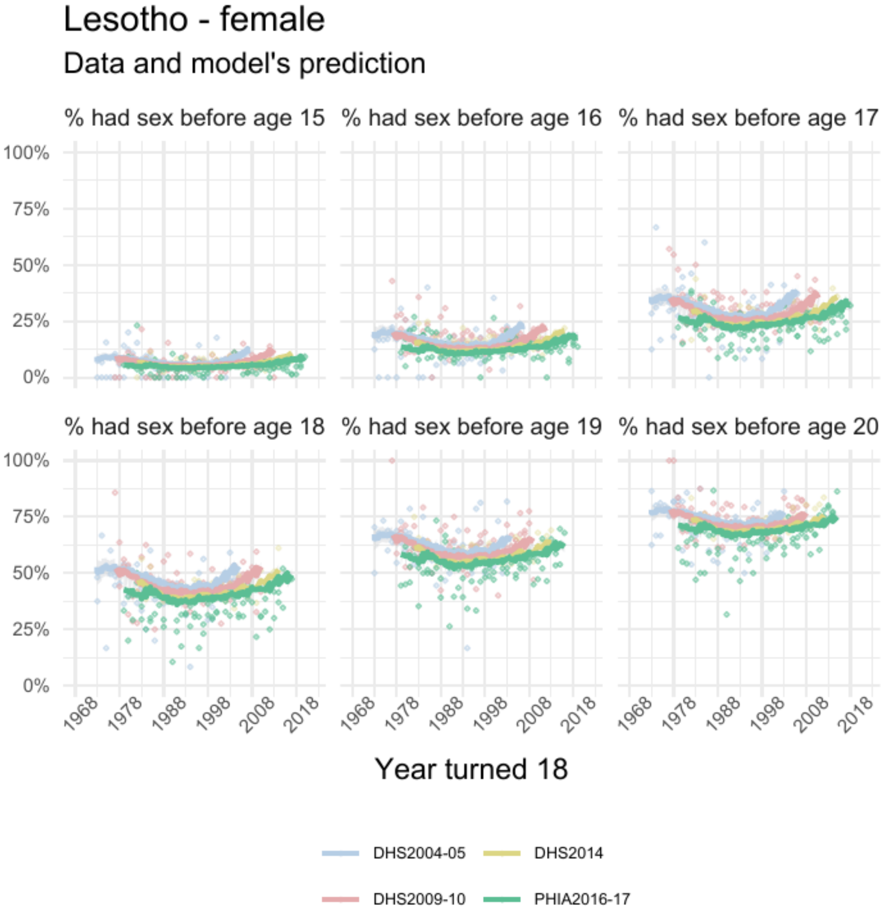


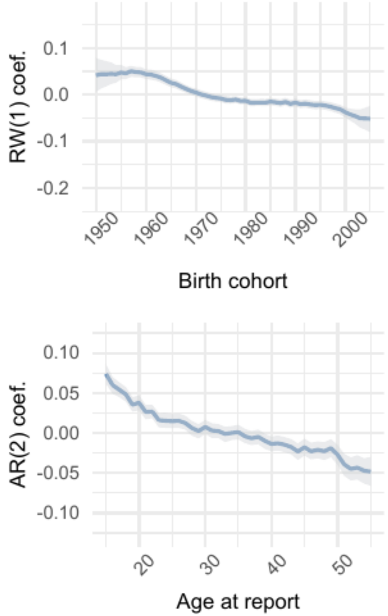


| | Parameter | Estimate |
|---|---|---|
| 1 | intercept | -0.05 [-0.06 – -0.04] |
| 2 | skew | 1.89 [1.78 – 2.00] |
| 3 | shape | 10.12 [ 9.95 – 10.31] |

### Madagascar - female

#### Data and model's prediction

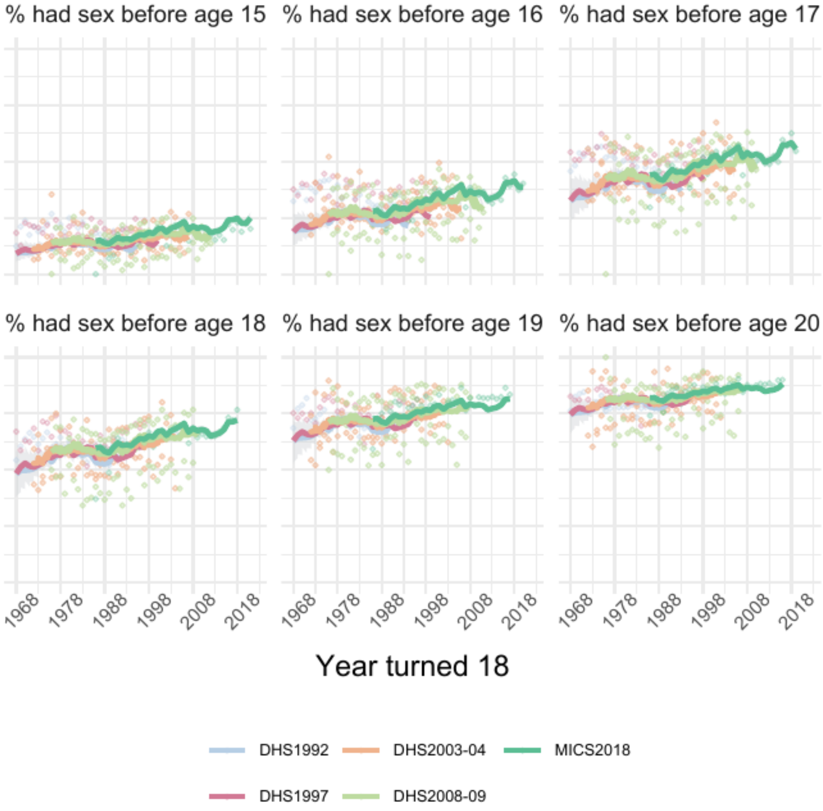


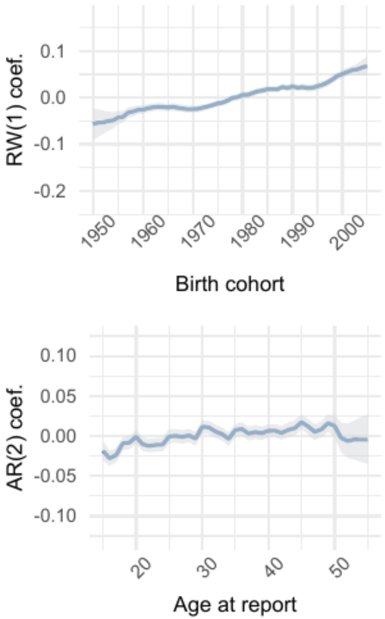


| | Parameter | Estimate |
|---|---|---|
| 1 | intercept | -0.011 [-0.019 – -0.004] |
| 2 | skew | 1.55 [1.48 – 1.61] |
| 3 | shape | 10.00 [ 9.87 – 10.13] |

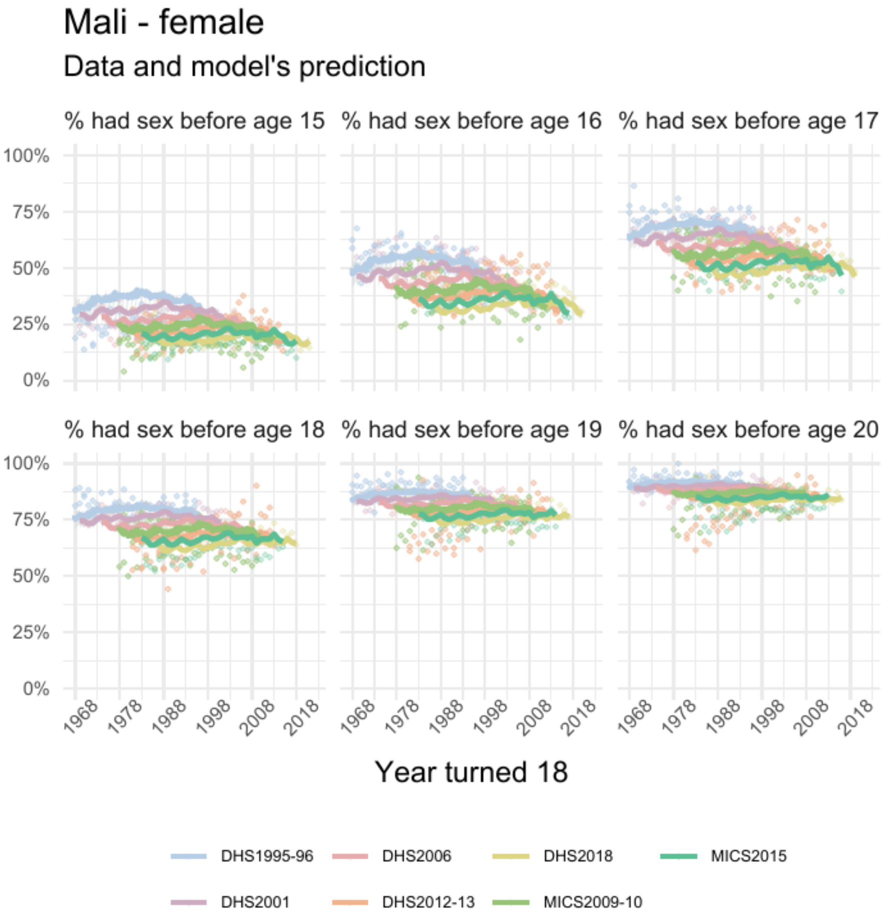


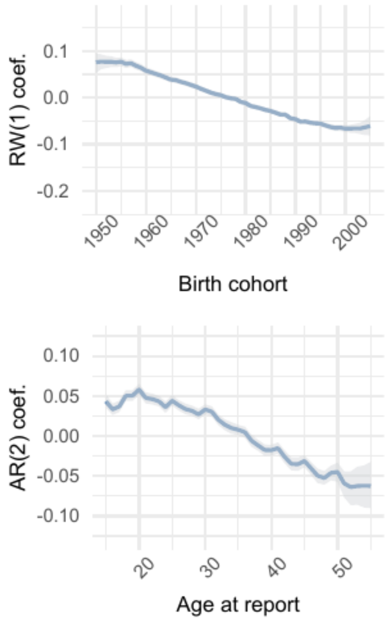


| | Parameter | Estimate |
|---|---|---|
| *1* | intercept | 0.04 [0.04 – 0.05] |
| *2* | skew | 1.83 [1.79 – 1.88] |
| *3* | shape | 9.35 [9.27 – 9.43] |

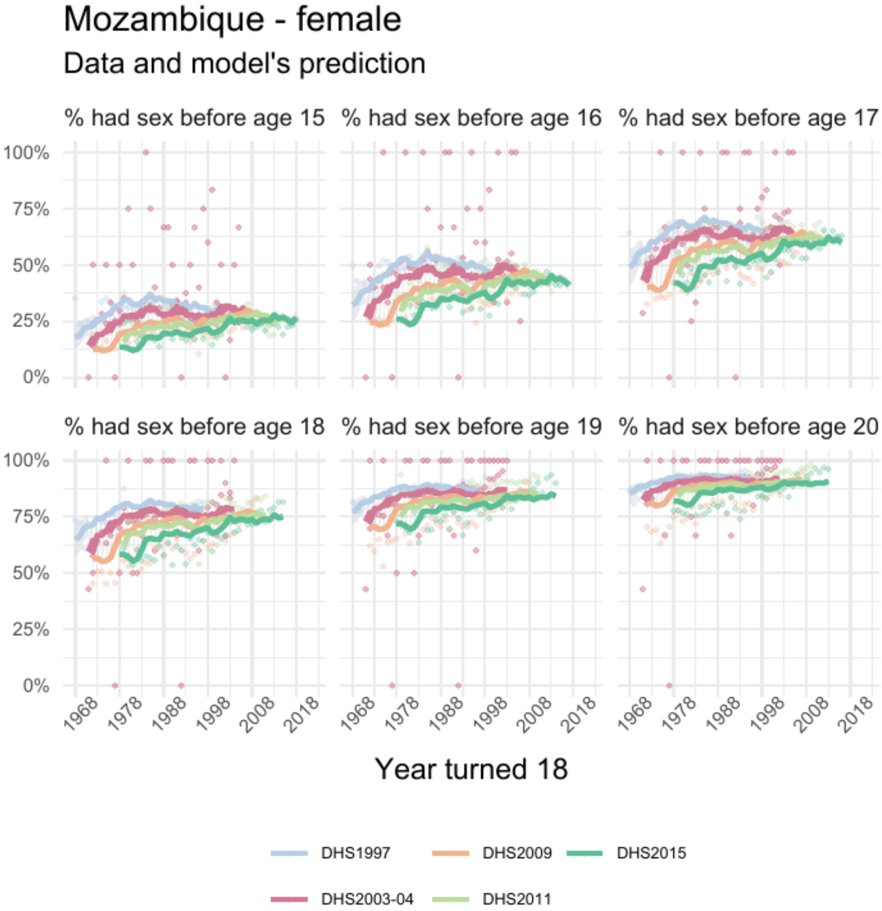


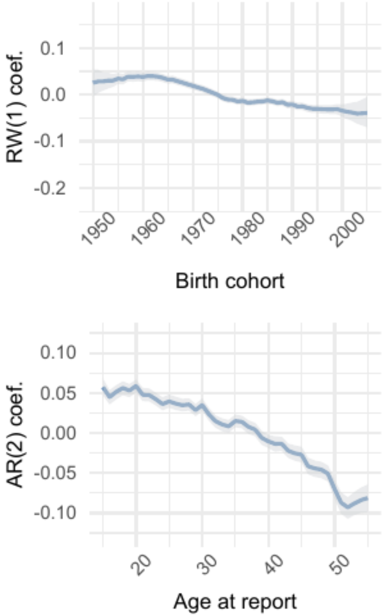


| | Parameter | Estimate |
|---|---|---|
| *1* | intercept | -0.009 [-0.017 – -0.001] |
| *2* | skew | 1.29 [1.24 – 1.35] |
| *3* | shape | 10.77 [10.61 – 10.94] |

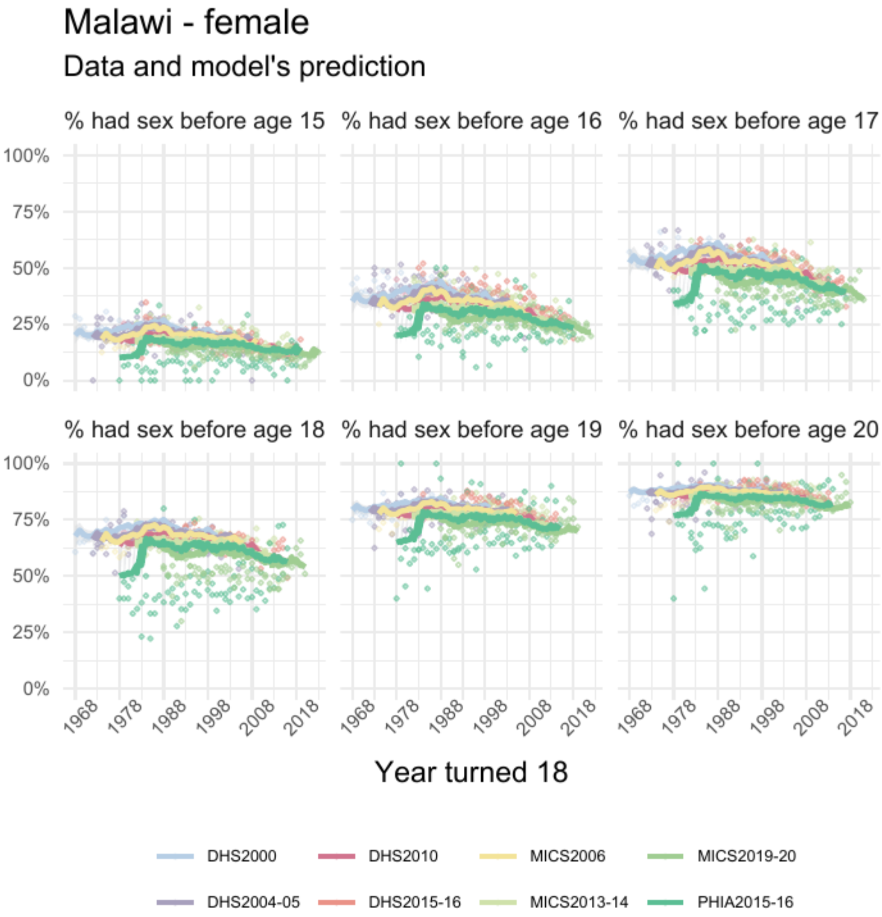


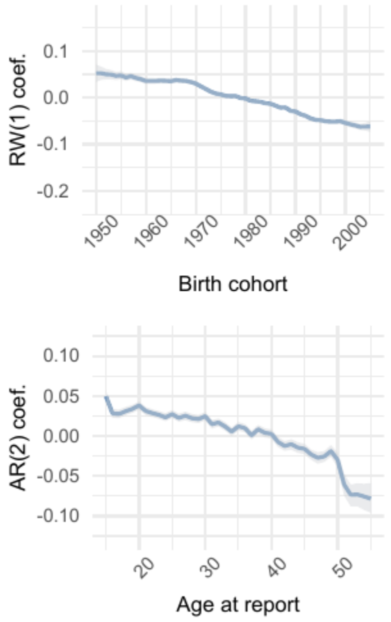


| | Parameter | Estimate |
|---|---|---|
| 1 | intercept | -0.05 [-0.06 – -0.05] |
| 2 | skew | 1.19 [1.16 – 1.21] |
| 3 | shape | 10.97 [10.88 – 11.06] |

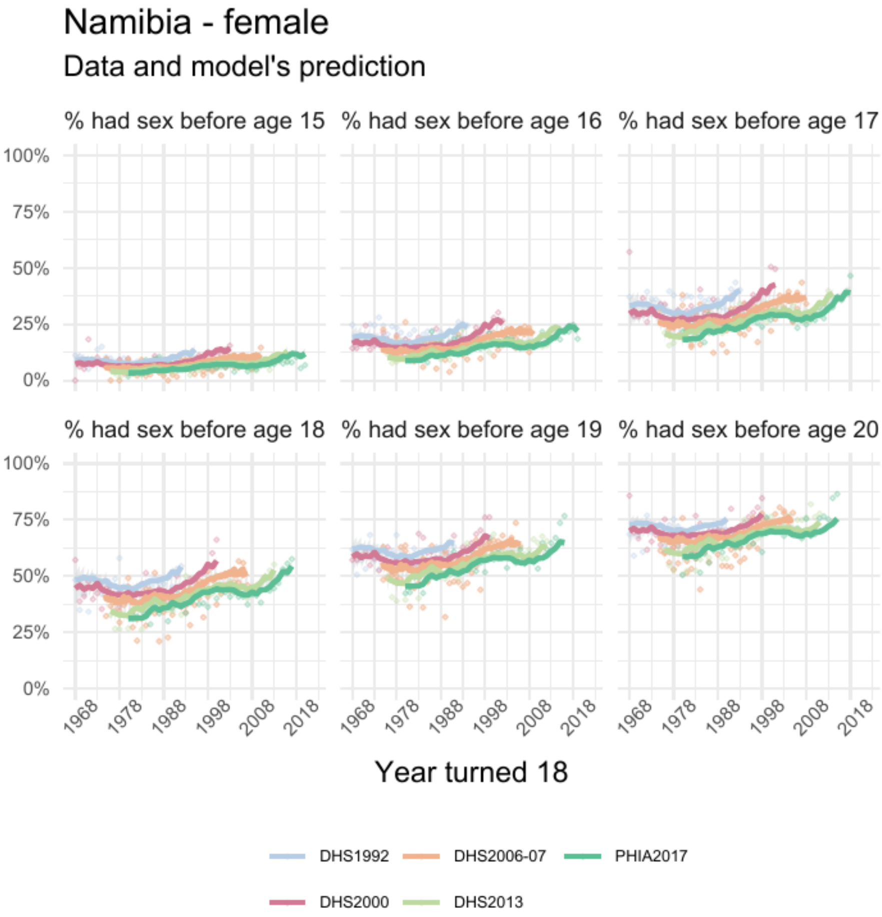


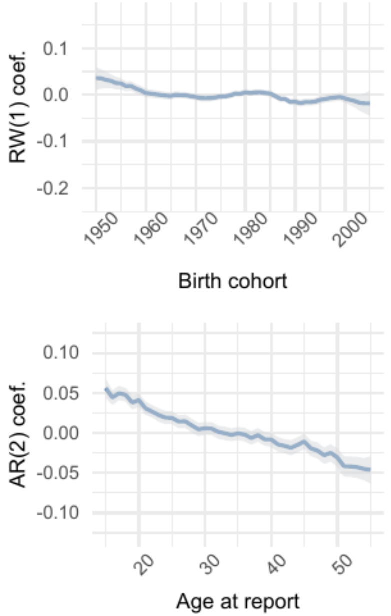


| | Parameter | Estimate |
|---|---|---|
| 1 | intercept | -0.016 [-0.027 – -0.006] |
| 2 | skew | 2.38 [2.24 – 2.53] |
| 3 | shape | 8.76 [8.61 – 8.89] |

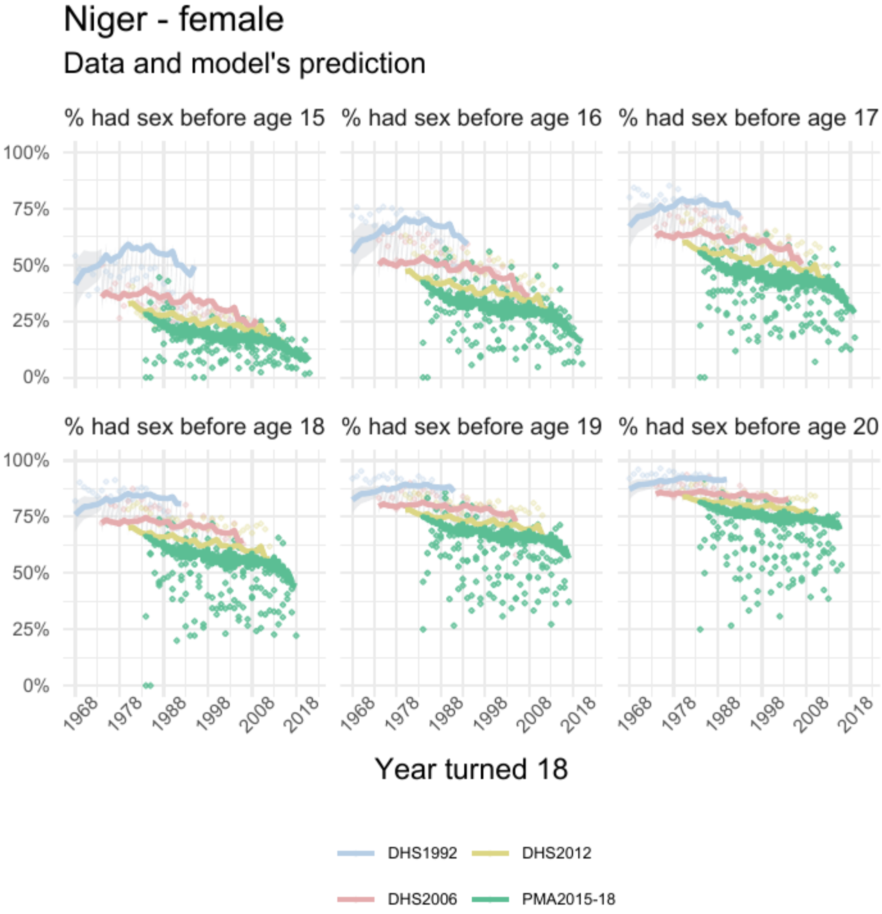


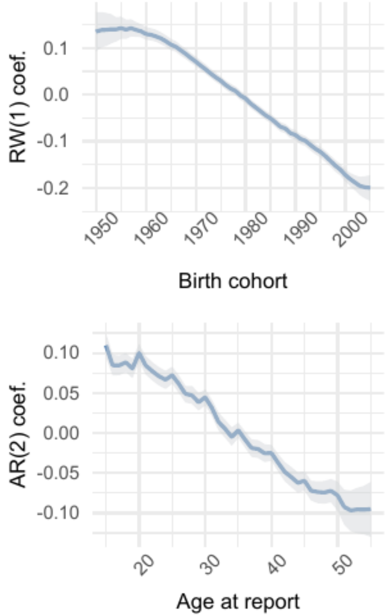


| | Parameter | Estimate |
|---|---|---|
| 1 | intercept | 0.19 [0.18 – 0.21] |
| 2 | skew | 4.19 [3.87 – 4.57] |
| 3 | shape | 6.80 [6.70 – 6.89] |

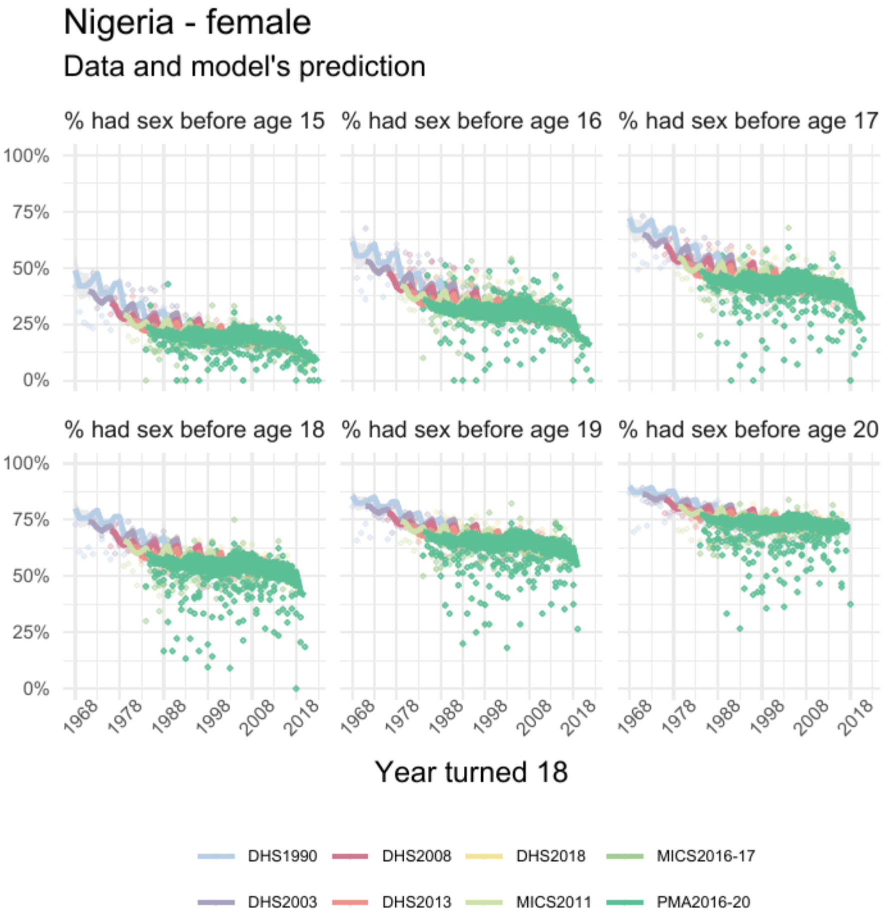


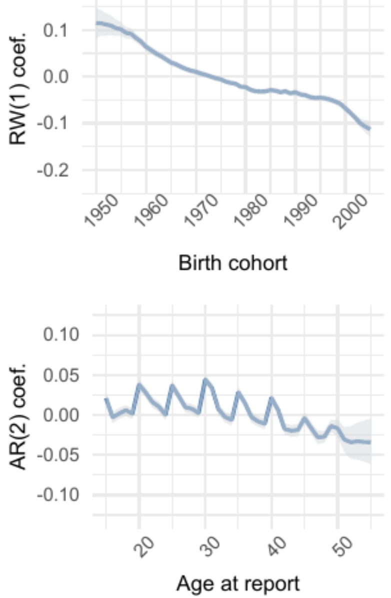


| | Parameter | Estimate |
|---|---|---|
| 1 | intercept | 0.04 [0.03 – 0.04] |
| 2 | skew | 1.87 [1.83 – 1.90] |
| 3 | shape | 7.16 [7.11 – 7.20] |

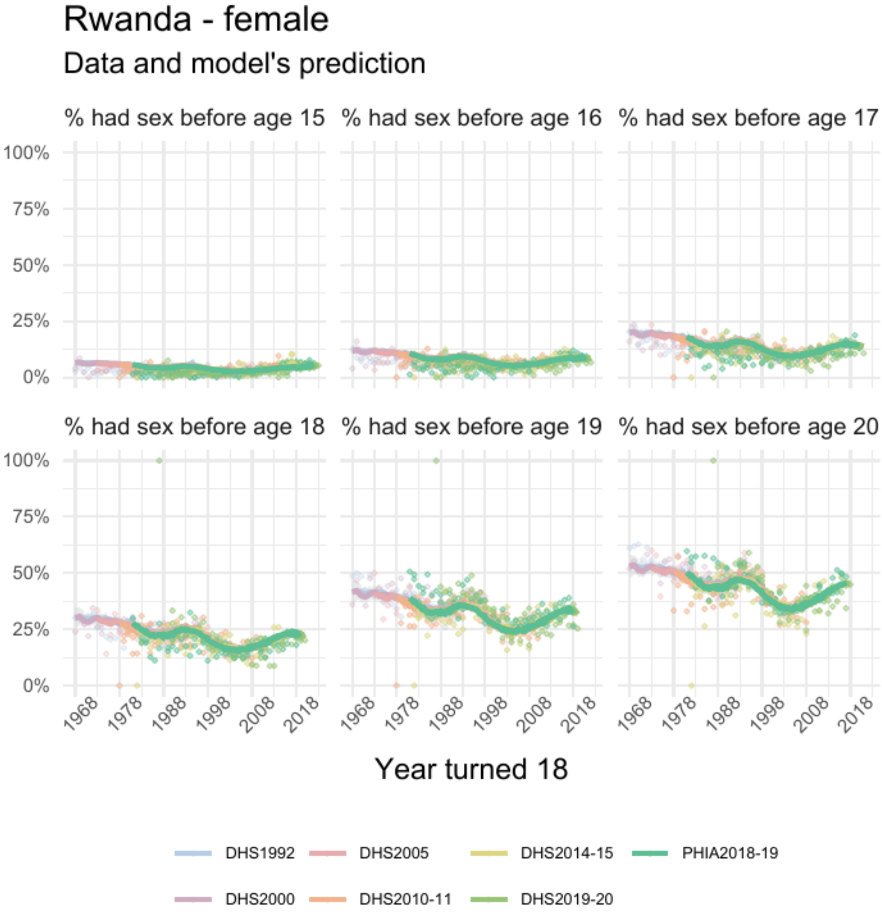


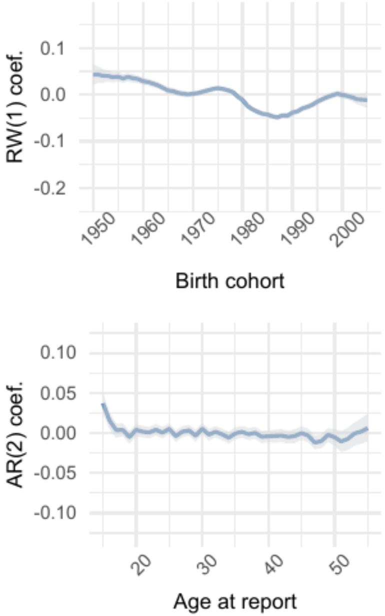


| | Parameter | Estimate |
|---|---|---|
| *1* | intercept | -0.19 [-0.19 – -0.18] |
| *2* | skew | 1.41 [1.37 – 1.45] |
| *3* | shape | 8.15 [8.06 – 8.25] |

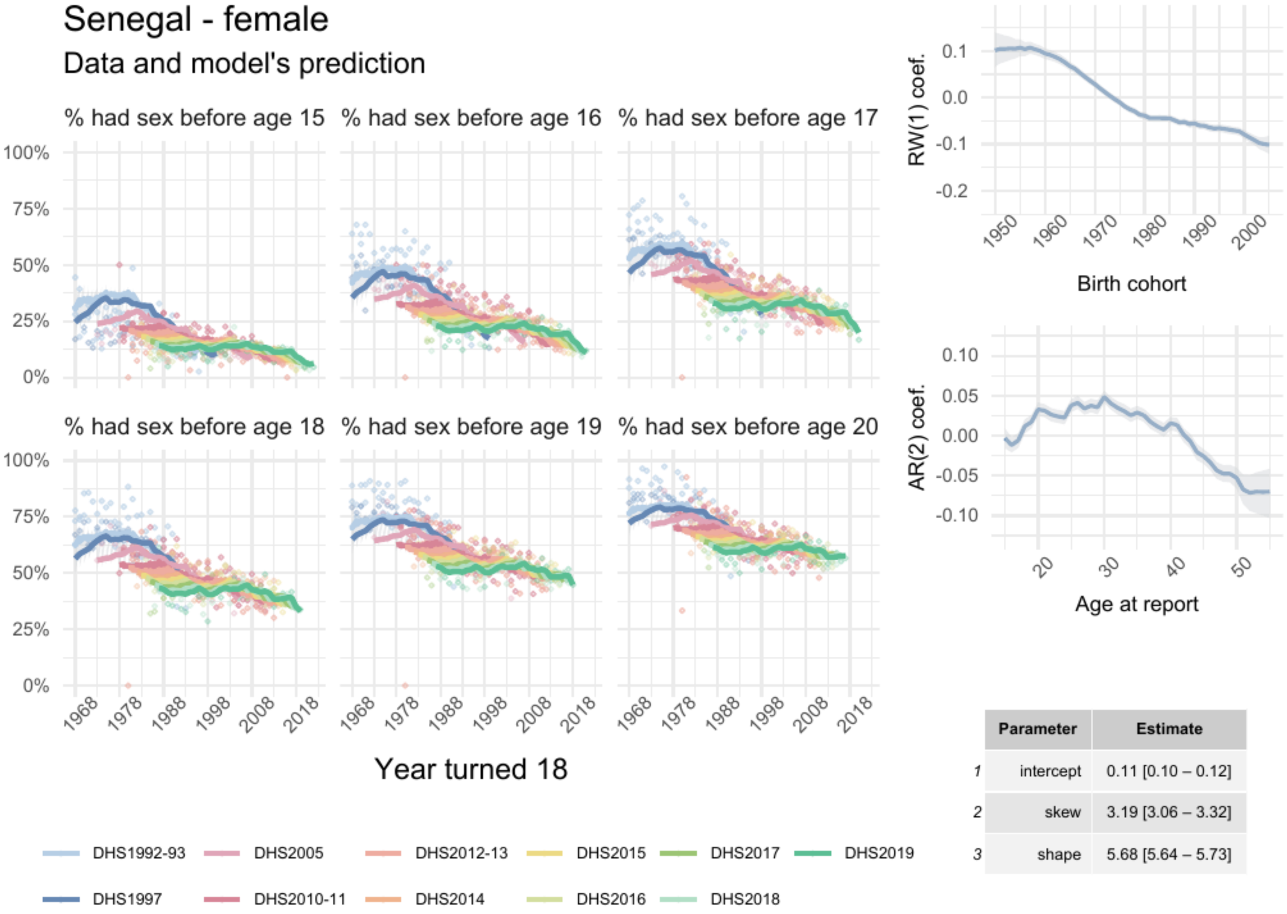


| | Parameter | Estimate |
|---|---|---|
| 1 | intercept | 0.11 [0.10 – 0.12] |
| 2 | skew | 3.19 [3.06 – 3.32] |
| 3 | shape | 5.68 [5.64 – 5.73] |

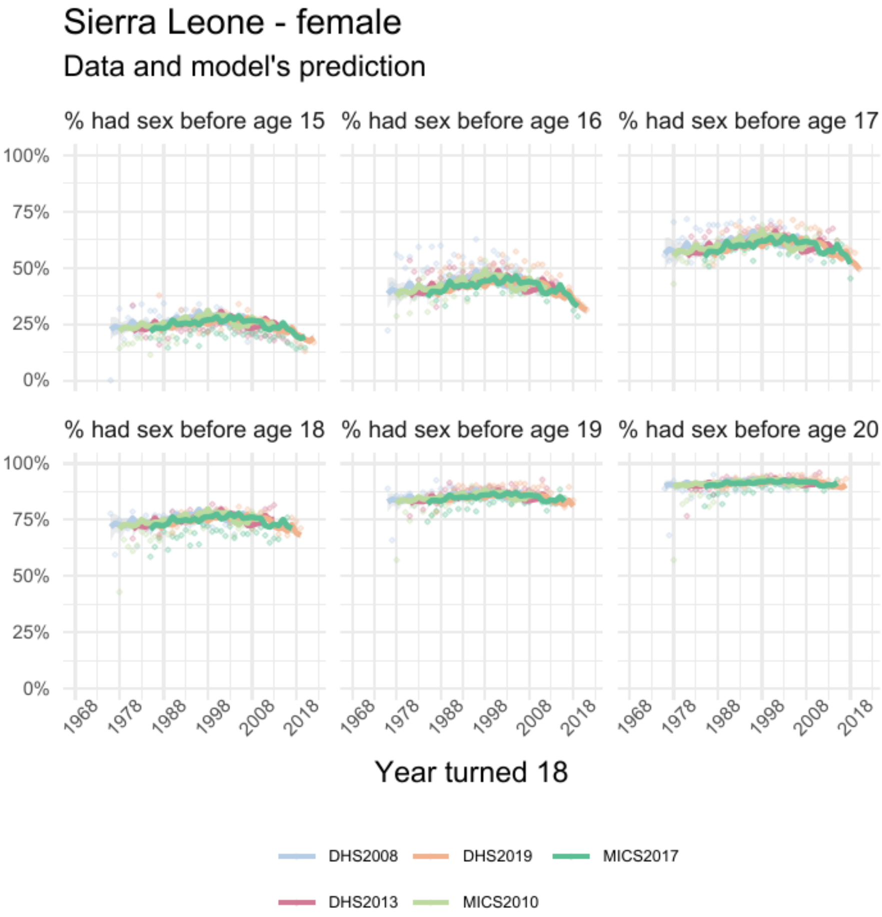


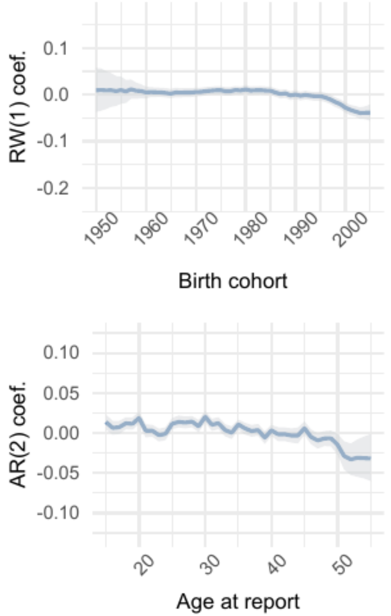


| | Parameter | Estimate |
|---|---|---|
| 1 | intercept | -0.04 [-0.04 – -0.03] |
| 2 | skew | 0.96 [0.93 – 0.98] |
| 3 | shape | 12.06 [11.93 – 12.19] |

### South Sudan - female

#### Data and model's prediction

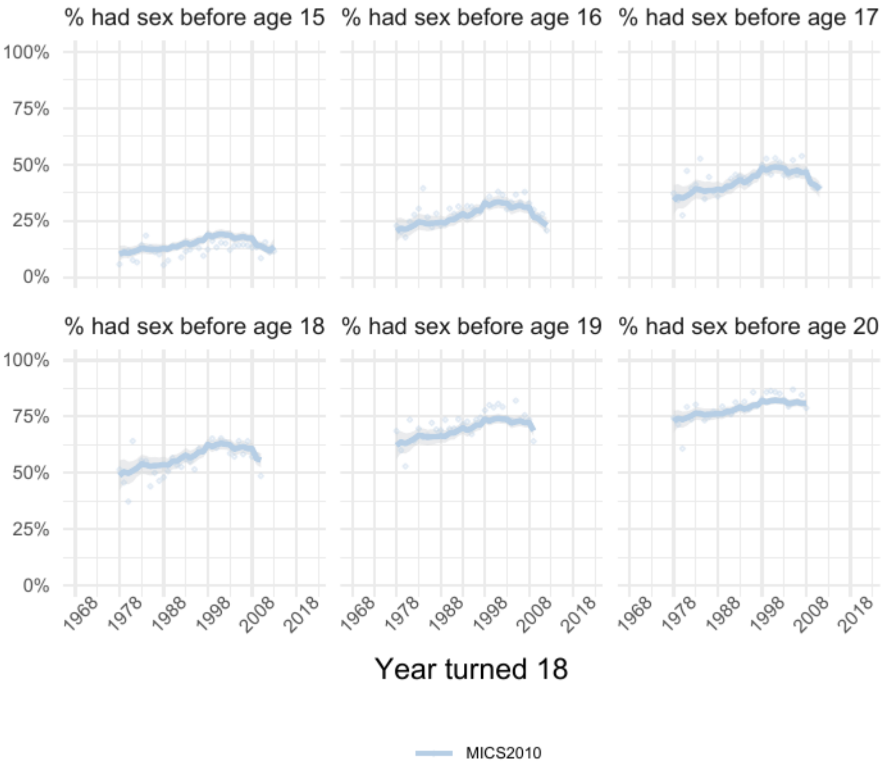


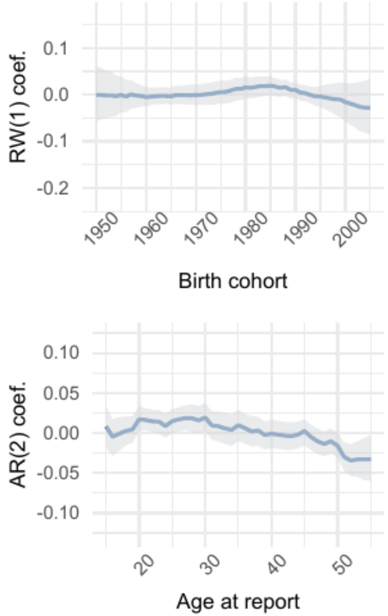


| | Parameter | Estimate |
|---|---|---|
| 1 | intercept | 0.002 [-0.020 – 0.023] |
| 2 | skew | 1.99 [1.79 – 2.20] |
| 3 | shape | 8.80 [8.53 – 9.06] |

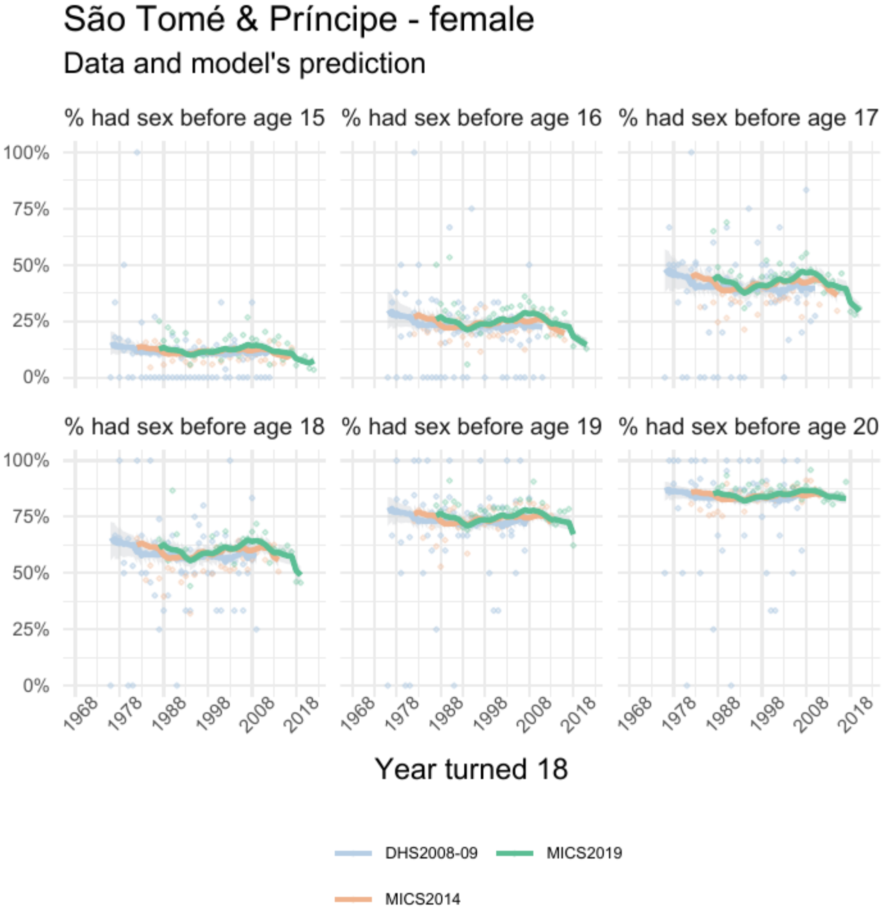


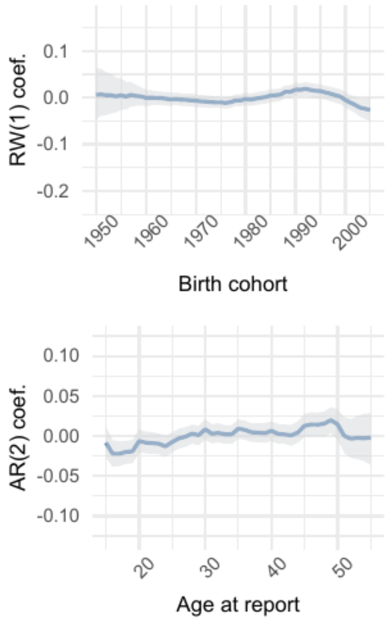


| | Parameter | Estimate |
|---|---|---|
| 1 | intercept | -0.06 [-0.07 – -0.05] |
| 2 | skew | 1.27 [1.17 – 1.38] |
| 3 | shape | 11.88 [11.47 – 12.30] |

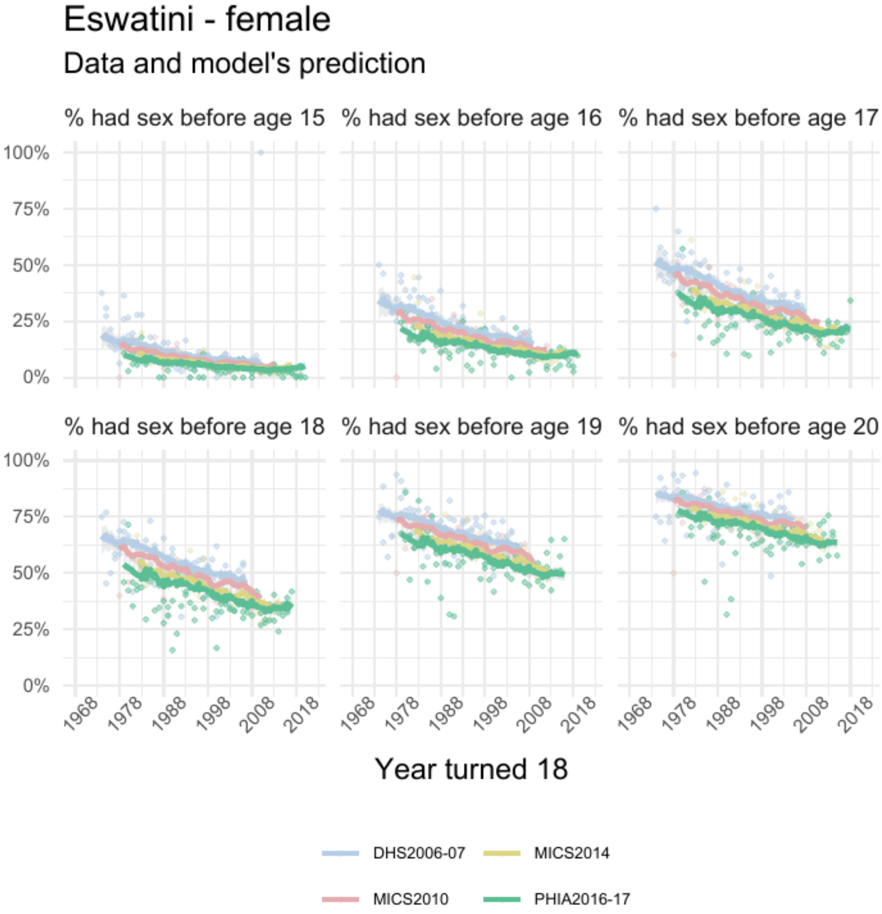


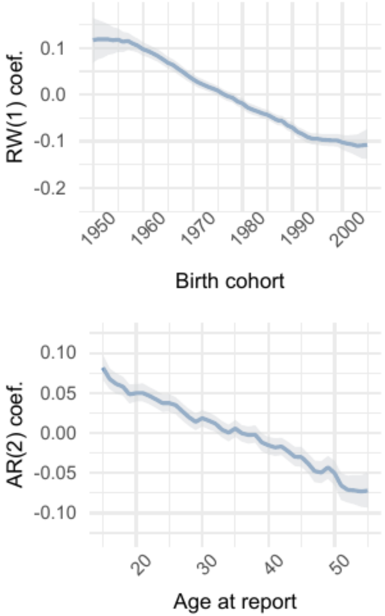


| | Parameter | Estimate |
|---|---|---|
| 1 | intercept | -1e-02 [-3e-02 – 7e-04] |
| 2 | skew | 2.27 [2.10 – 2.44] |
| 3 | shape | 9.45 [ 9.26 – 9.64] |

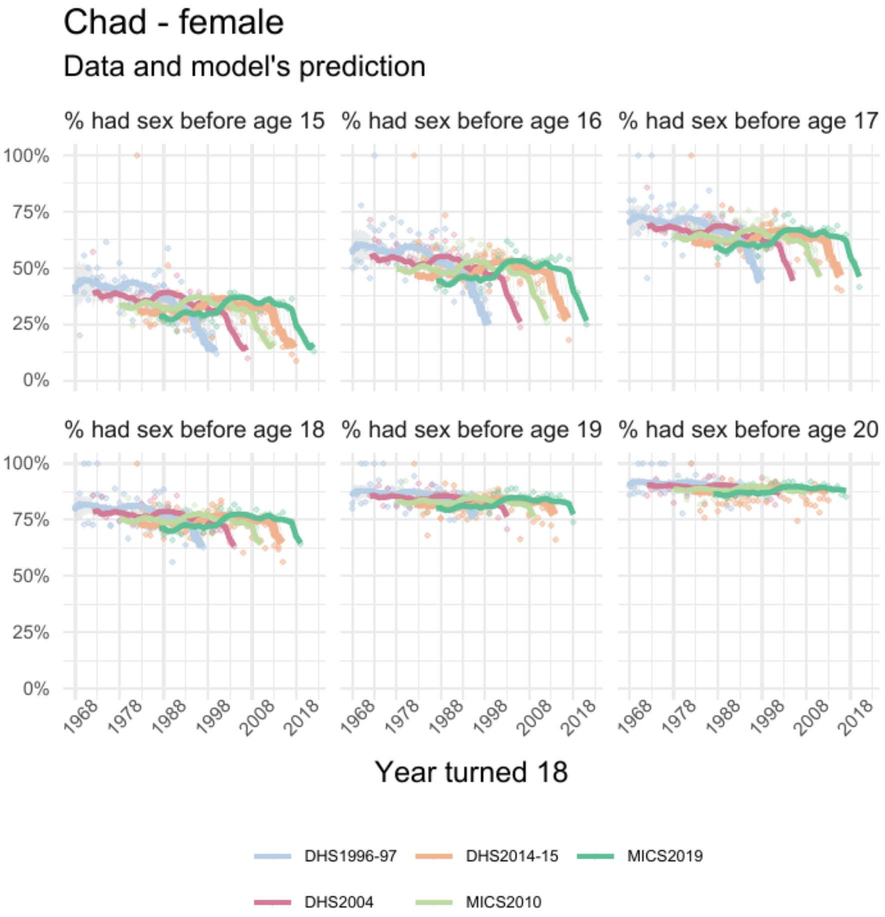


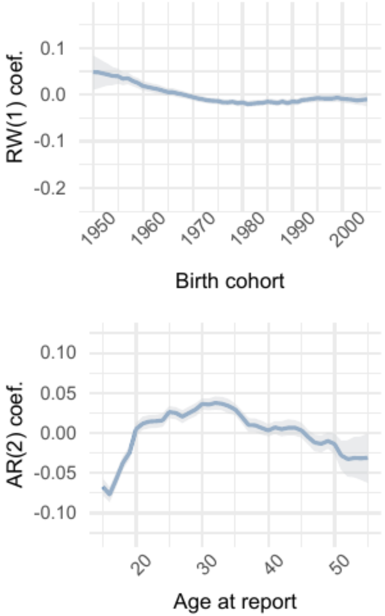


| | Parameter | Estimate |
|---|---|---|
| 1 | intercept | 0.10 [0.09 – 0.11] |
| 2 | skew | 1.97 [1.90 – 2.05] |
| 3 | shape | 8.53 [8.43 – 8.63] |

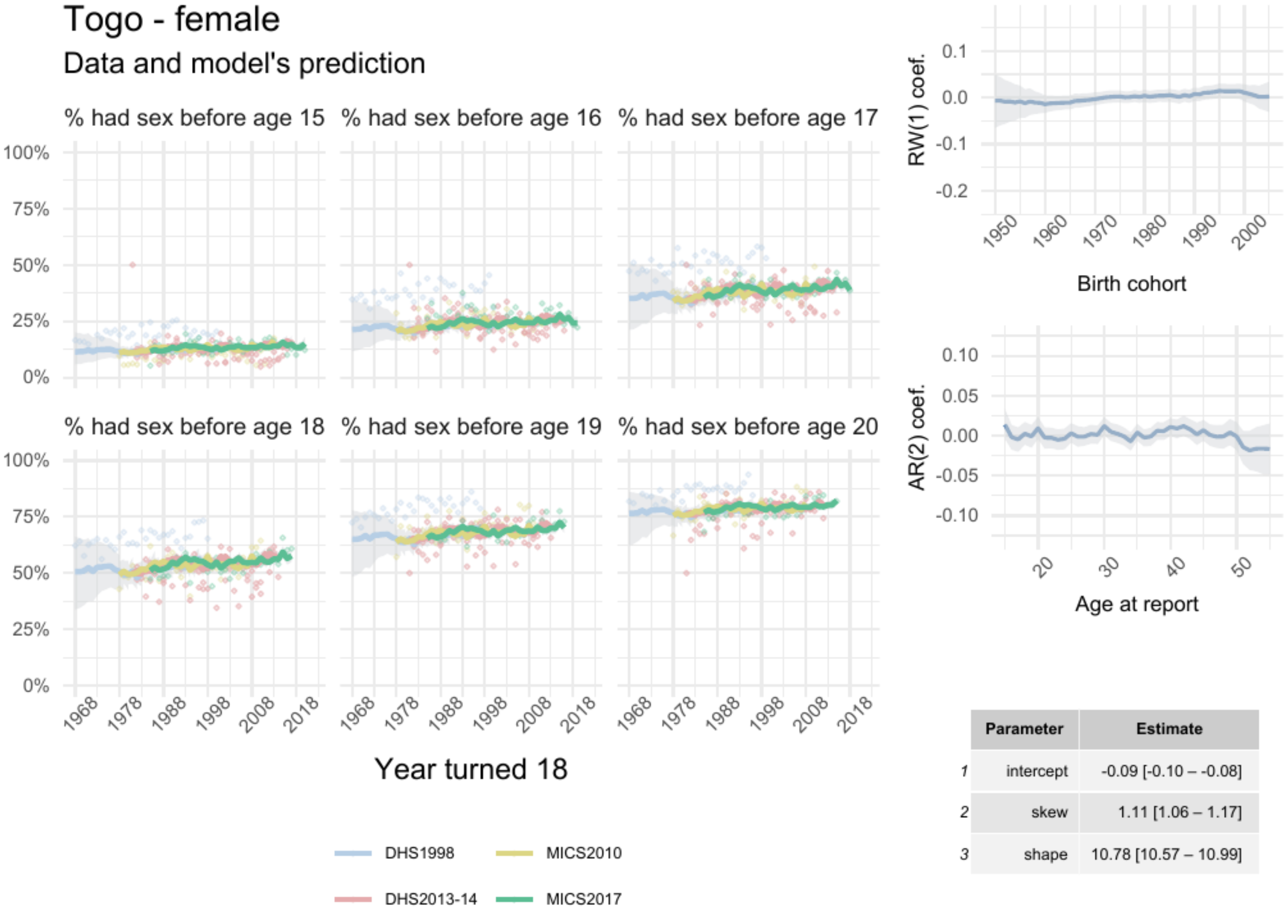


| | Parameter | Estimate |
|---|---|---|
| 1 | intercept | -0.09 [-0.10 – -0.08] |
| 2 | skew | 1.11 [1.06 – 1.17] |
| 3 | shape | 10.78 [10.57 – 10.99] |

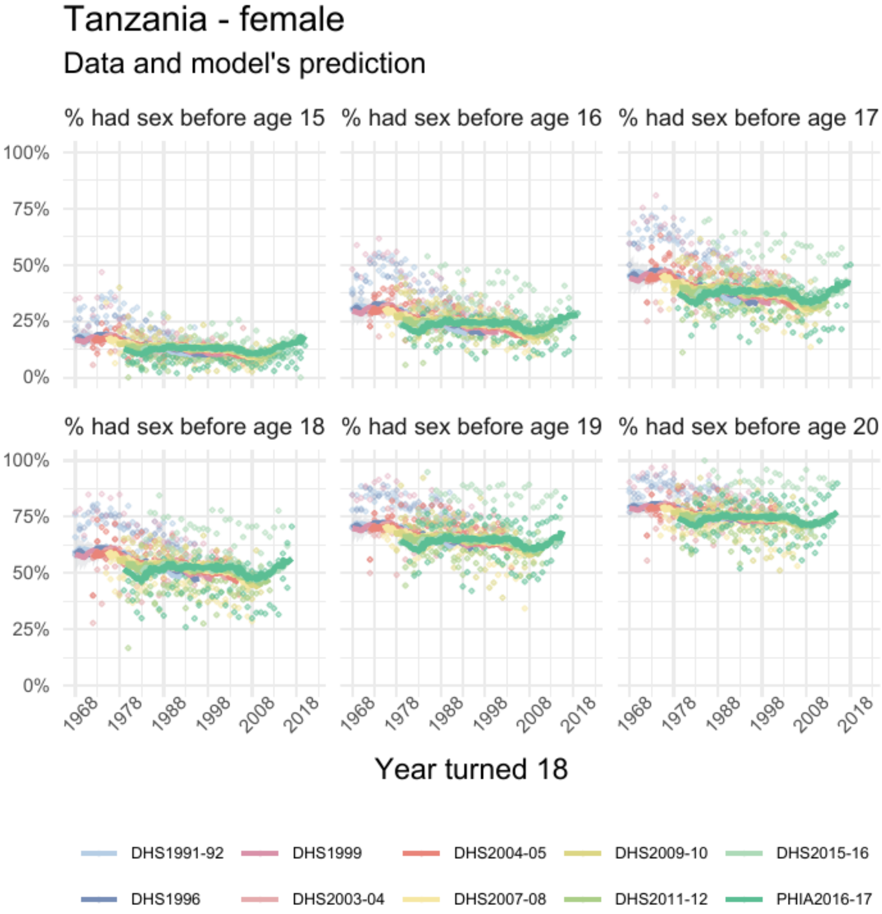


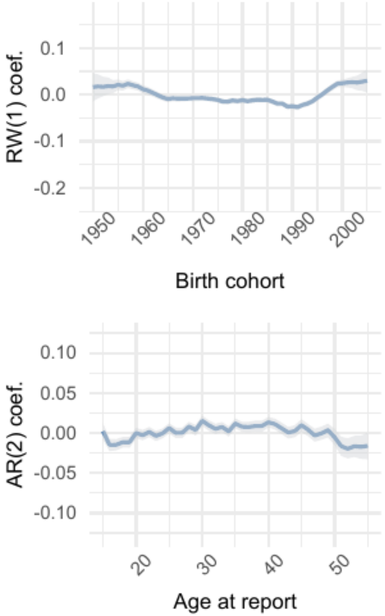


| | Parameter | Estimate |
|---|---|---|
| 1 | intercept | -0.006 [-0.013 – 0.001] |
| 2 | skew | 1.87 [1.81 – 1.92] |
| 3 | shape | 8.57 [8.50 – 8.65] |

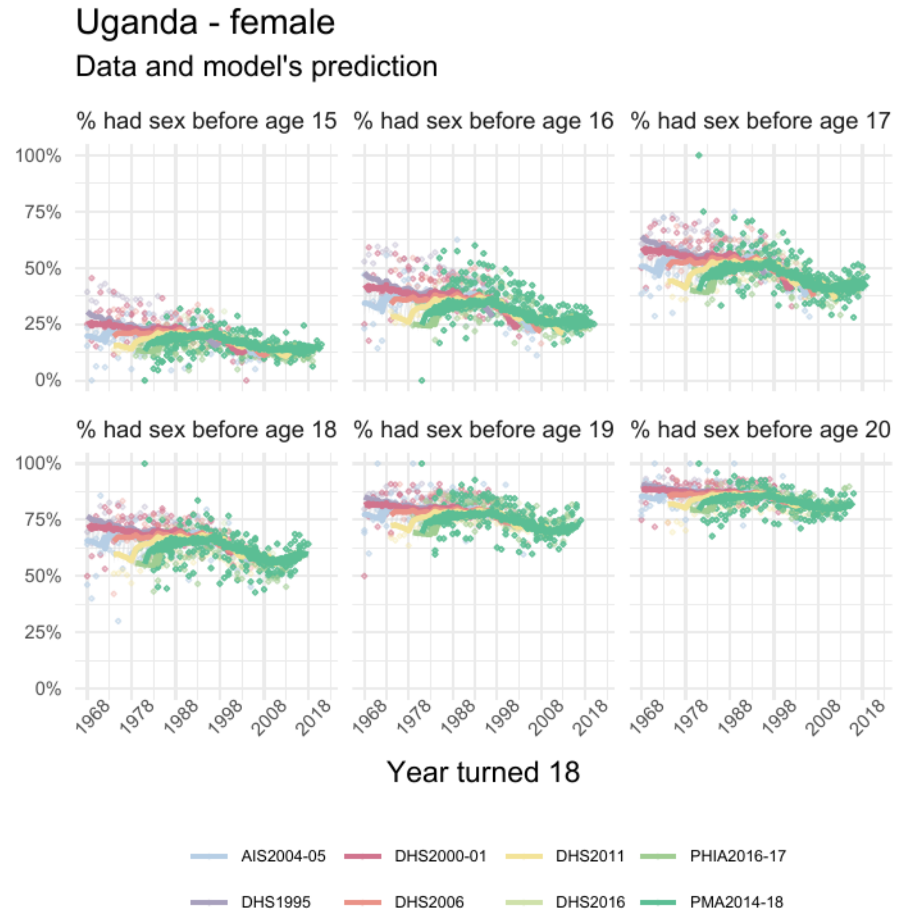


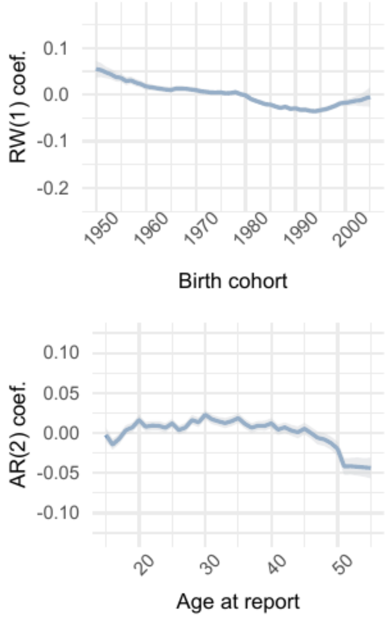


| | Parameter | Estimate |
|---|---|---|
| 1 | intercept | -0.03 [-0.04 – -0.02] |
| 2 | skew | 1.32 [1.29 – 1.35] |
| 3 | shape | 10.25 [10.15 – 10.34] |

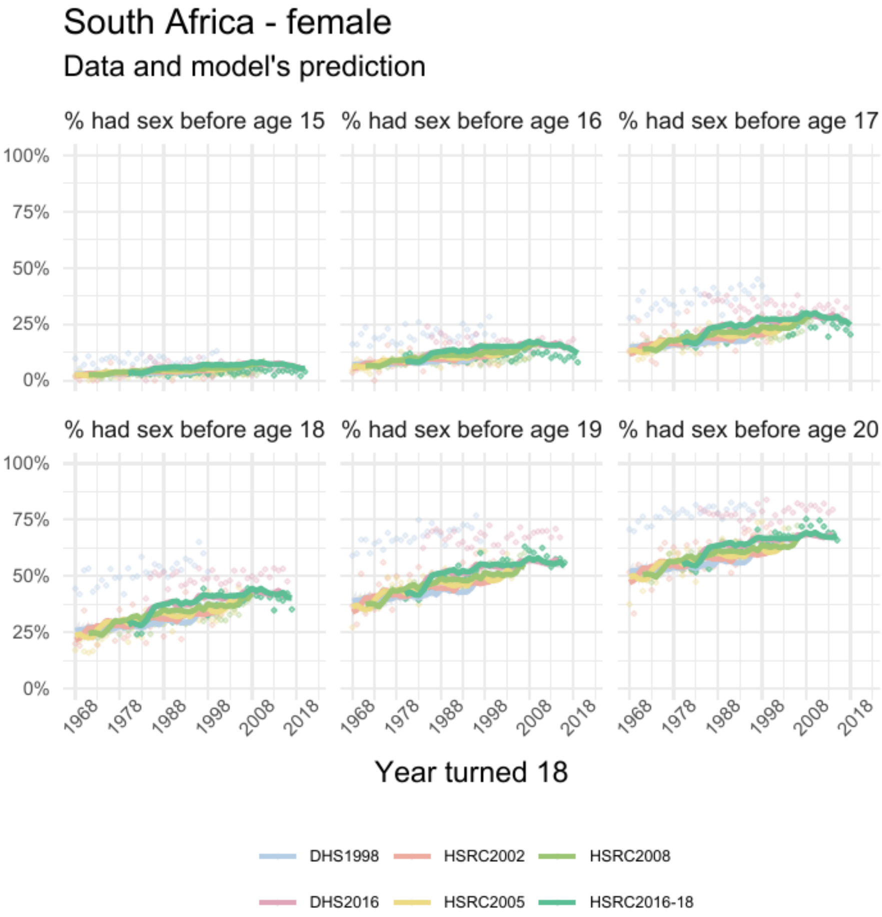


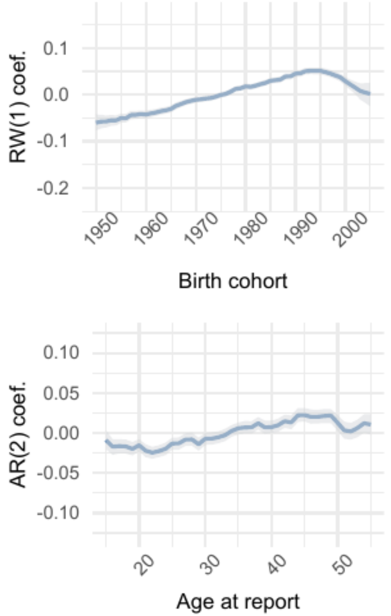


| | Parameter | Estimate |
|---|---|---|
| 1 | intercept | -0.05 [-0.06 – -0.05] |
| 2 | skew | 2.24 [2.17 – 2.32] |
| 3 | shape | 8.52 [8.42 – 8.61] |

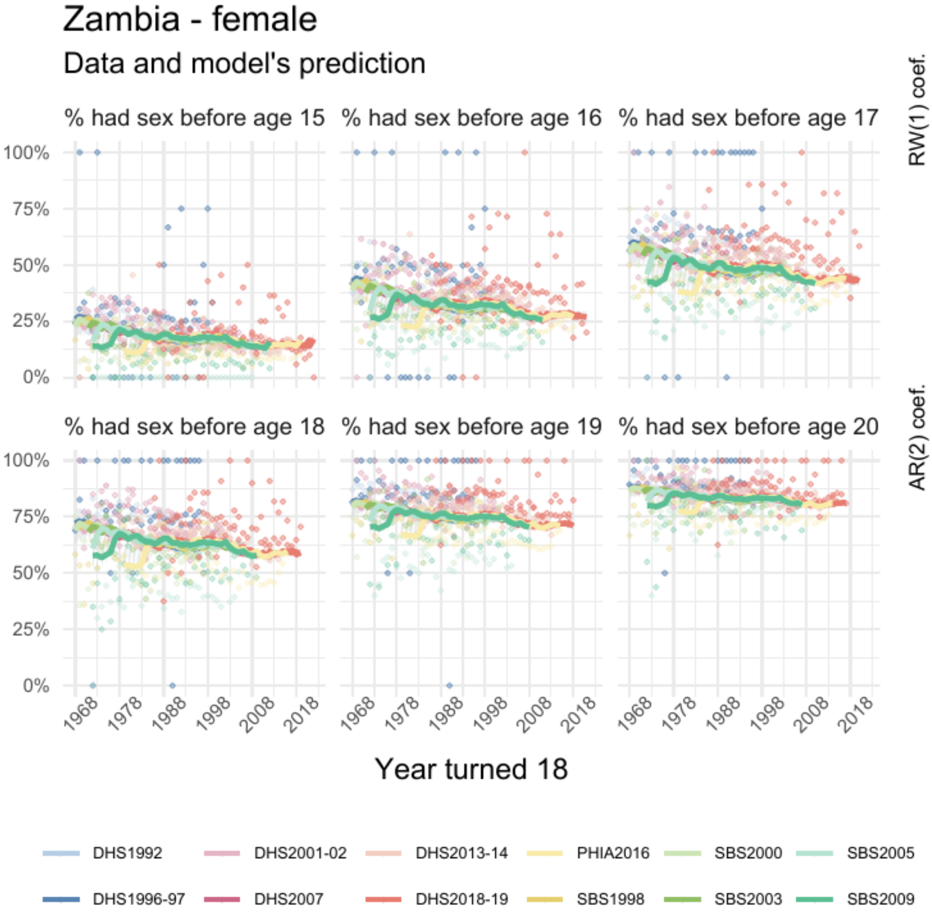


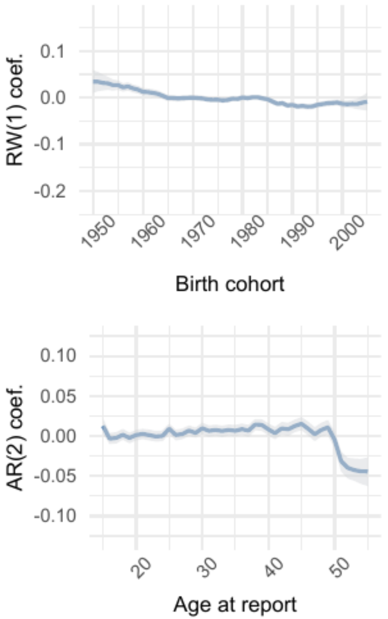


| | Parameter | Estimate |
|---|---|---|
| 1 | intercept | 0.02 [0.01 – 0.02] |
| 2 | skew | 1.86 [1.79 – 1.93] |
| 3 | shape | 9.38 [9.27 – 9.49] |

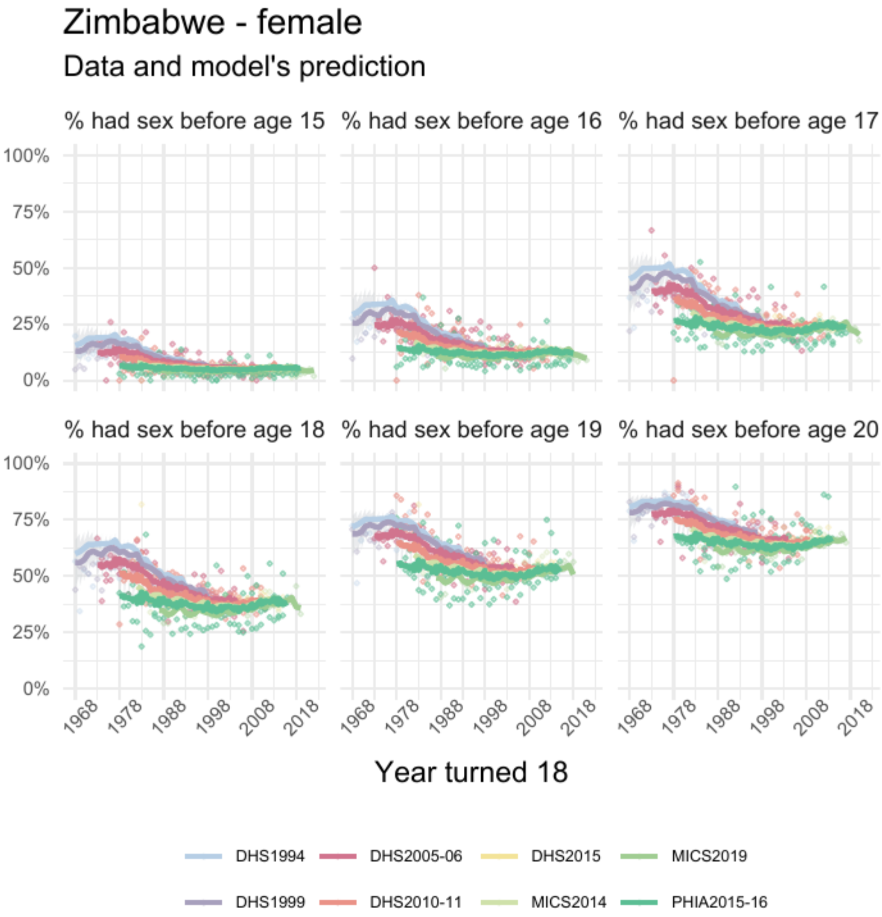


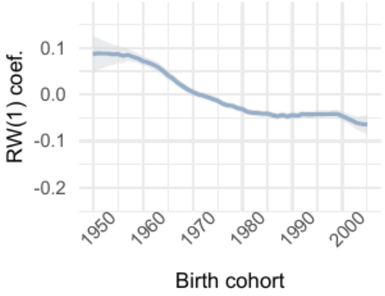


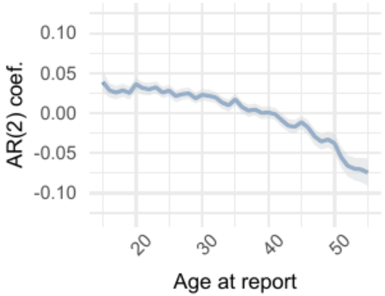


| | Parameter | Estimate |
|---|---|---|
| 1 | intercept | -0.04 [-0.05 – -0.03] |
| 2 | skew | 2.10 [2.01 – 2.19] |
| 3 | shape | 8.99 [8.88 – 9.10] |
