## Supplementary material for "Trends and country-level variation in age at first sex in sub-Saharan Africa among birth cohorts entering adulthood between 1985 and 2020": Figure_S3_male.pdf

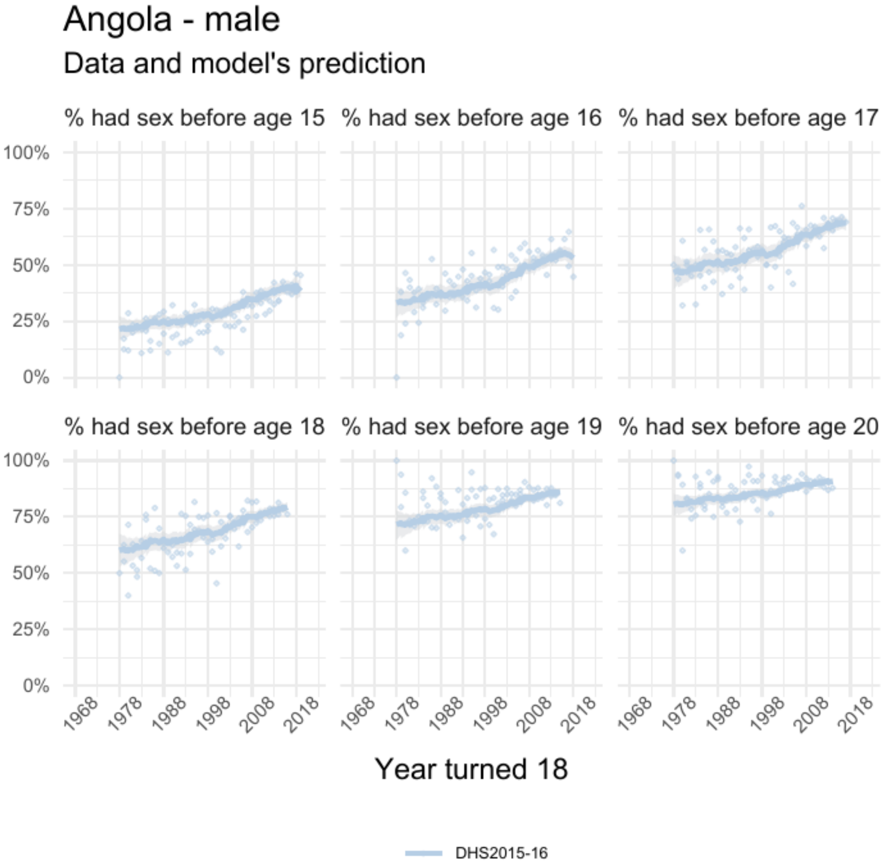


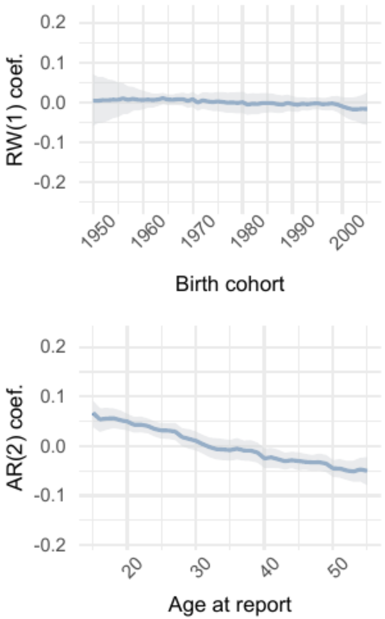


| | Parameter | Estimate |
|---|---|---|
| *1* | intercept | 0.08 [0.07 – 0.10] |
| *2* | skew | 0.89 [0.81 – 0.97] |
| *3* | shape | 9.81 [ 9.40 – 10.28] |

### Burundi - male

#### Data and model's prediction

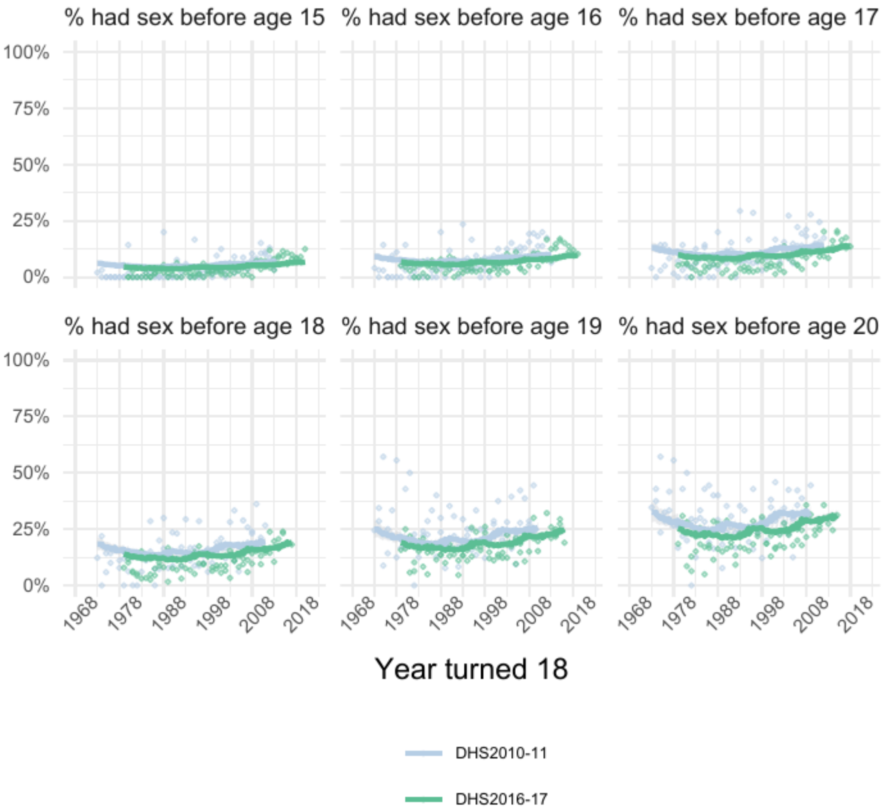


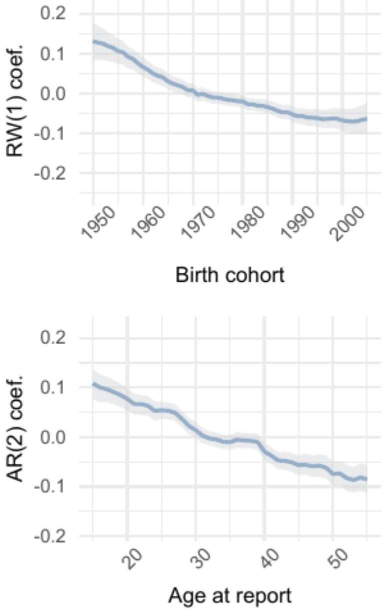


| | Parameter | Estimate |
|---|---|---|
| 1 | intercept | -0.26 [-0.28 – -0.25] |
| 2 | skew | 0.72 [0.68 – 0.76] |
| 3 | shape | 8.84 [8.58 – 9.09] |

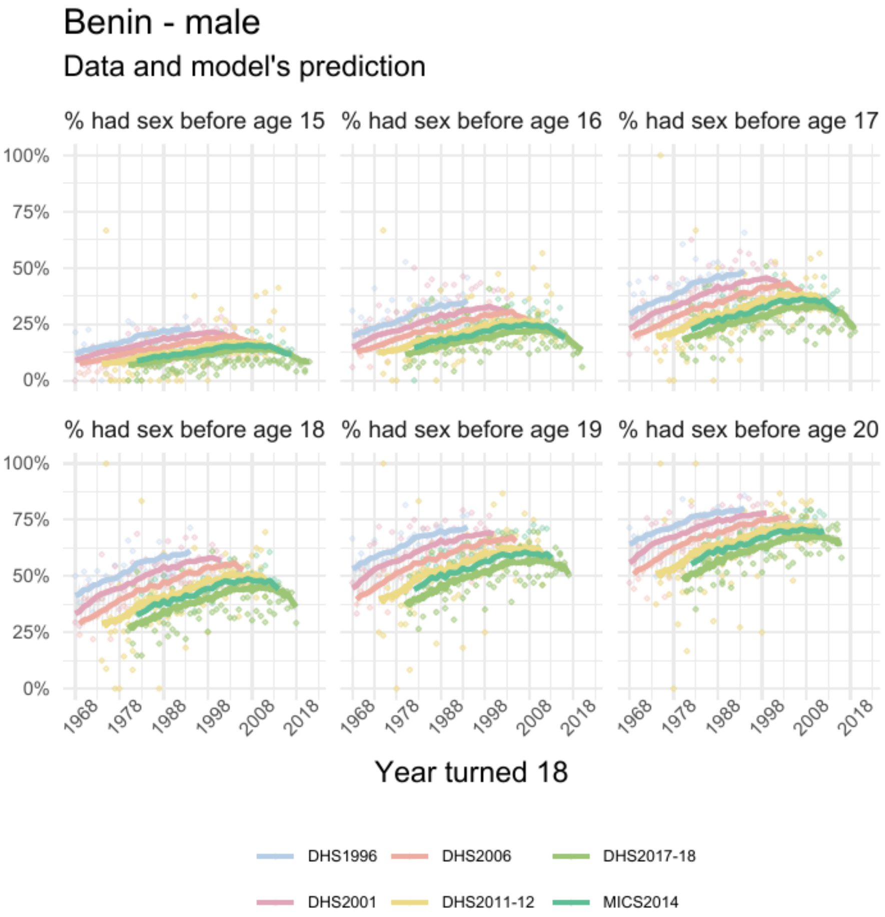


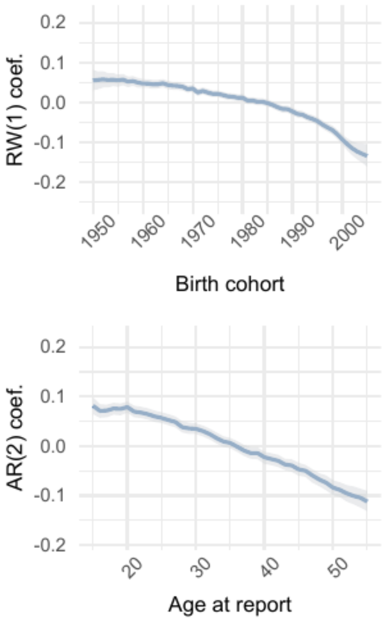


| | Parameter | Estimate |
|---|---|---|
| *1* | intercept | -0.03 [-0.04 – -0.02] |
| *2* | skew | 1.00 [0.95 – 1.04] |
| *3* | shape | 8.91 [8.73 – 9.11] |

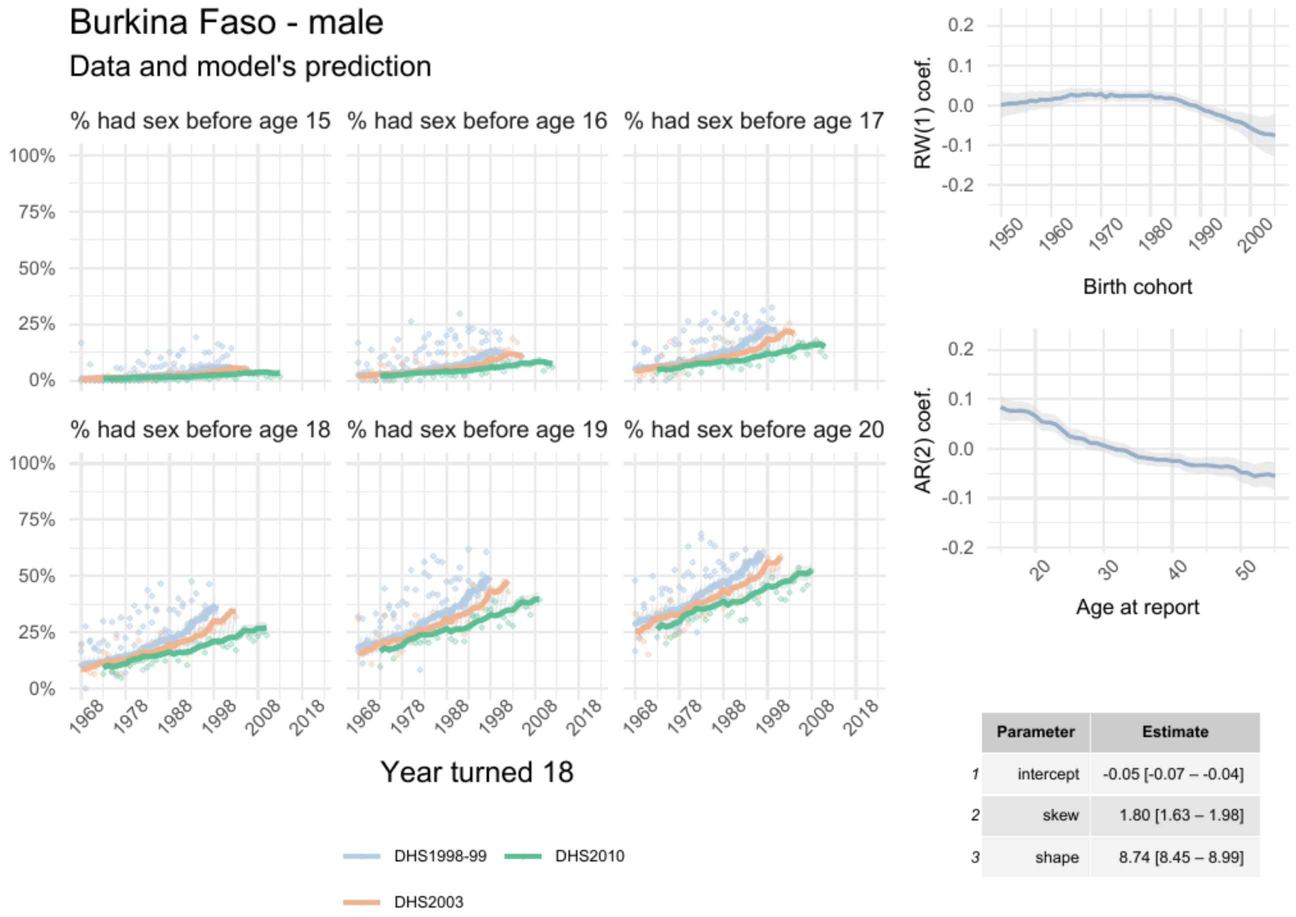


| | Parameter | Estimate |
|---|---|---|
| 1 | intercept | -0.05 [-0.07 – -0.04] |
| 2 | skew | 1.80 [1.63 – 1.98] |
| 3 | shape | 8.74 [8.45 – 8.99] |

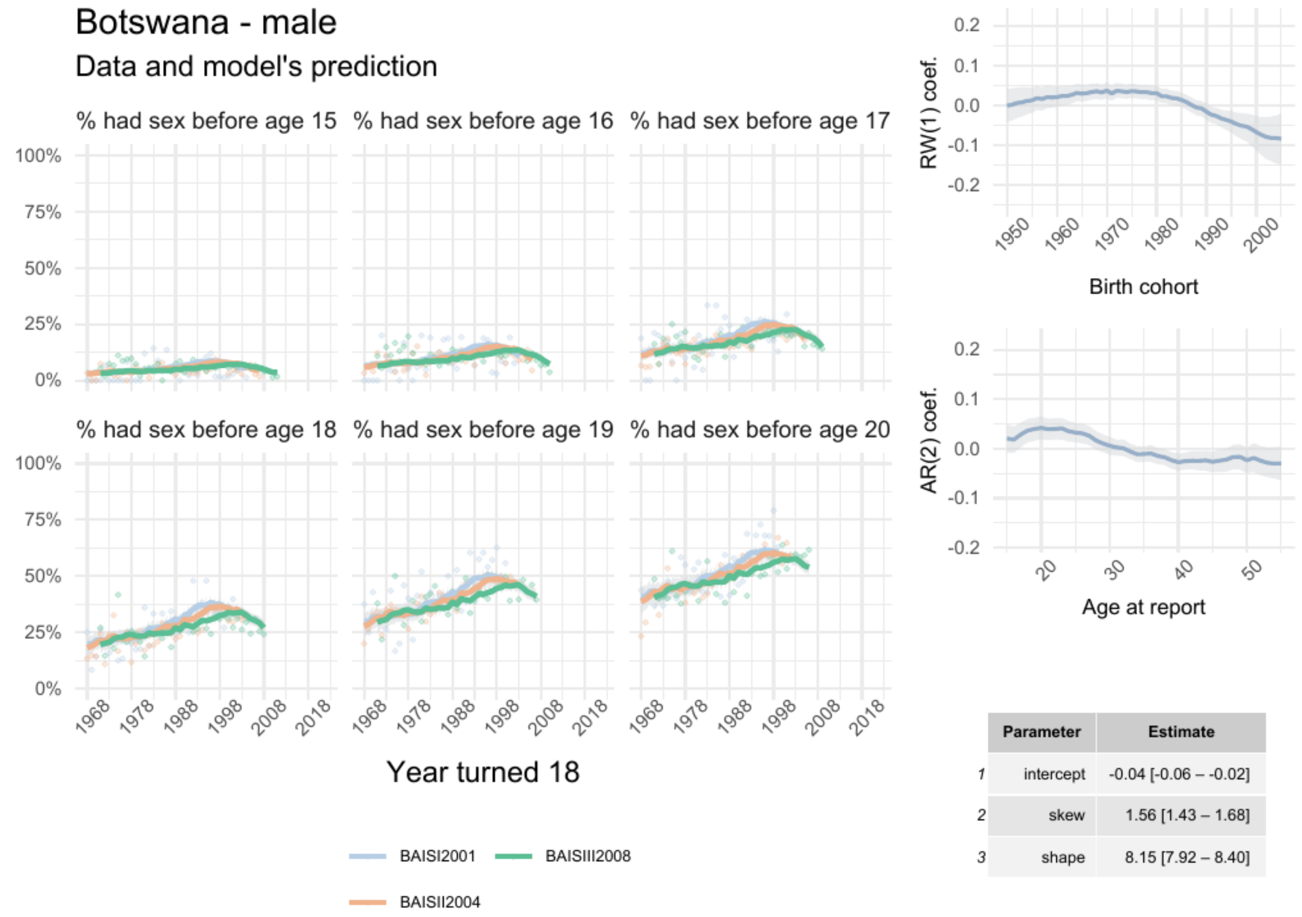


| | Parameter | Estimate |
|---|---|---|
| 1 | intercept | -0.04 [-0.06 – -0.02] |
| 2 | skew | 1.56 [1.43 – 1.68] |
| 3 | shape | 8.15 [7.92 – 8.40] |

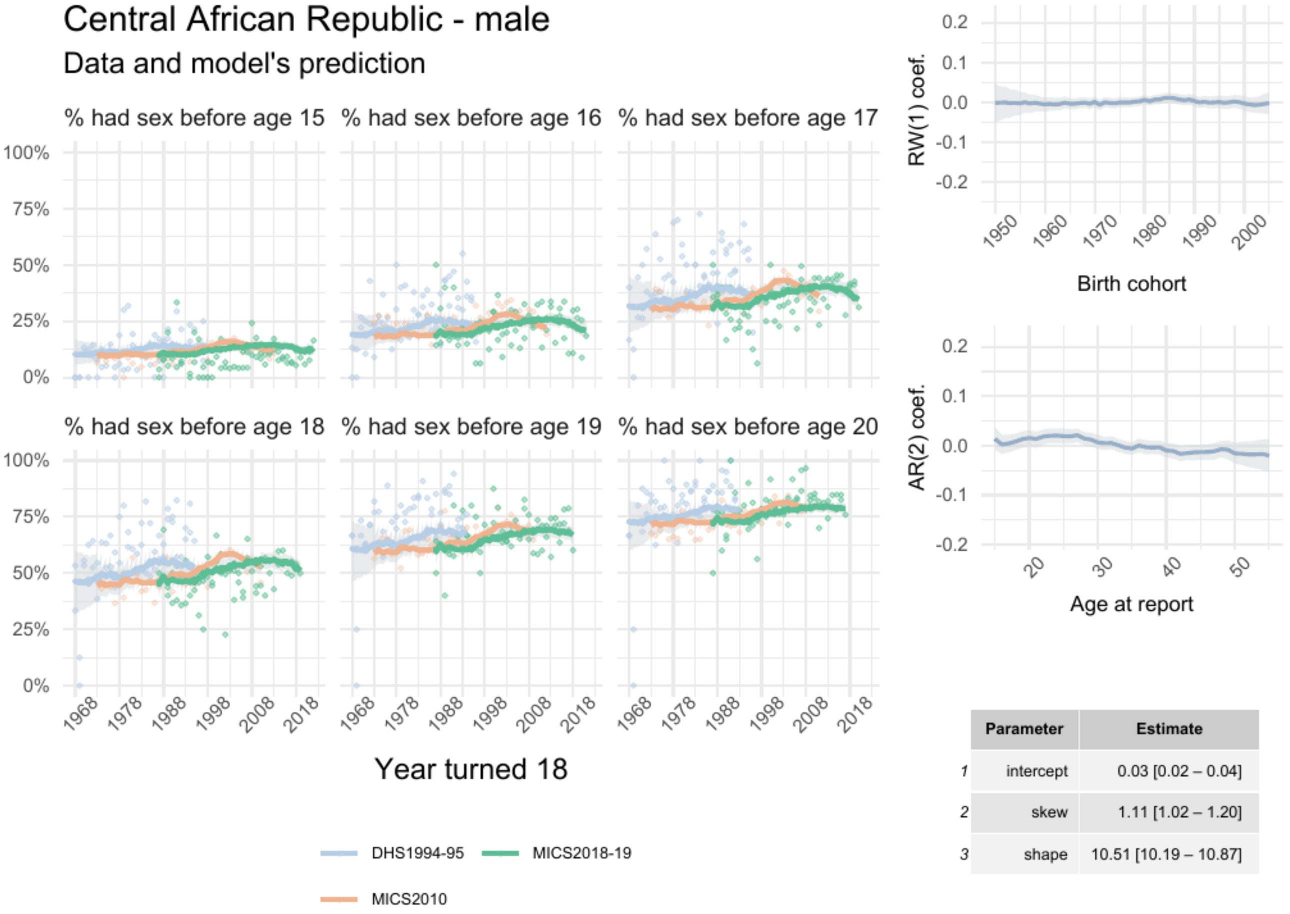


| | Parameter | Estimate |
|---|---|---|
| 1 | intercept | 0.03 [0.02 – 0.04] |
| 2 | skew | 1.11 [1.02 – 1.20] |
| 3 | shape | 10.51 [10.19 – 10.87] |

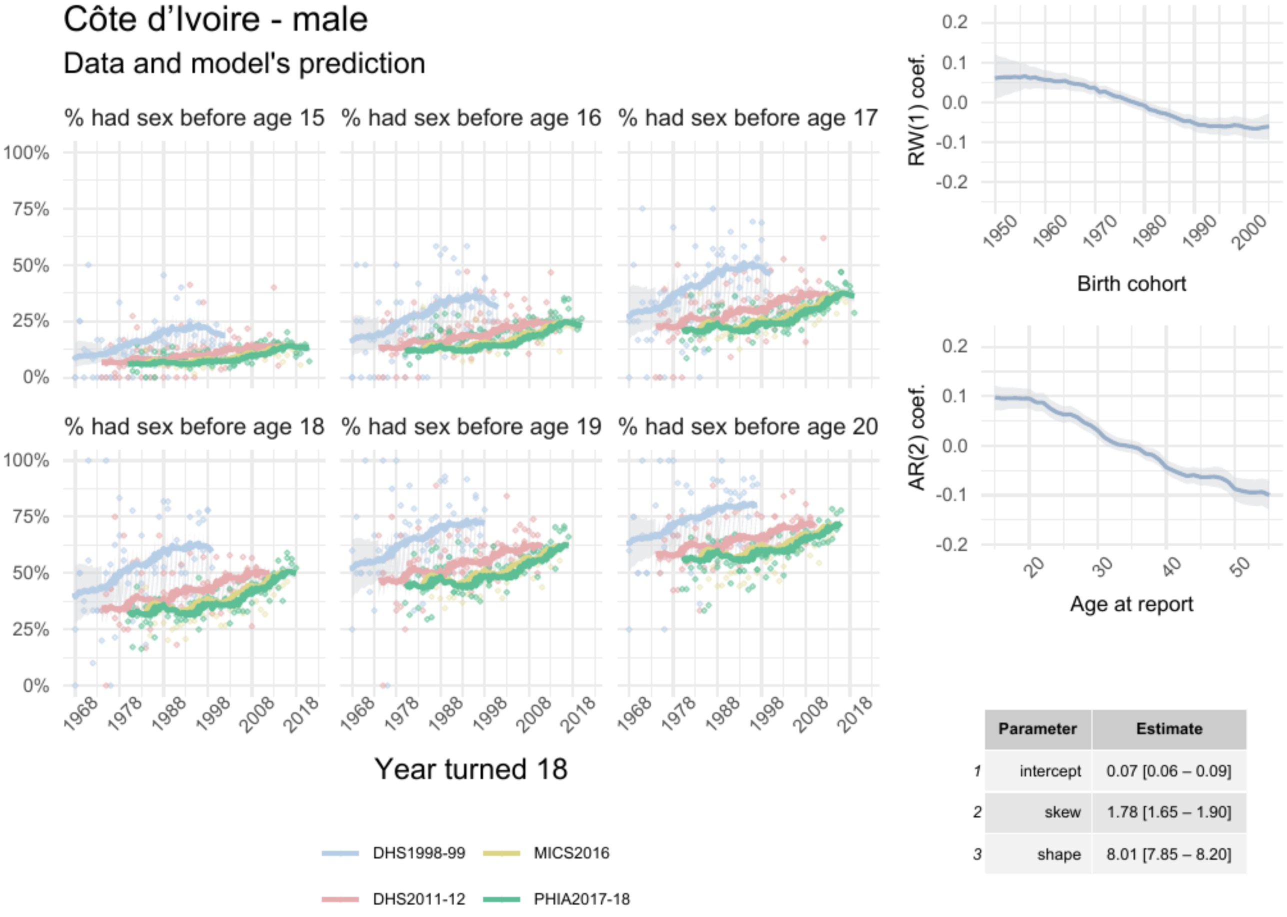


| | Parameter | Estimate |
|---|---|---|
| *1* | intercept | 0.07 [0.06 – 0.09] |
| *2* | skew | 1.78 [1.65 – 1.90] |
| *3* | shape | 8.01 [7.85 – 8.20] |

Cameroon - male

Data and model's prediction

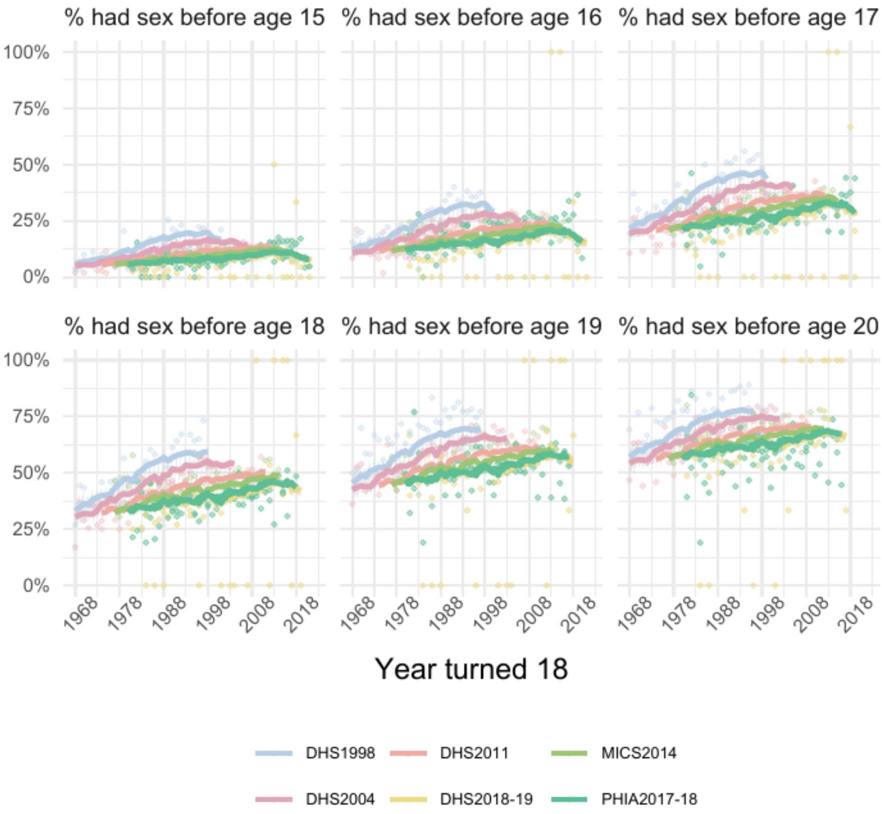


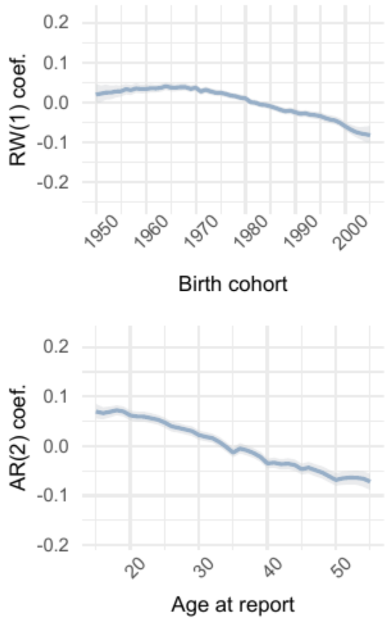


| | Parameter | Estimate |
|---|---|---|
| 1 | intercept | 0.10 [0.09 – 0.11] |
| 2 | skew | 2.13 [2.01 – 2.27] |
| 3 | shape | 7.72 [7.60 – 7.85] |

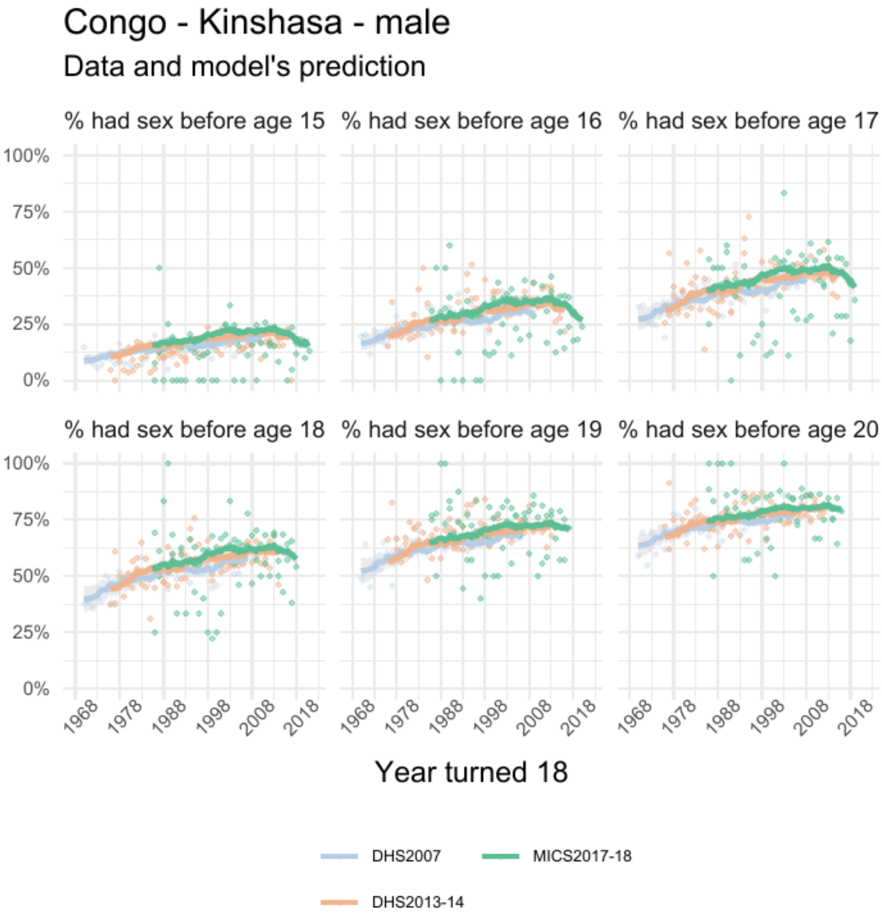


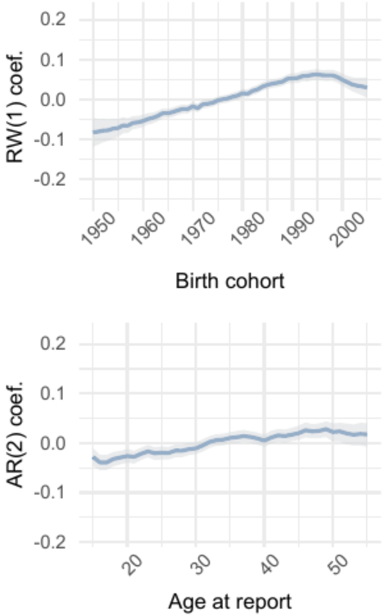


| | Parameter | Estimate |
|---|---|---|
| 1 | intercept | 0.11 [0.09 – 0.12] |
| 2 | skew | 1.62 [1.53 – 1.73] |
| 3 | shape | 8.24 [8.06 – 8.42] |

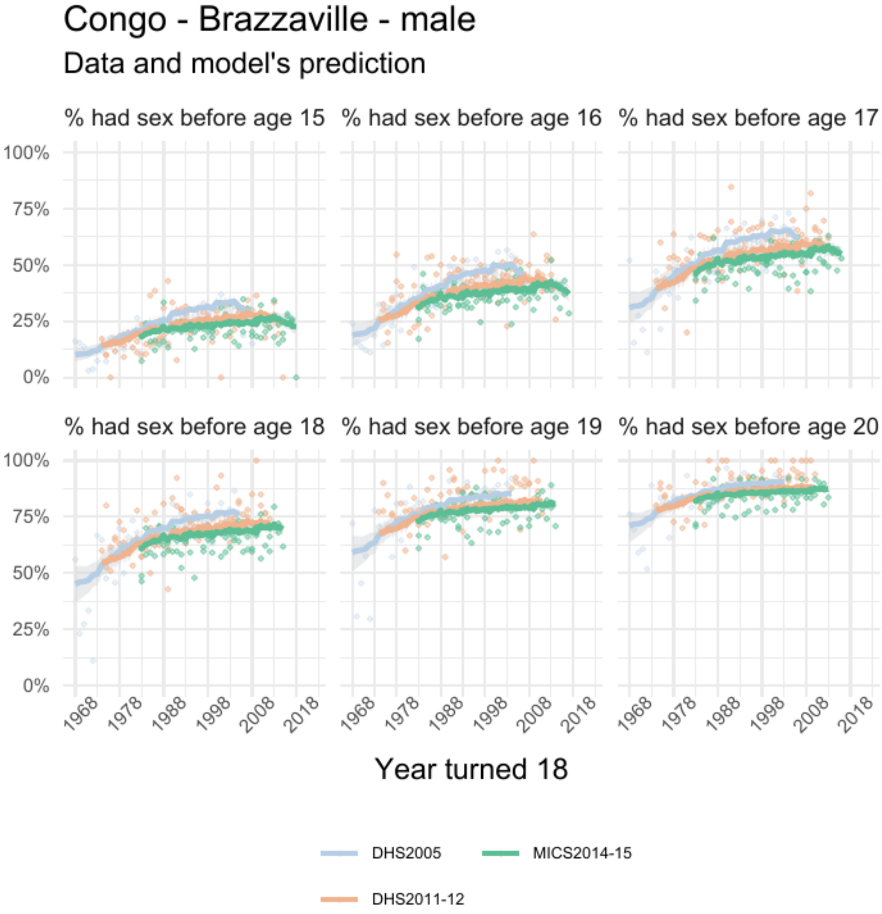


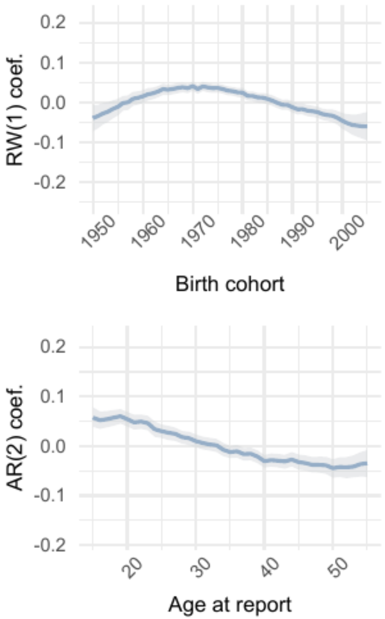


| | Parameter | Estimate |
|---|---|---|
| 1 | intercept | 0.09 [0.08 – 0.11] |
| 2 | skew | 1.21 [1.13 – 1.29] |
| 3 | shape | 9.93 [ 9.68 – 10.21] |

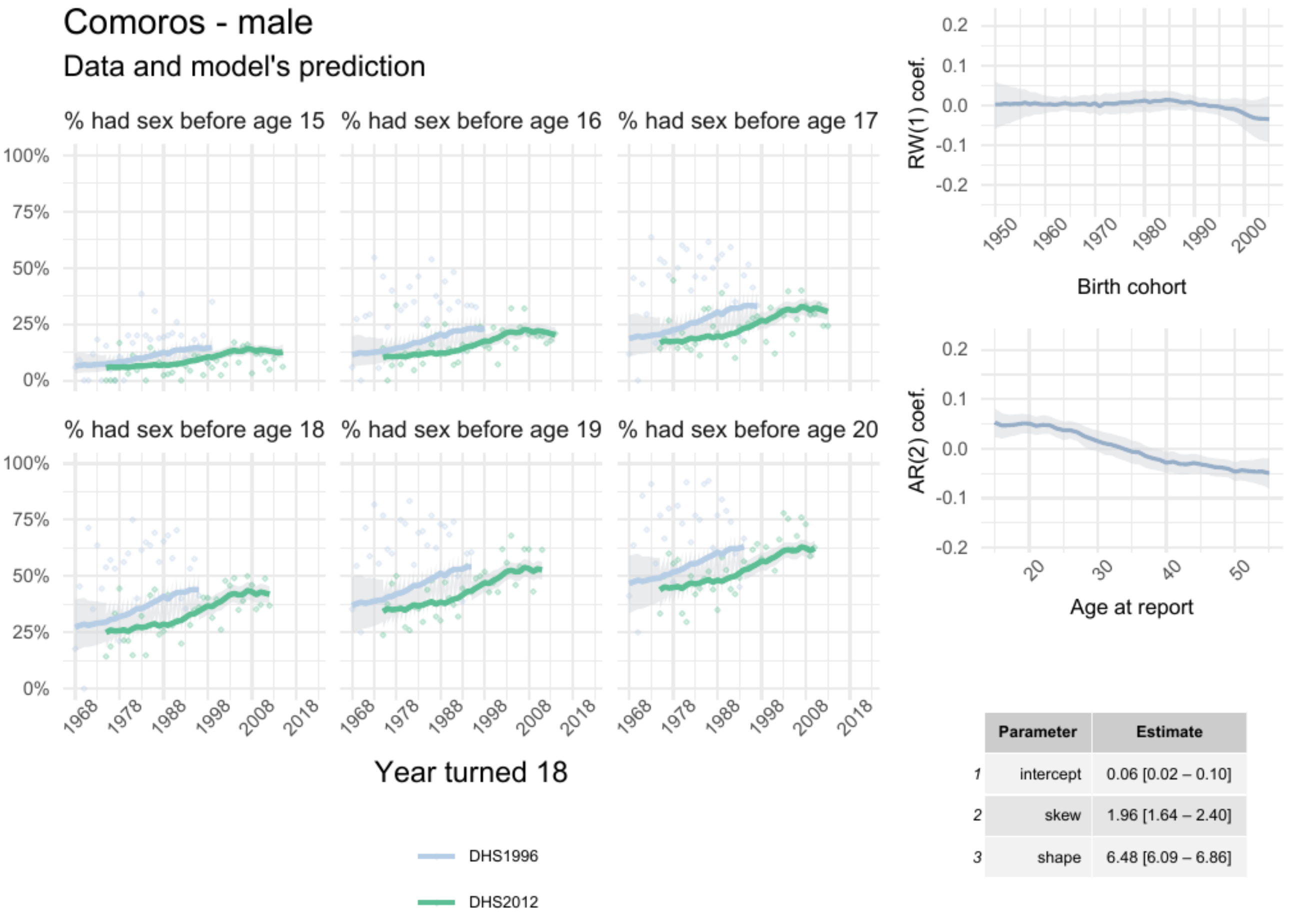


| | Parameter | Estimate |
|---|---|---|
| 1 | intercept | 0.06 [0.02 – 0.10] |
| 2 | skew | 1.96 [1.64 – 2.40] |
| 3 | shape | 6.48 [6.09 – 6.86] |

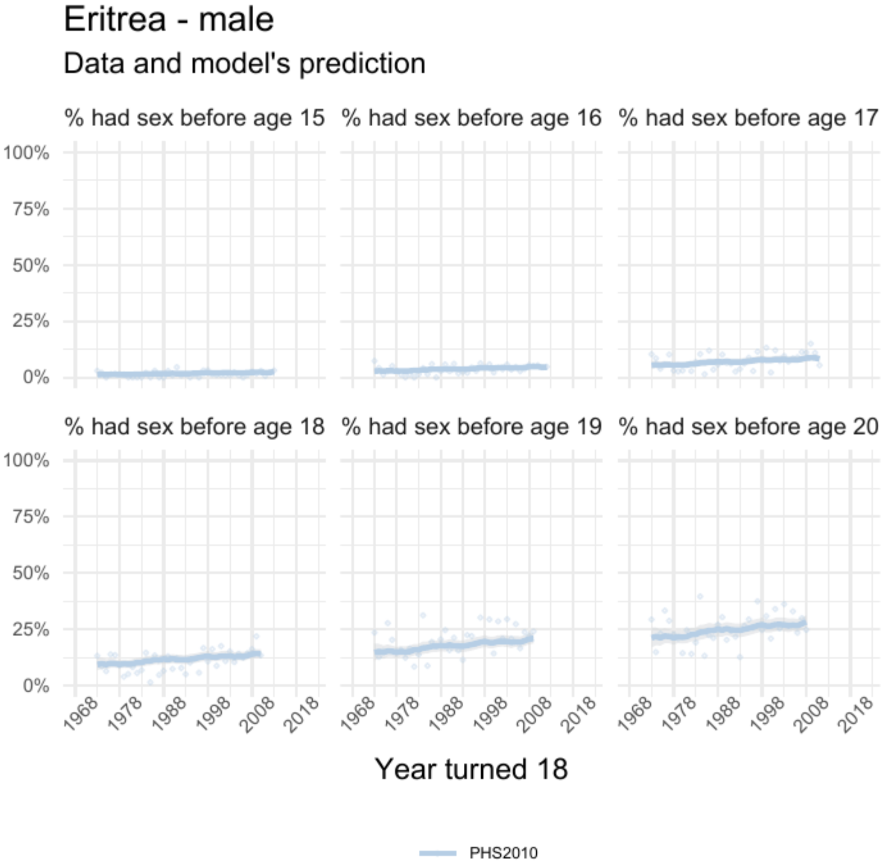


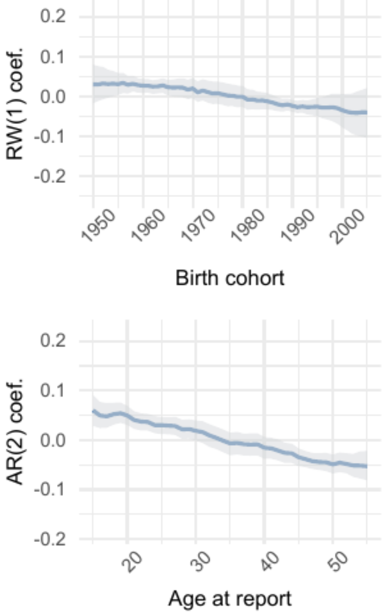


| | Parameter | Estimate |
|---|---|---|
| 1 | intercept | 0.02 [-0.04 – 0.07] |
| 2 | skew | 3.20 [2.57 – 3.93] |
| 3 | shape | 5.36 [5.12 – 5.64] |

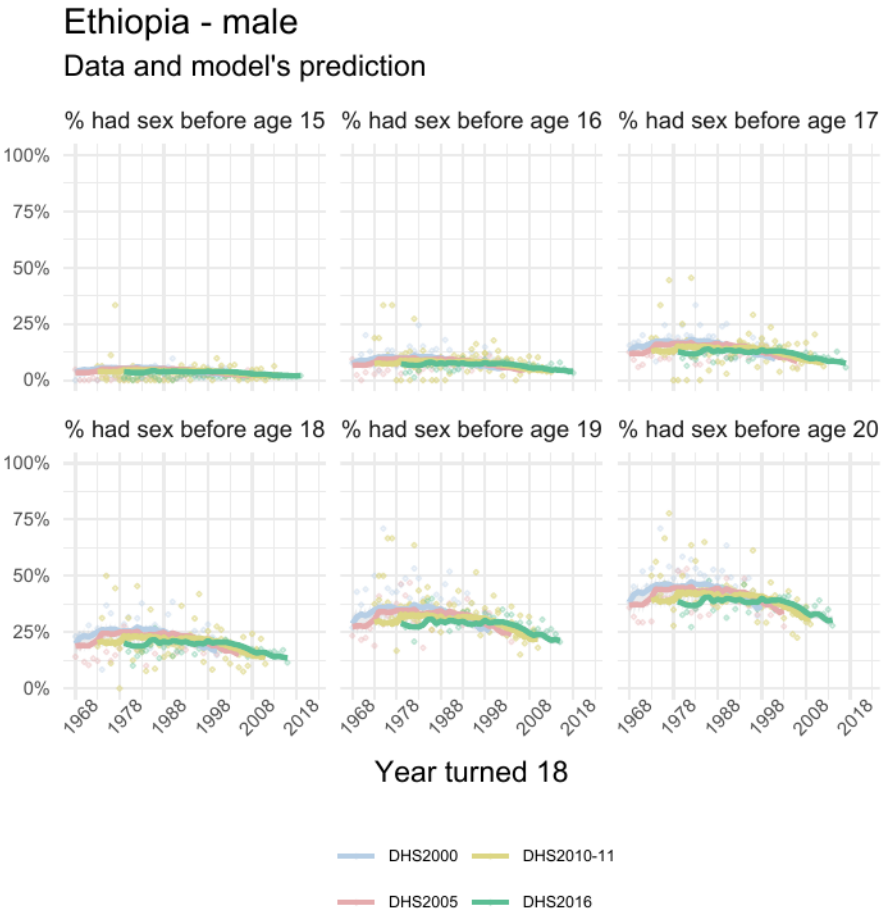


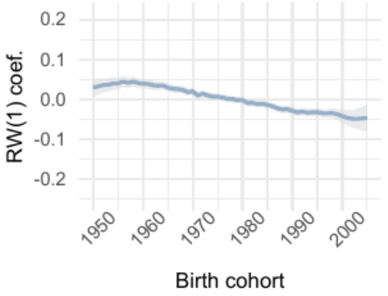


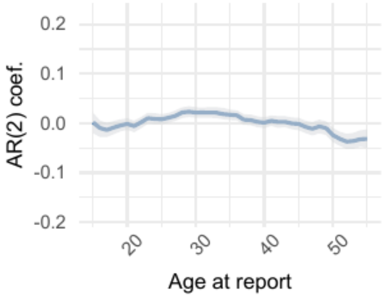


| | Parameter | Estimate |
|---|---|---|
| 1 | intercept | -0.020 [-0.035 – -0.007] |
| 2 | skew | 2.04 [1.91 – 2.16] |
| 3 | shape | 6.65 [6.55 – 6.76] |

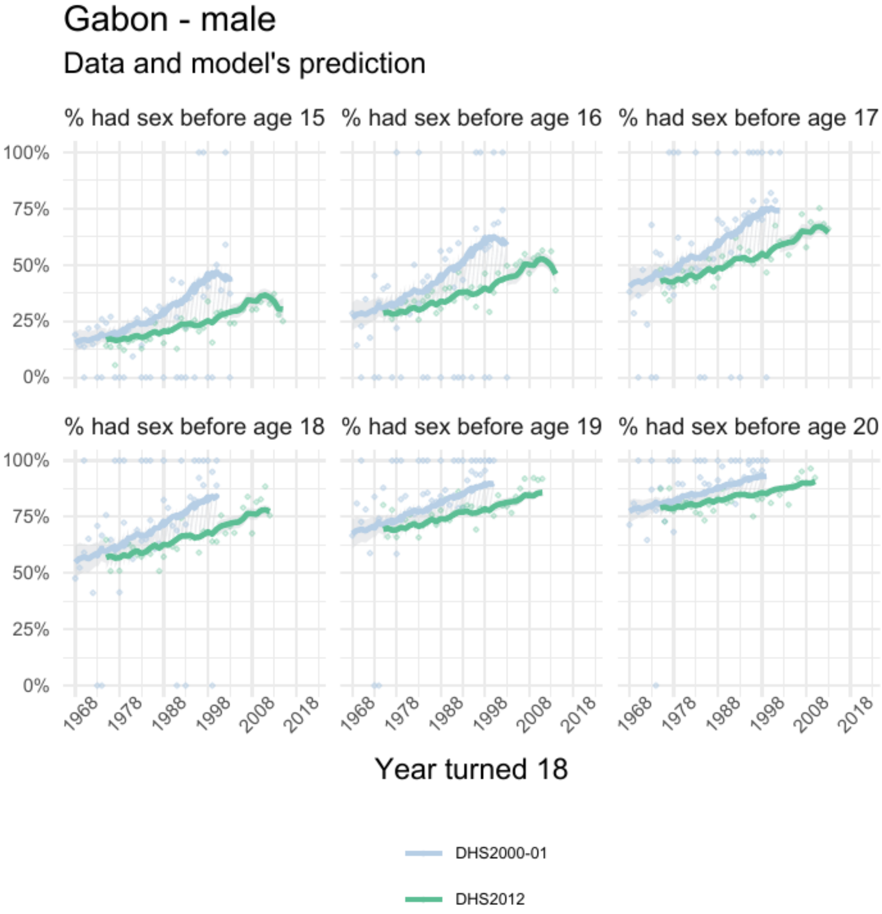


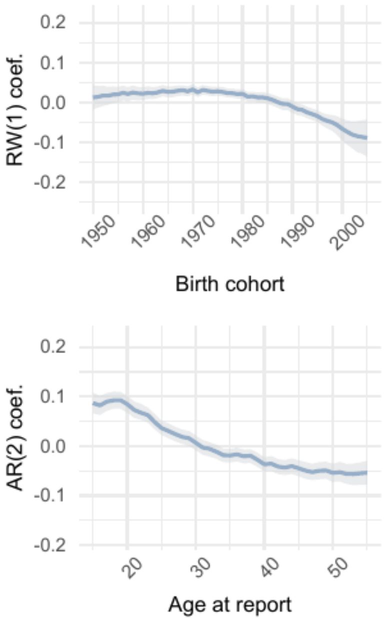


| | Parameter | Estimate |
|---|---|---|
| *1* | intercept | 0.10 [0.09 – 0.12] |
| *2* | skew | 1.21 [1.11 – 1.33] |
| *3* | shape | 9.57 [ 9.22 – 9.90] |

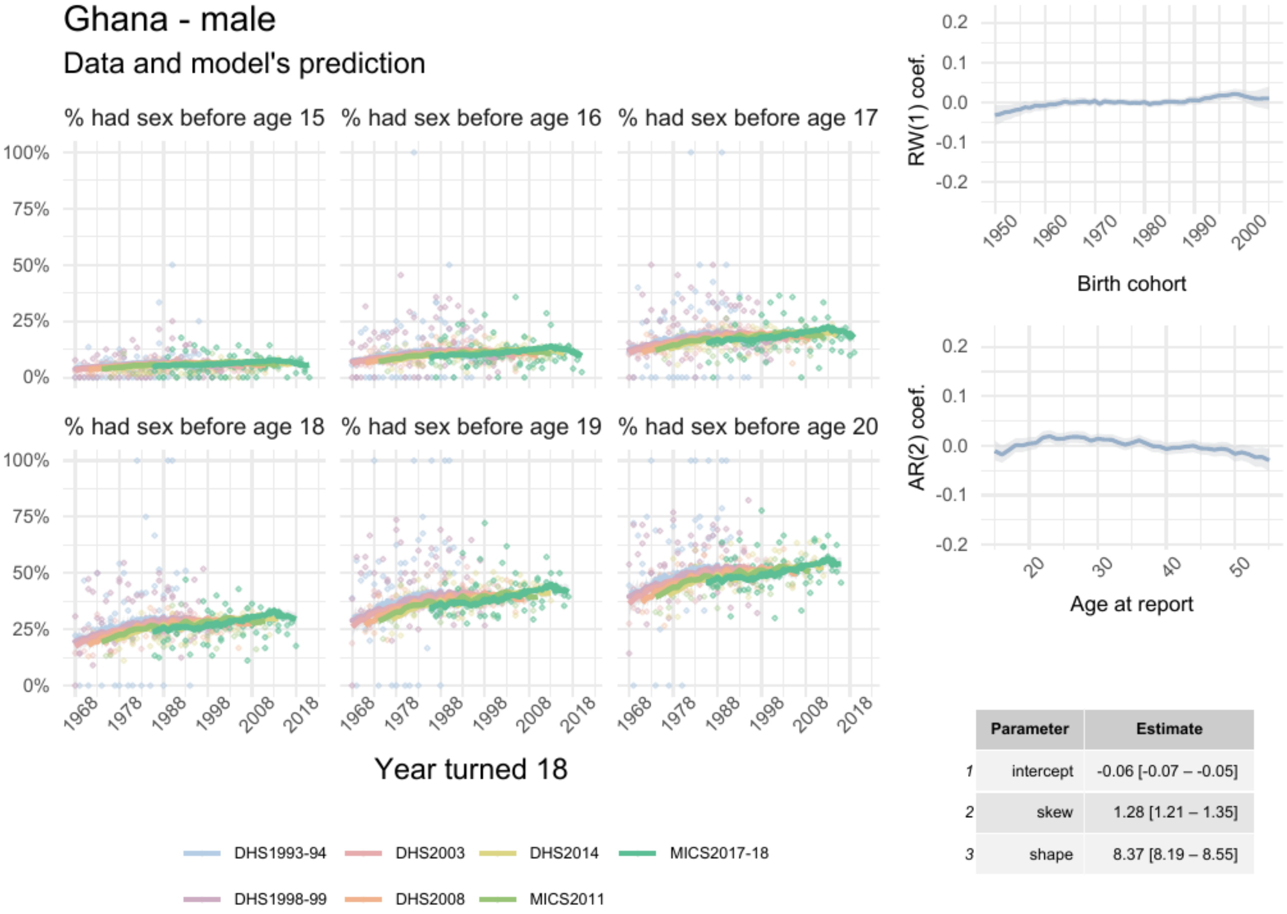


| | Parameter | Estimate |
|---|---|---|
| 1 | intercept | -0.06 [-0.07 – -0.05] |
| 2 | skew | 1.28 [1.21 – 1.35] |
| 3 | shape | 8.37 [8.19 – 8.55] |

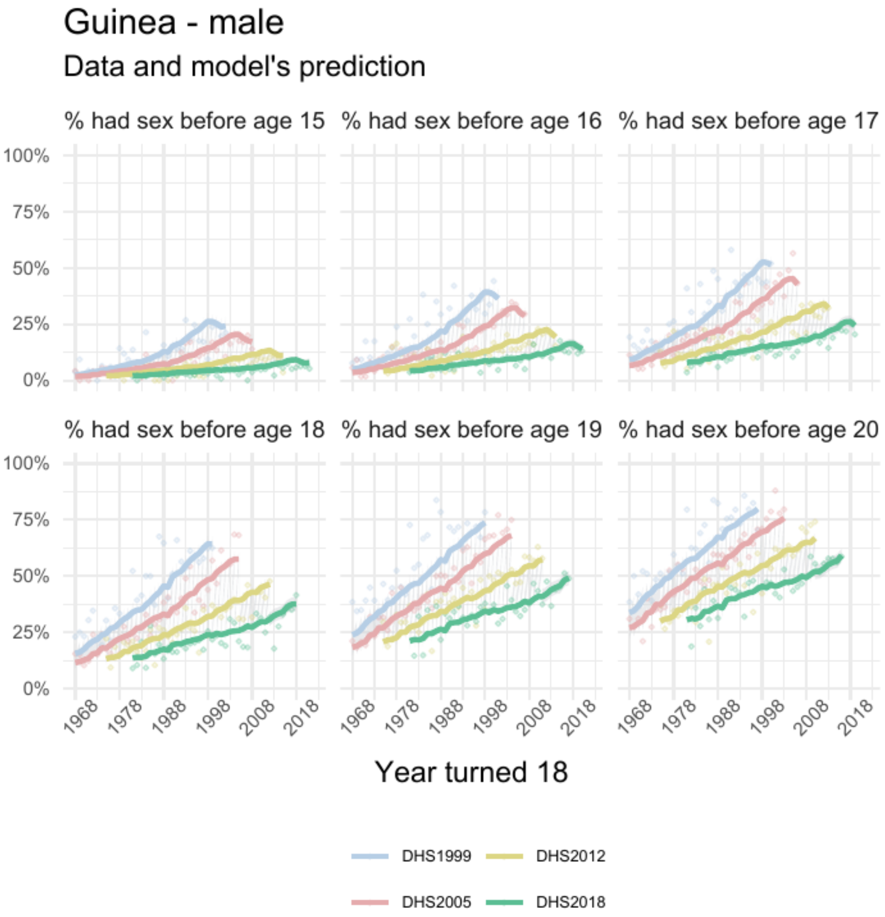


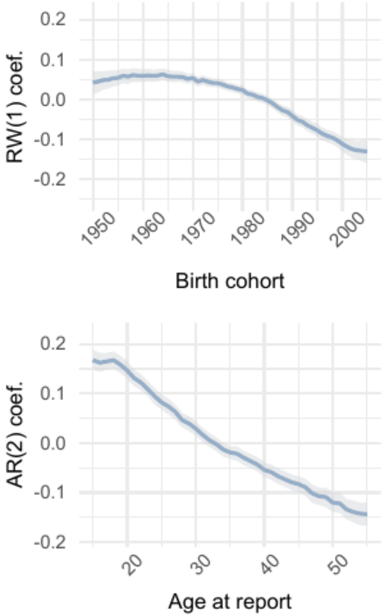


| | Parameter | Estimate |
|---|---|---|
| 1 | intercept | -0.023 [-0.041 – -0.005] |
| 2 | skew | 1.57 [1.44 – 1.73] |
| 3 | shape | 7.78 [7.53 – 8.02] |

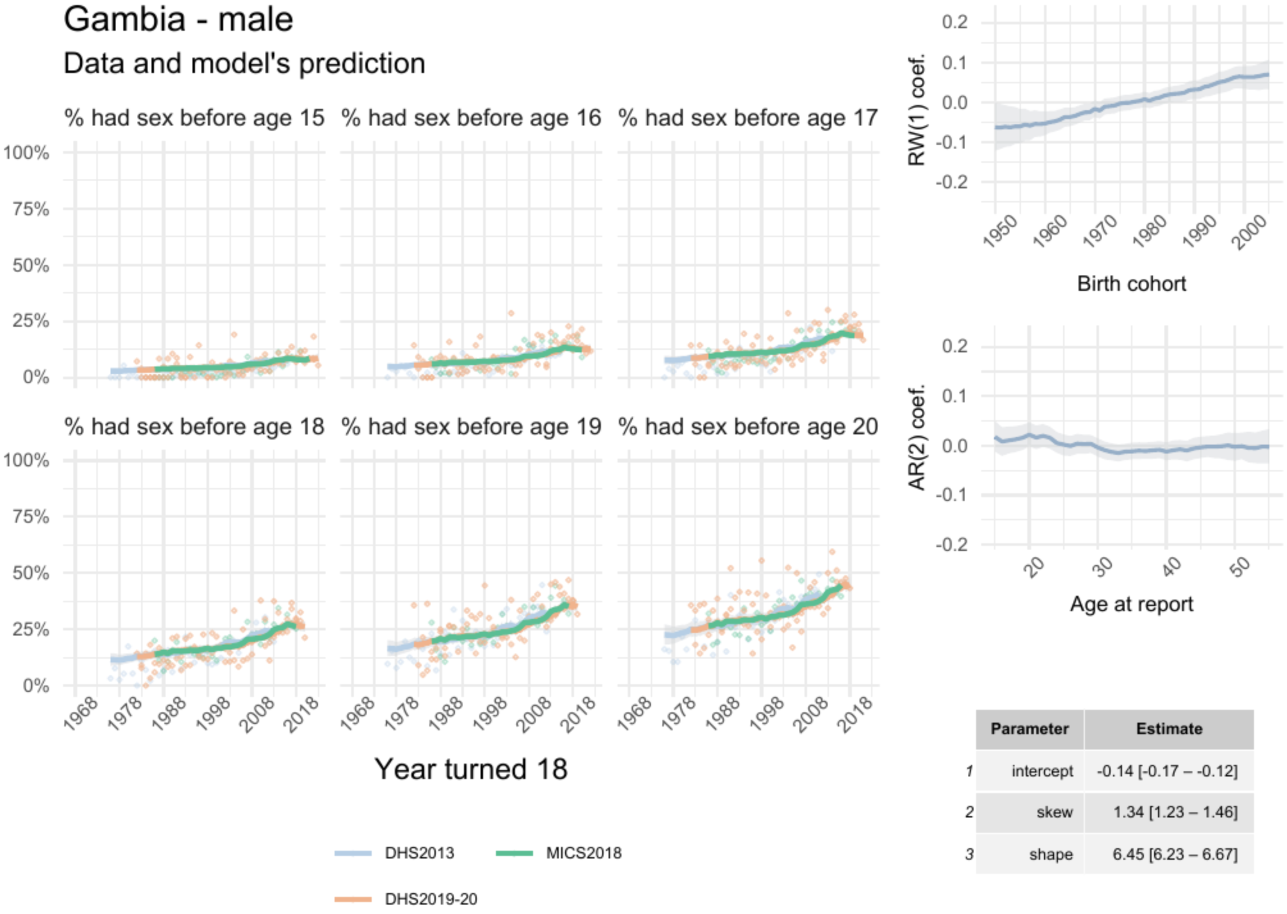


| | Parameter | Estimate |
|---|---|---|
| 1 | intercept | -0.14 [-0.17 – -0.12] |
| 2 | skew | 1.34 [1.23 – 1.46] |
| 3 | shape | 6.45 [6.23 – 6.67] |

### Guinea-Bissau - male

#### Data and model's prediction

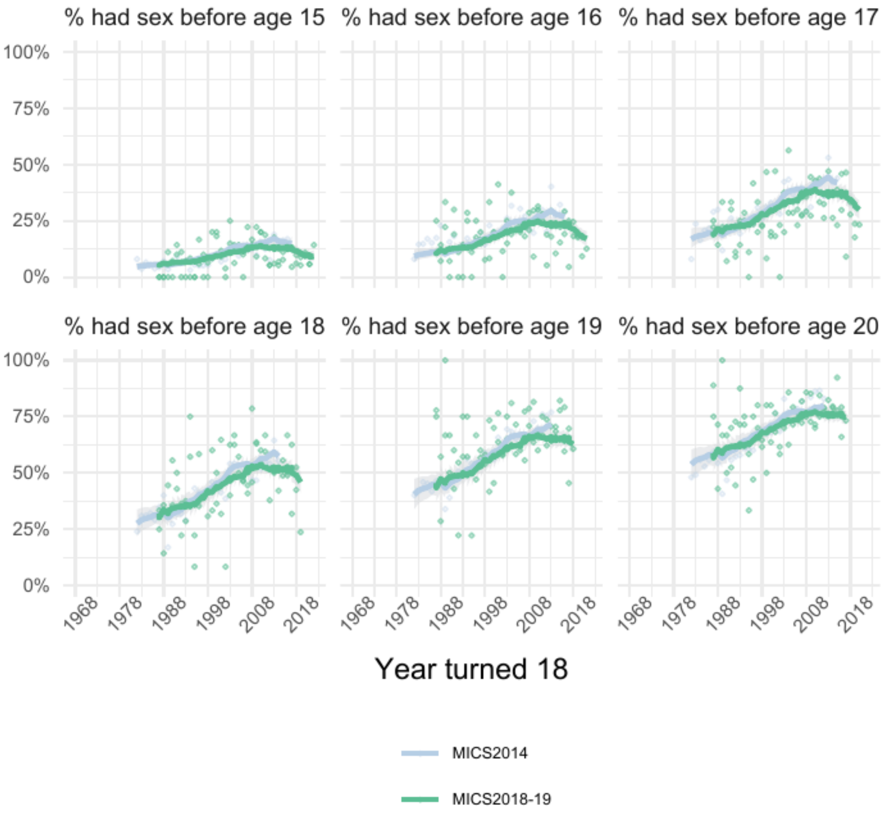


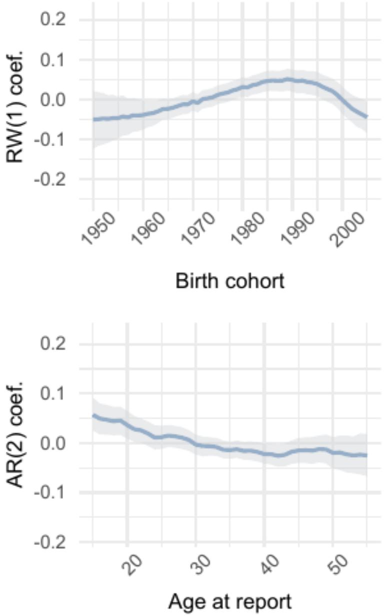


| | Parameter | Estimate |
|---|---|---|
| 1 | intercept | -0.003 [-0.025 – 0.020] |
| 2 | skew | 1.24 [1.12 – 1.38] |
| 3 | shape | 9.80 [ 9.41 – 10.22] |

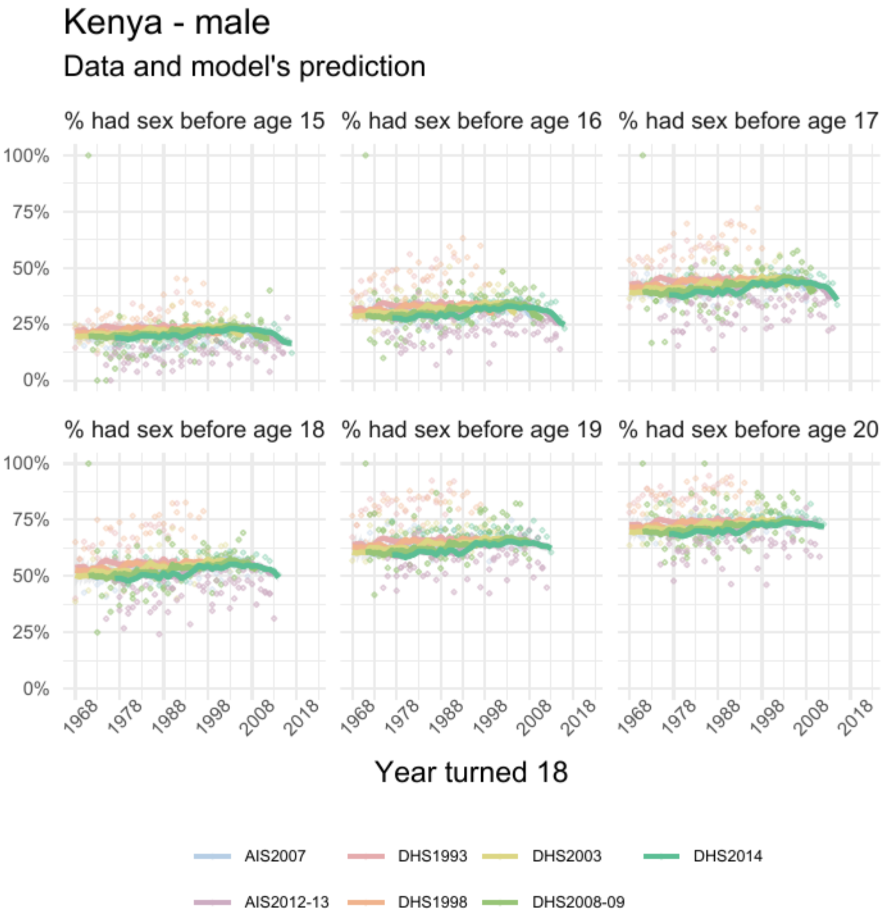


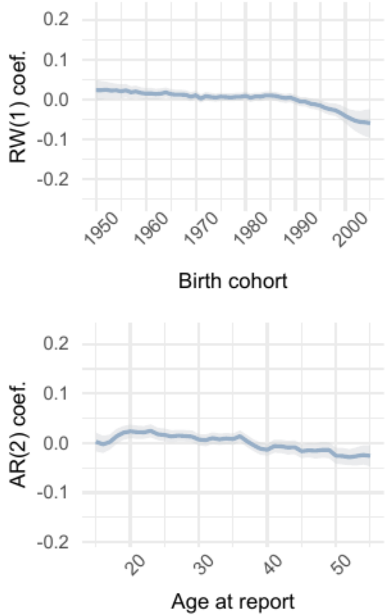


| | Parameter | Estimate |
|---|---|---|
| 1 | intercept | 9e-03 [-5e-04 – 2e-02] |
| 2 | skew | 0.95 [0.92 – 0.99] |
| 3 | shape | 7.94 [7.80 – 8.08] |

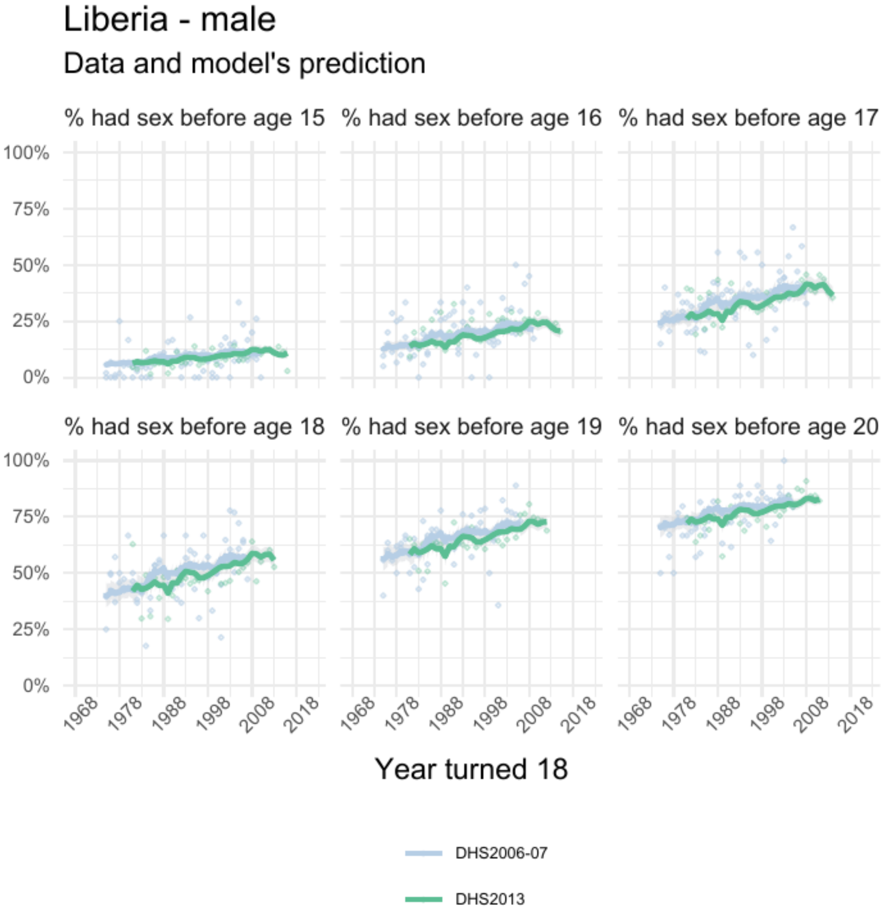


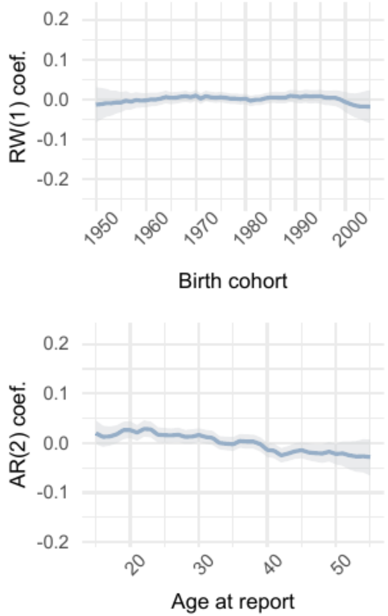


| | Parameter | Estimate |
|---|---|---|
| 1 | intercept | 0.04 [0.02 – 0.05] |
| 2 | skew | 1.27 [1.18 – 1.39] |
| 3 | shape | 11.40 [11.05 – 11.74] |

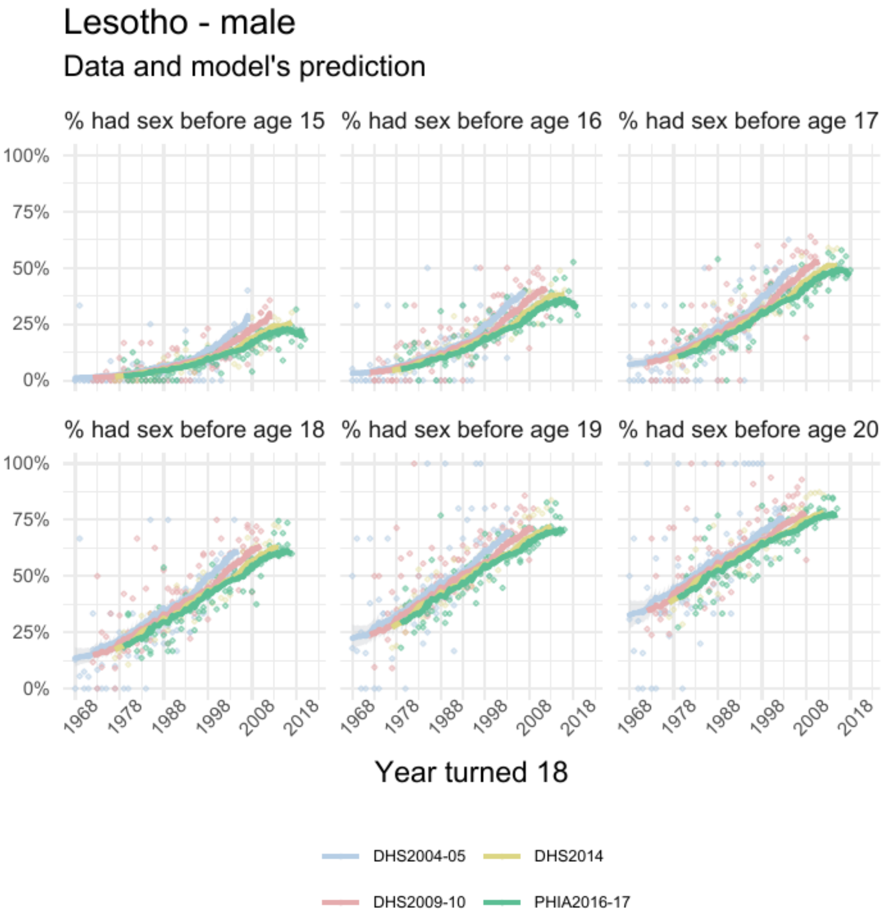


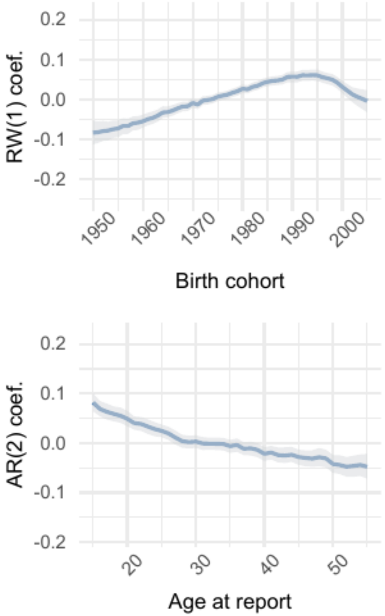


| | Parameter | Estimate |
|---|---|---|
| 1 | intercept | 0.16 [0.14 – 0.18] |
| 2 | skew | 3.11 [2.81 – 3.43] |
| 3 | shape | 6.96 [6.80 – 7.11] |

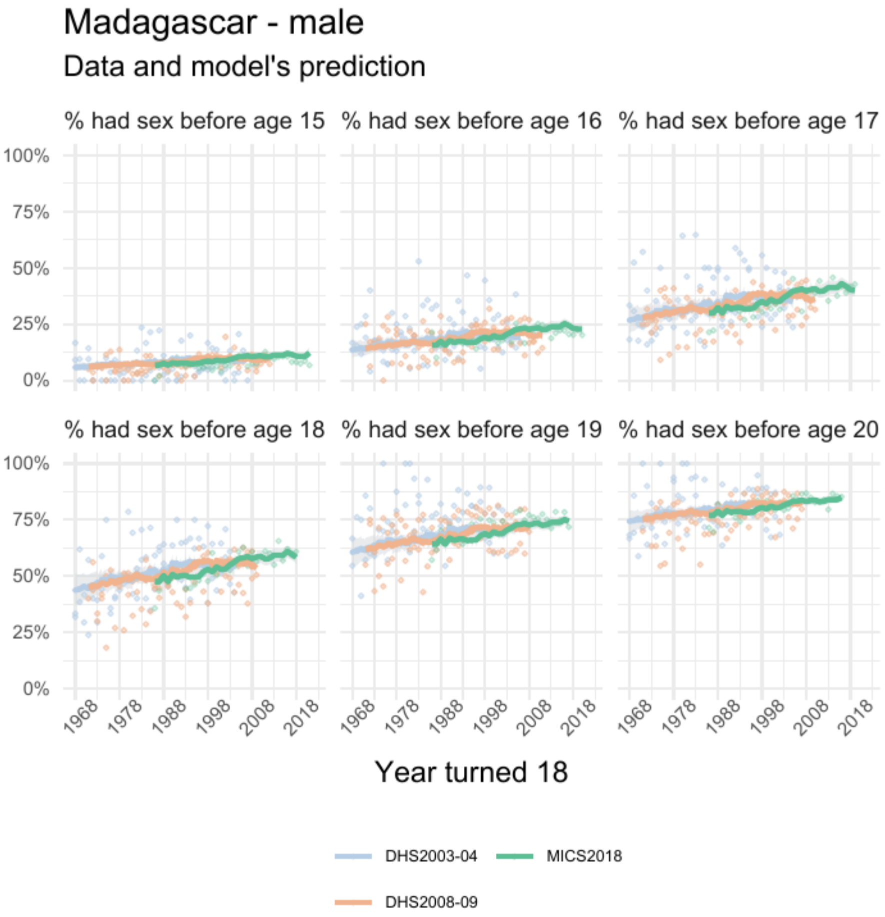


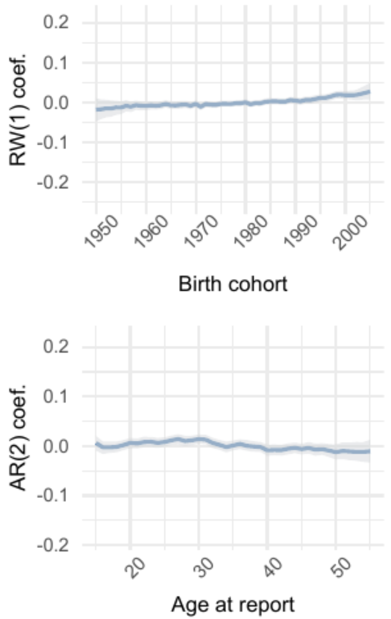


| | Parameter | Estimate |
|---|---|---|
| 1 | intercept | 0.06 [0.05 – 0.07] |
| 2 | skew | 1.38 [1.29 – 1.45] |
| 3 | shape | 11.68 [11.40 – 11.96] |

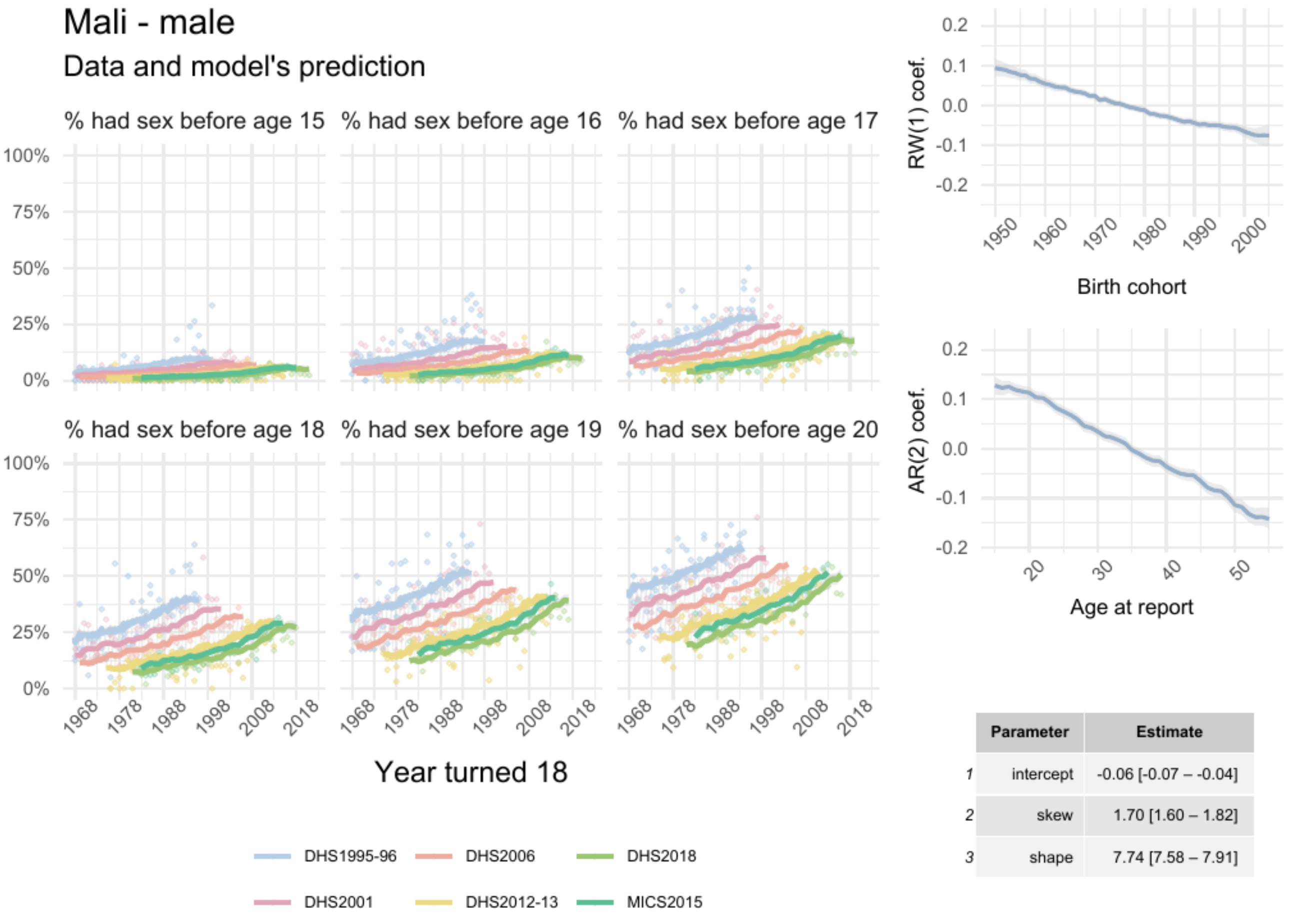


| | Parameter | Estimate |
|---|---|---|
| 1 | intercept | -0.06 [-0.07 – -0.04] |
| 2 | skew | 1.70 [1.60 – 1.82] |
| 3 | shape | 7.74 [7.58 – 7.91] |

### Mozambique - male

#### Data and model's prediction

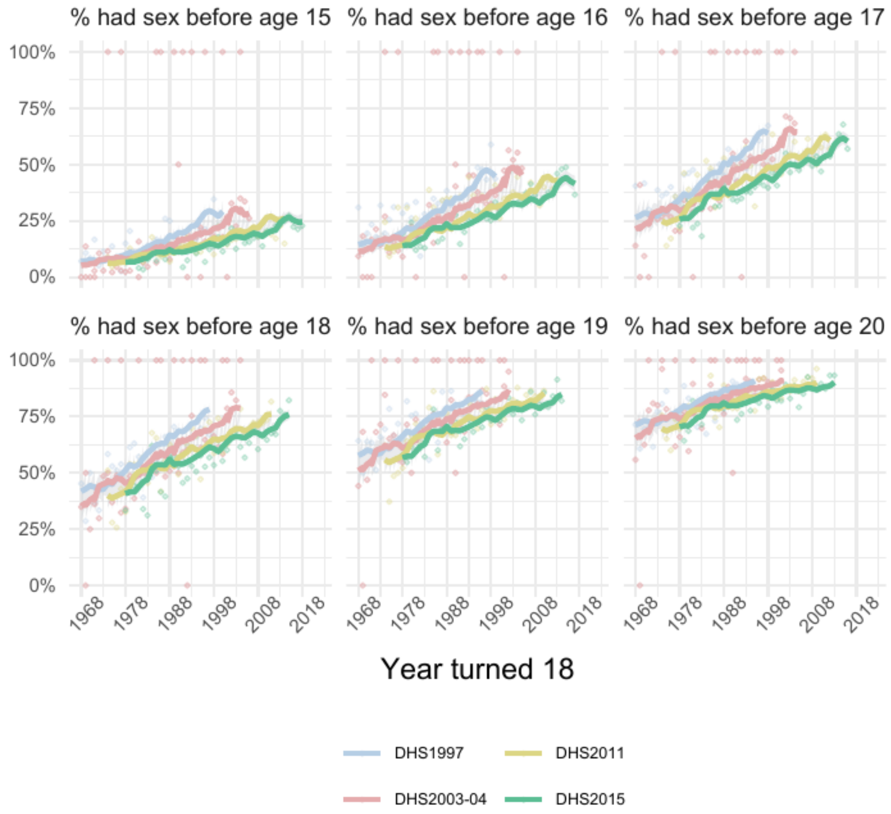


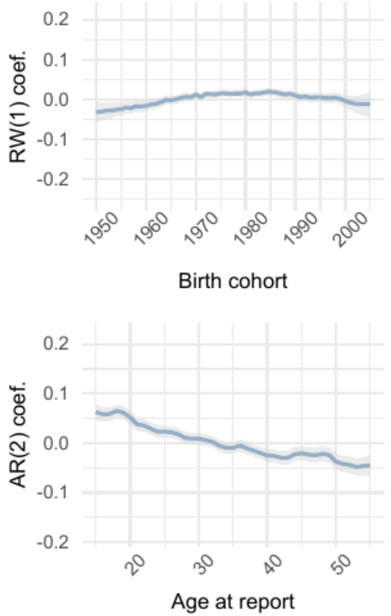


| | Parameter | Estimate |
|---|---|---|
| 1 | intercept | 0.07 [0.06 – 0.08] |
| 2 | skew | 1.21 [1.12 – 1.30] |
| 3 | shape | 11.22 [10.89 – 11.52] |

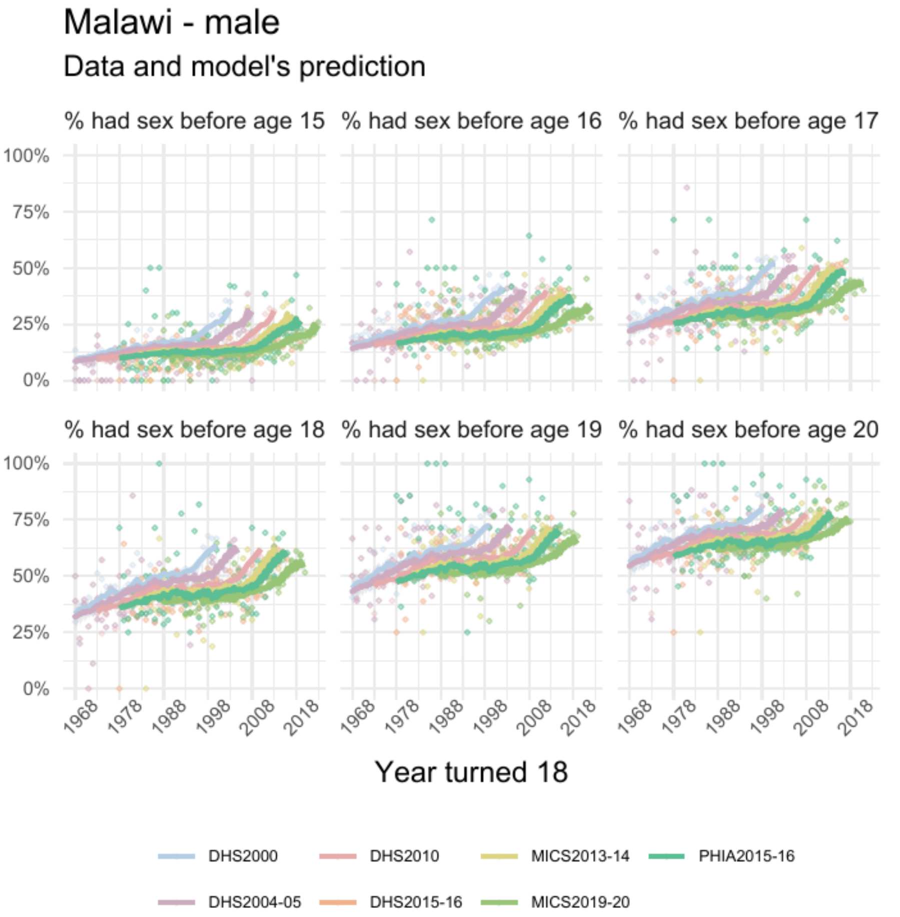


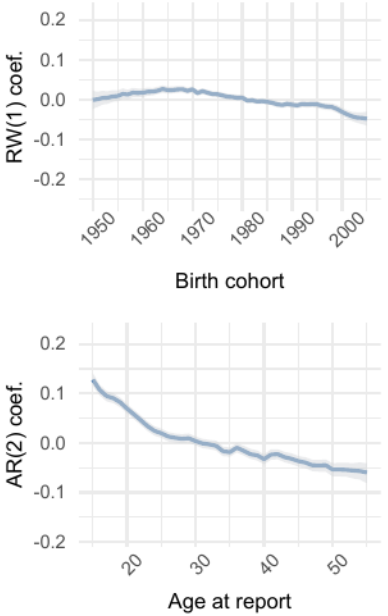


| | Parameter | Estimate |
|---|---|---|
| 1 | intercept | -0.013 [-0.021 – -0.005] |
| 2 | skew | 0.98 [0.95 – 1.02] |
| 3 | shape | 8.96 [8.81 – 9.10] |

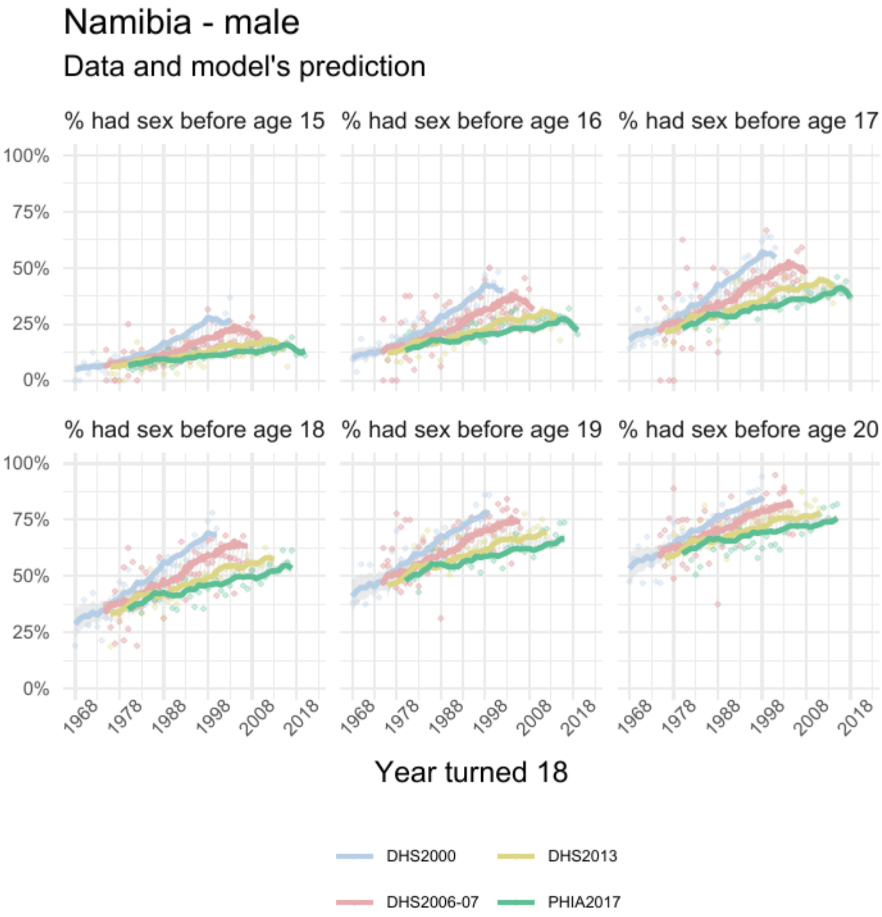


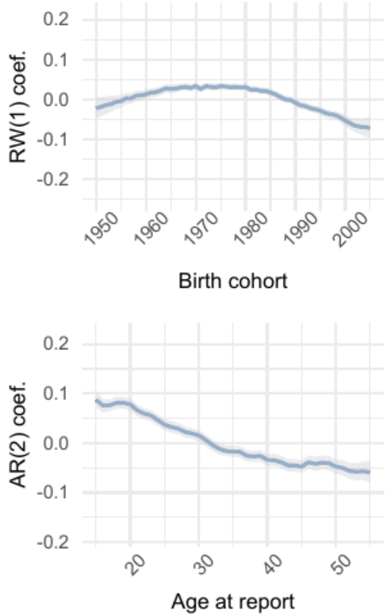


| | Parameter | Estimate |
|---|---|---|
| *1* | intercept | 0.07 [0.06 – 0.09] |
| *2* | skew | 1.68 [1.58 – 1.79] |
| *3* | shape | 8.39 [8.20 – 8.57] |

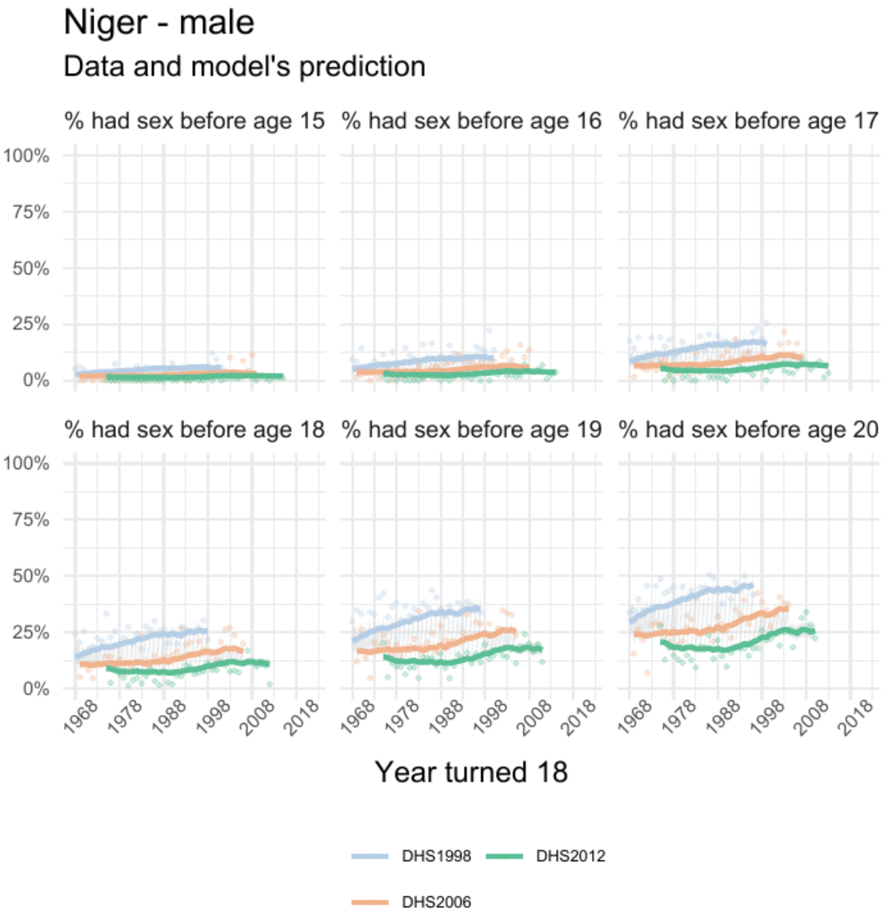


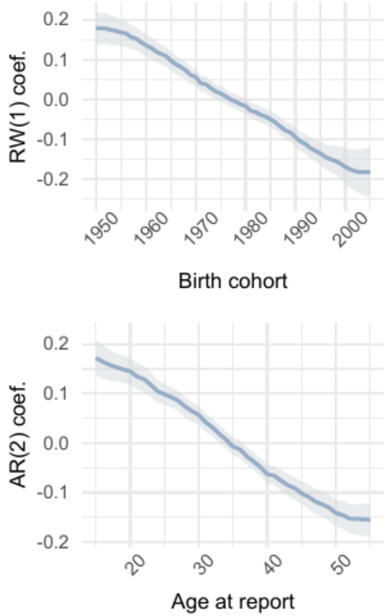


| | Parameter | Estimate |
|---|---|---|
| 1 | intercept | -0.18 [-0.21 – -0.16] |
| 2 | skew | 1.43 [1.28 – 1.61] |
| 3 | shape | 7.49 [7.18 – 7.82] |

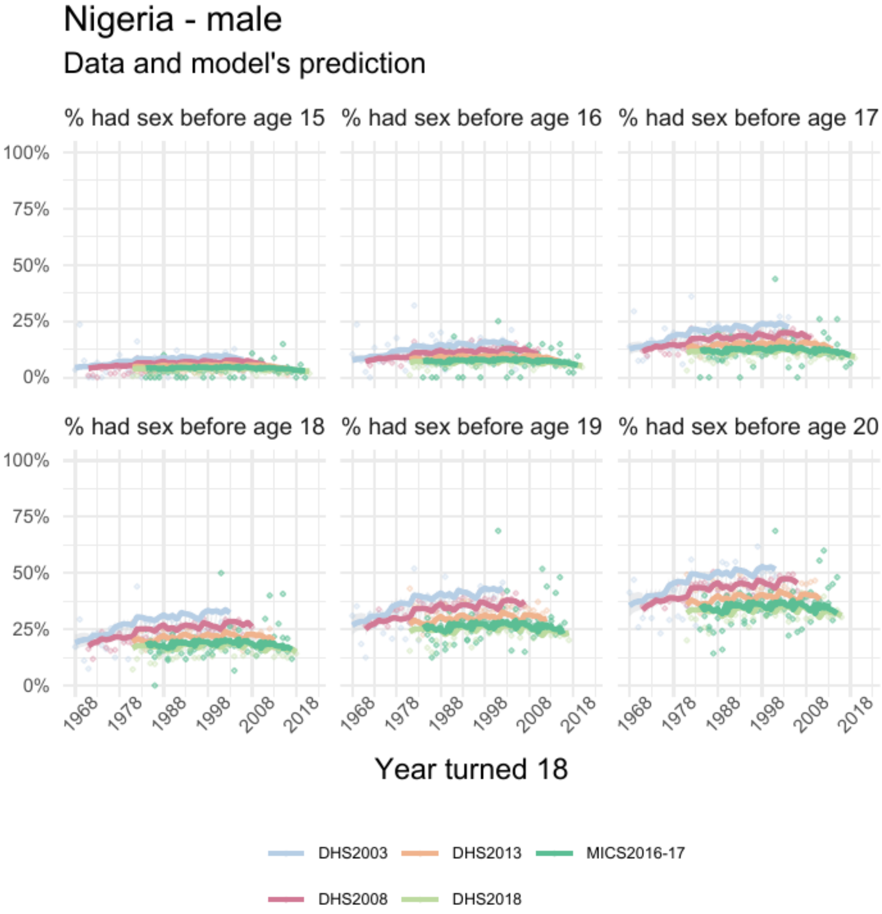


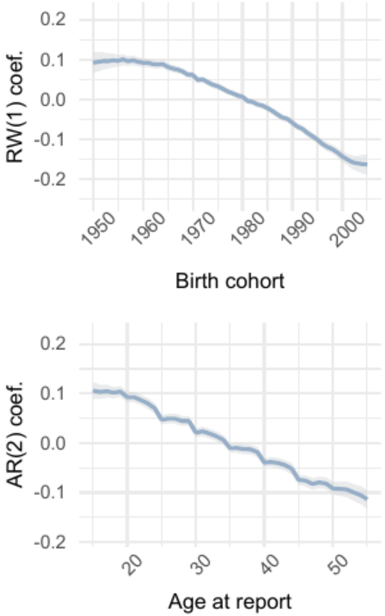


| | Parameter | Estimate |
|---|---|---|
| 1 | intercept | -0.07 [-0.08 – -0.06] |
| 2 | skew | 1.57 [1.52 – 1.63] |
| 3 | shape | 6.66 [6.56 – 6.74] |

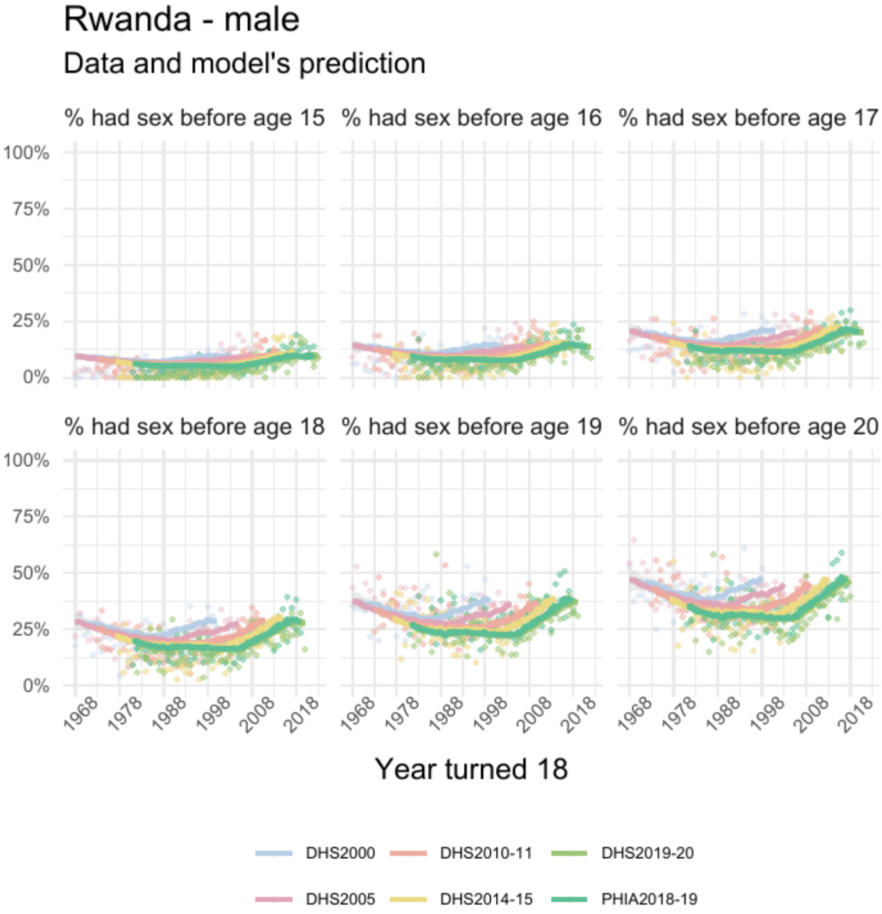


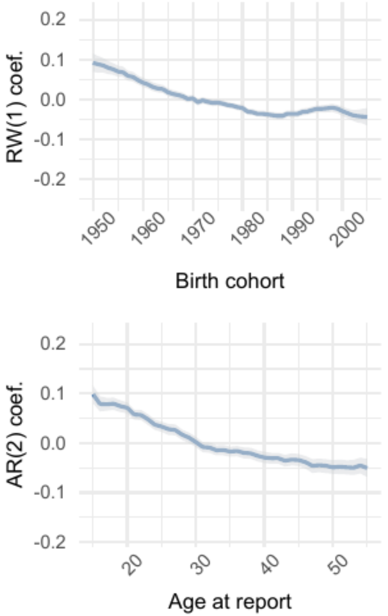


| | Parameter | Estimate |
|---|---|---|
| *1* | intercept | -0.18 [-0.19 – -0.17] |
| *2* | skew | 0.85 [0.82 – 0.89] |
| *3* | shape | 8.17 [8.01 – 8.34] |

### Sudan - male

#### Data and model's prediction

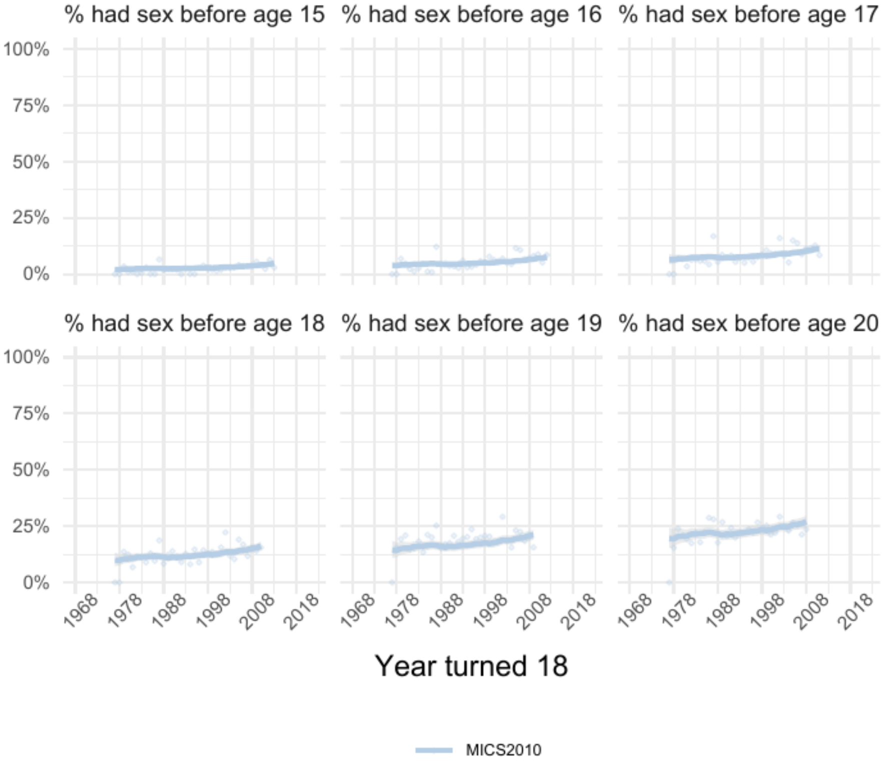


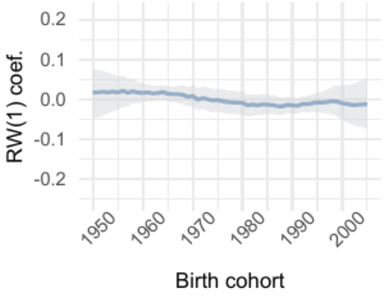


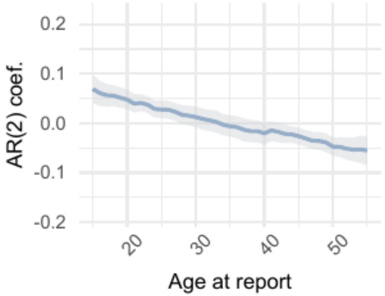


| | Parameter | Estimate |
|---|---|---|
| 1 | intercept | 0.03 [-0.04 – 0.10] |
| 2 | skew | 3.17 [2.56 – 3.93] |
| 3 | shape | 4.42 [4.22 – 4.66] |

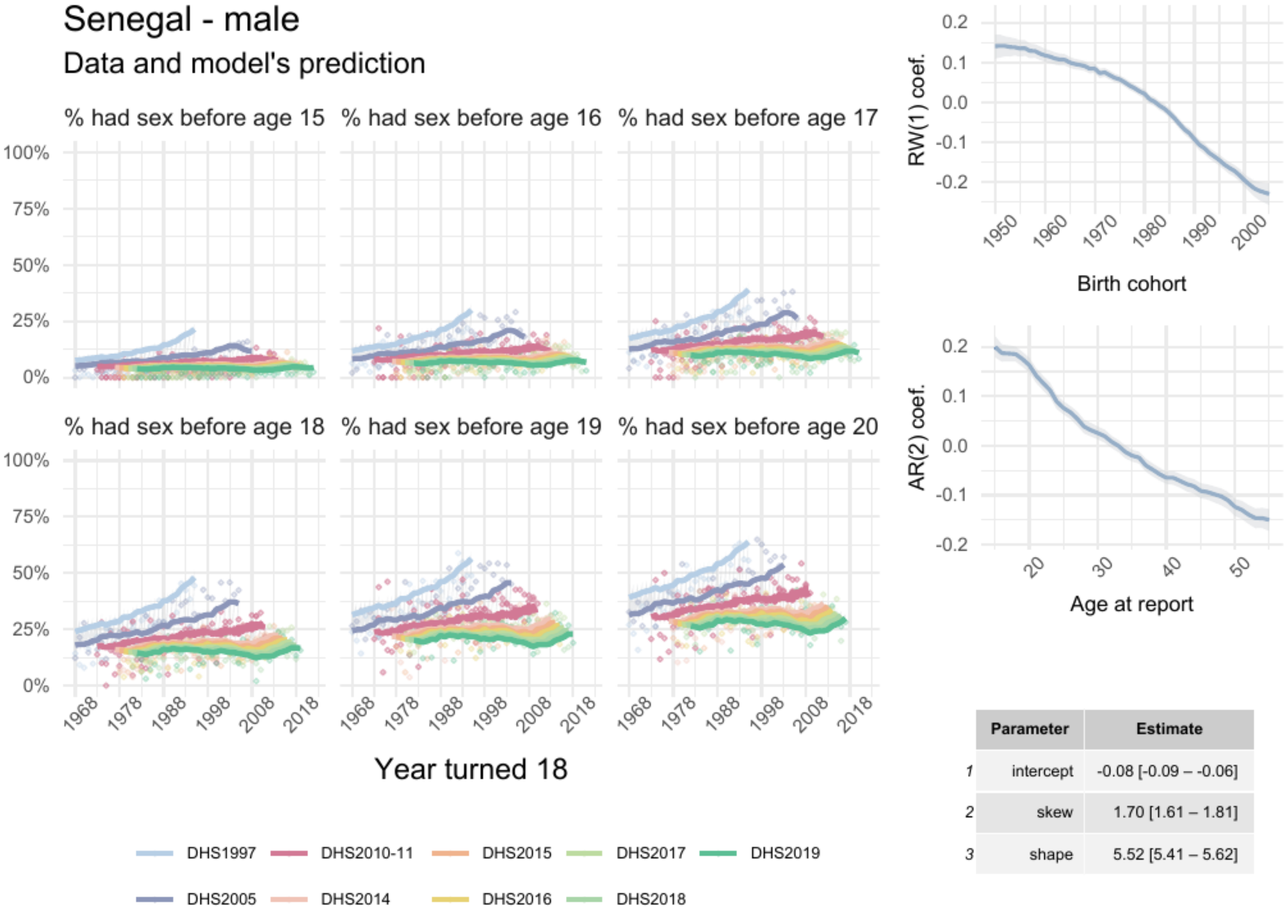


| | Parameter | Estimate |
|---|---|---|
| 1 | intercept | -0.08 [-0.09 – -0.06] |
| 2 | skew | 1.70 [1.61 – 1.81] |
| 3 | shape | 5.52 [5.41 – 5.62] |

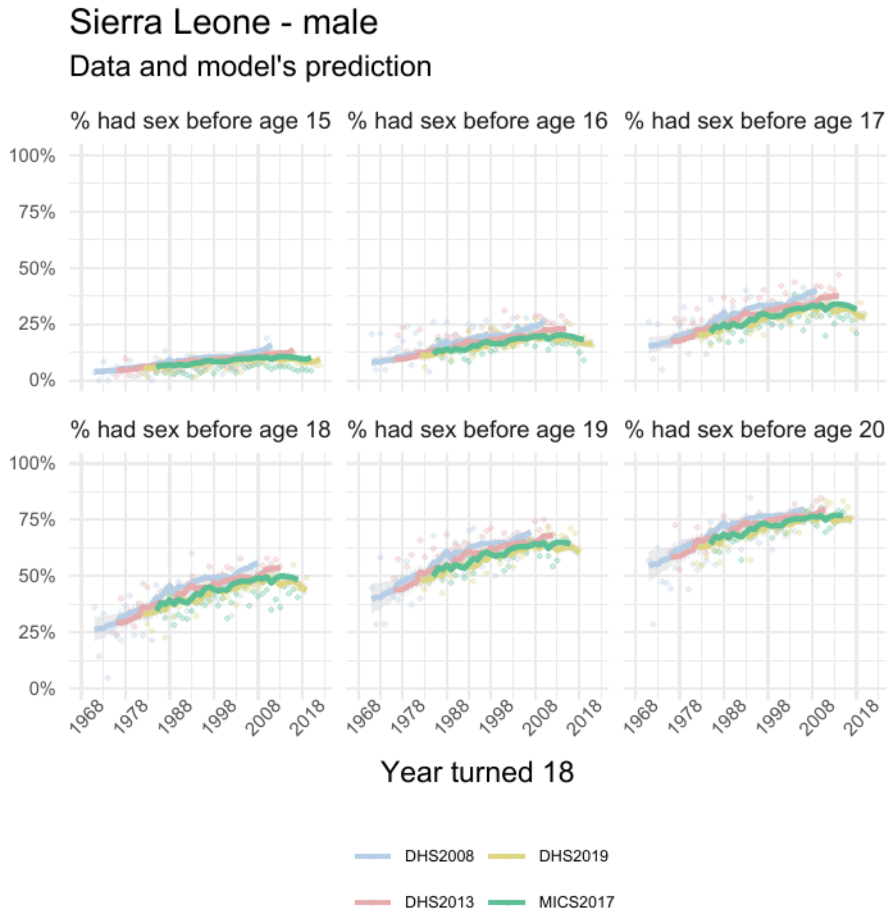


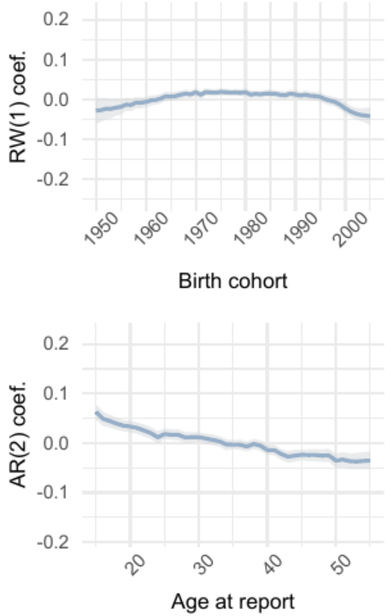


| | Parameter | Estimate |
|---|---|---|
| 1 | intercept | -0.011 [-0.020 – -0.001] |
| 2 | skew | 1.05 [1.01 – 1.11] |
| 3 | shape | 11.39 [11.15 – 11.62] |

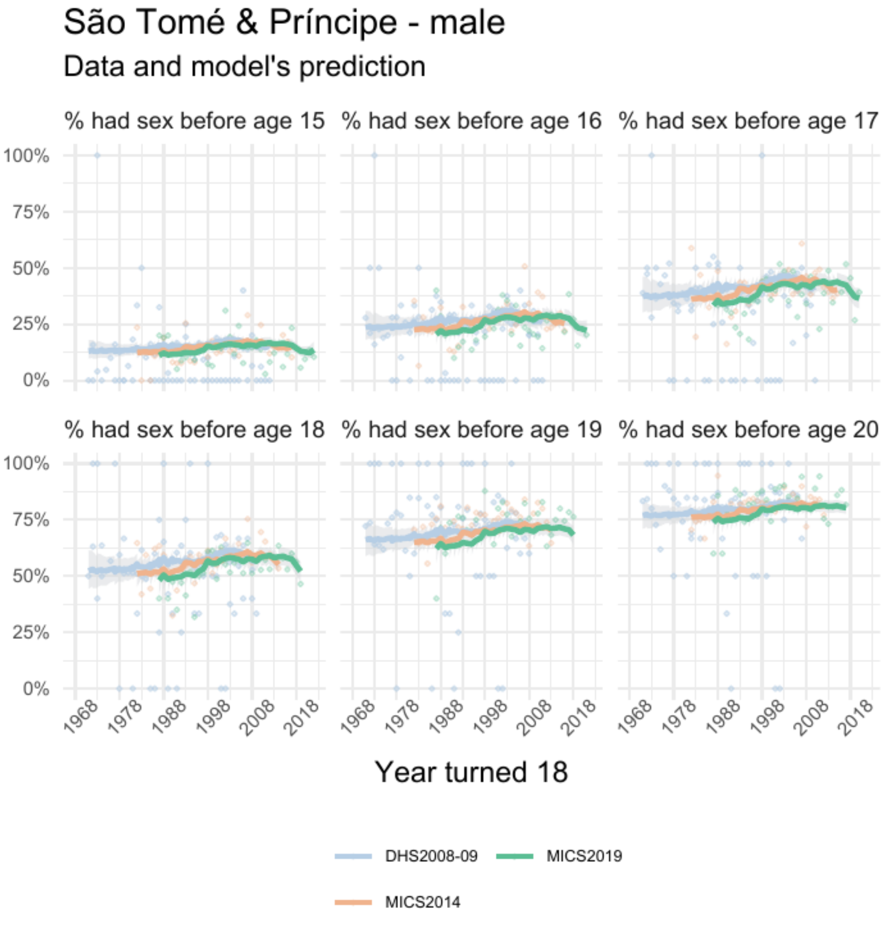


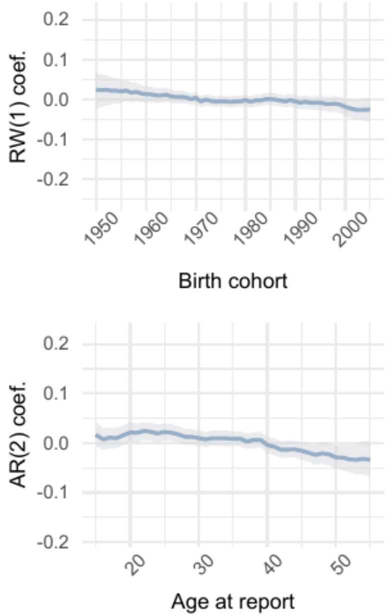


| | Parameter | Estimate |
|---|---|---|
| 1 | intercept | 0.05 [0.04 – 0.07] |
| 2 | skew | 1.13 [1.04 – 1.23] |
| 3 | shape | 10.32 [ 9.92 – 10.72] |

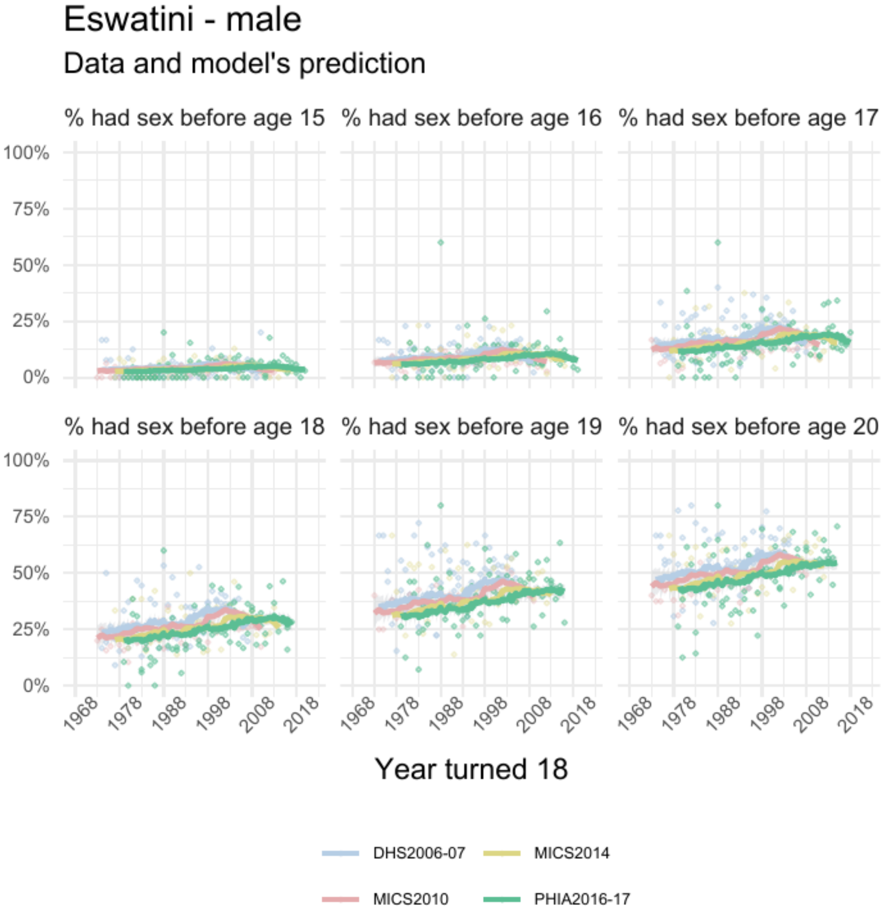


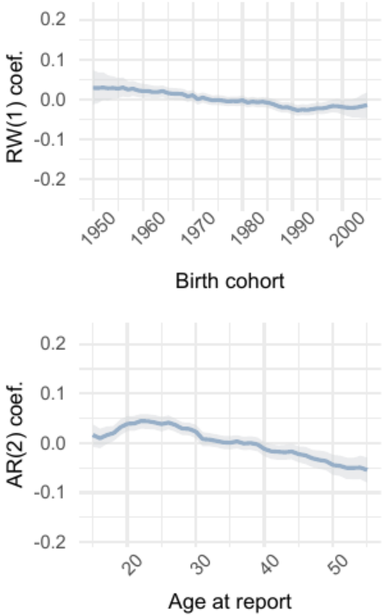


| | Parameter | Estimate |
|---|---|---|
| 1 | intercept | 0.012 [-0.005 – 0.028] |
| 2 | skew | 1.90 [1.75 – 2.06] |
| 3 | shape | 8.08 [7.86 – 8.30] |

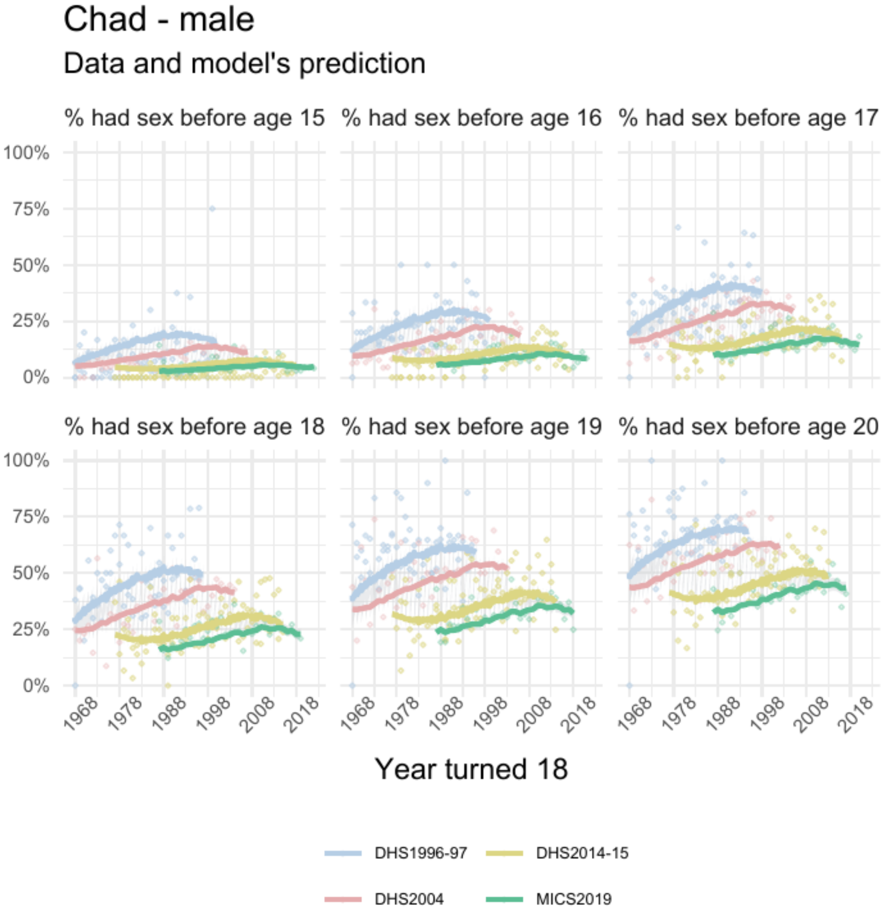


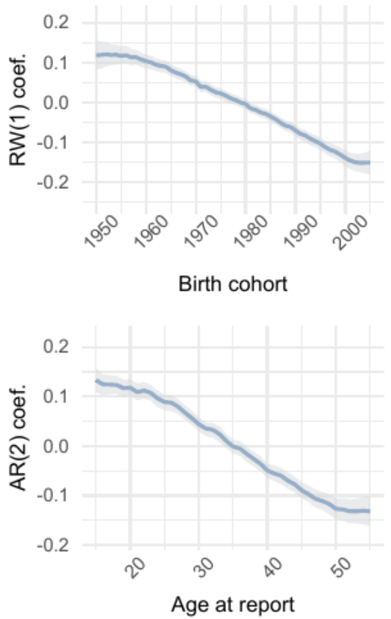


| | Parameter | Estimate |
|---|---|---|
| 1 | intercept | 0.04 [0.01 – 0.06] |
| 2 | skew | 2.13 [1.93 – 2.34] |
| 3 | shape | 6.42 [6.24 – 6.60] |

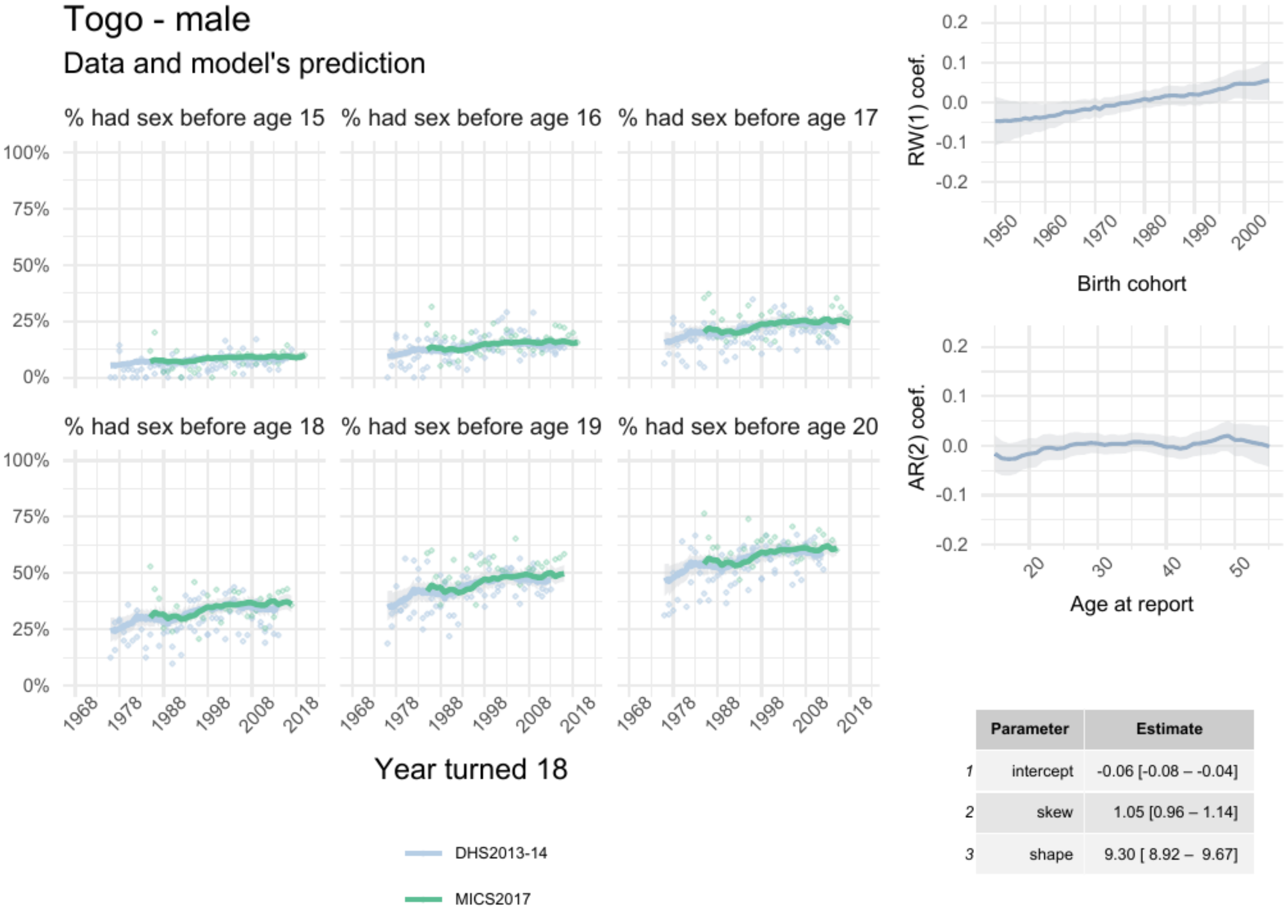


| | Parameter | Estimate |
|---|---|---|
| 1 | intercept | -0.06 [-0.08 – -0.04] |
| 2 | skew | 1.05 [0.96 – 1.14] |
| 3 | shape | 9.30 [ 8.92 – 9.67] |

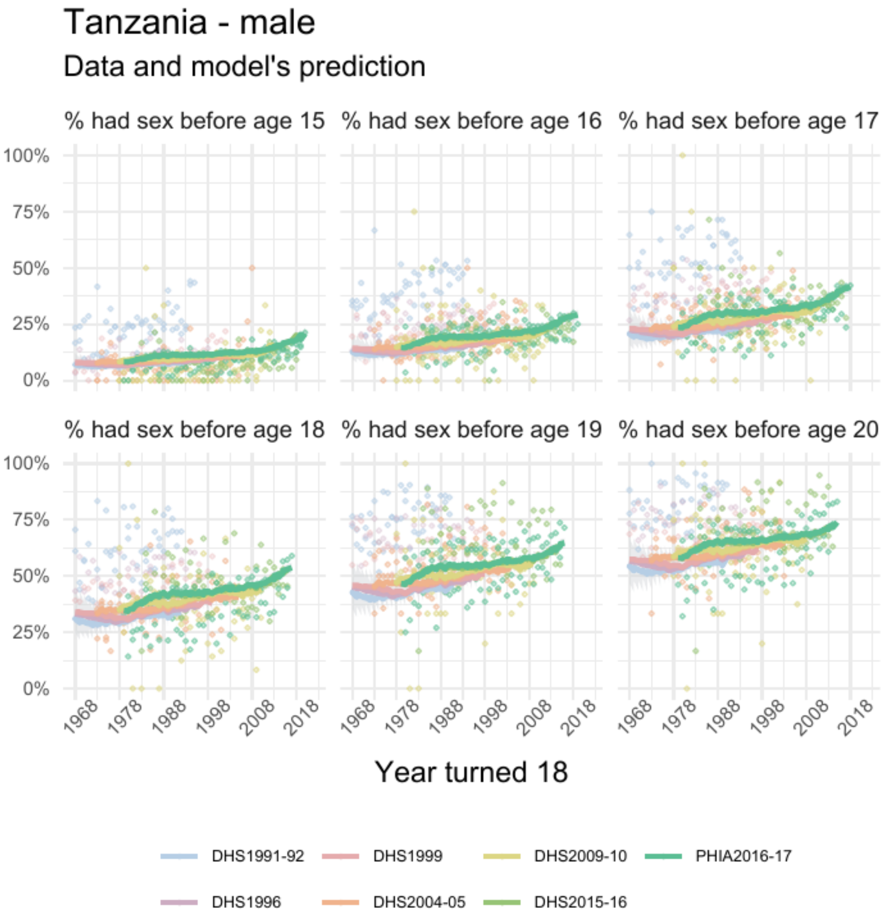


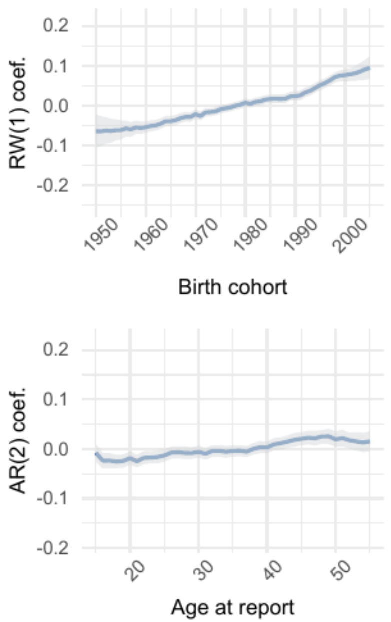


| | Parameter | Estimate |
|---|---|---|
| 1 | intercept | 0.017 [0.007 – 0.029] |
| 2 | skew | 1.27 [1.20 – 1.33] |
| 3 | shape | 8.50 [8.34 – 8.68] |

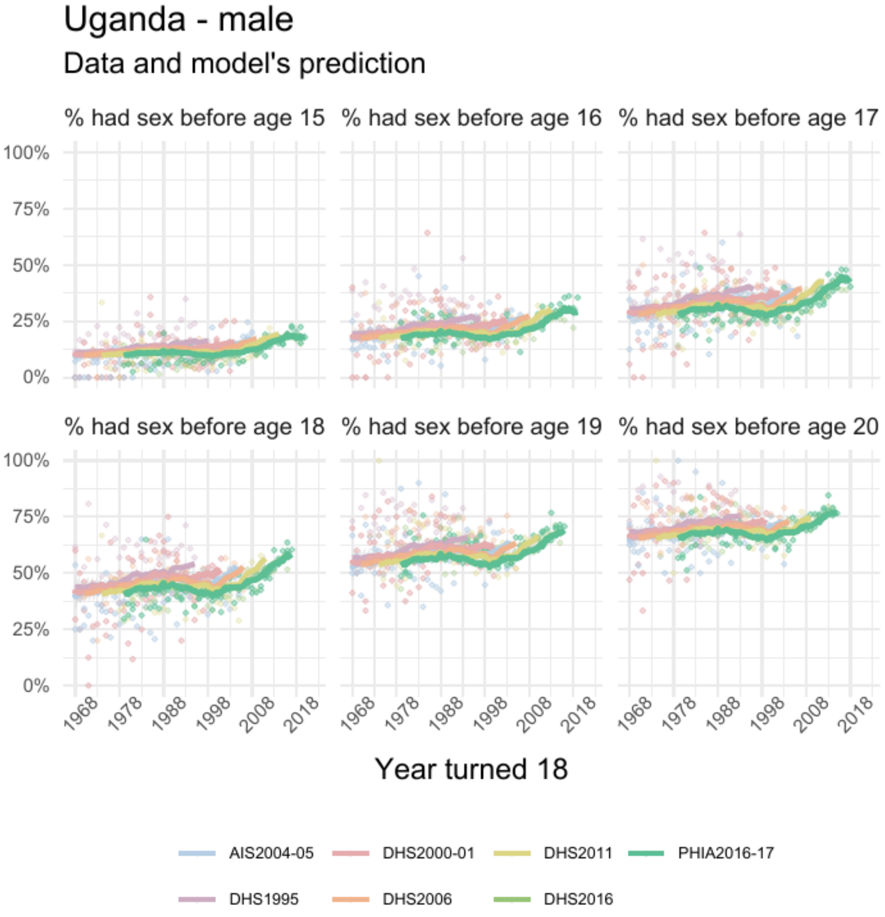


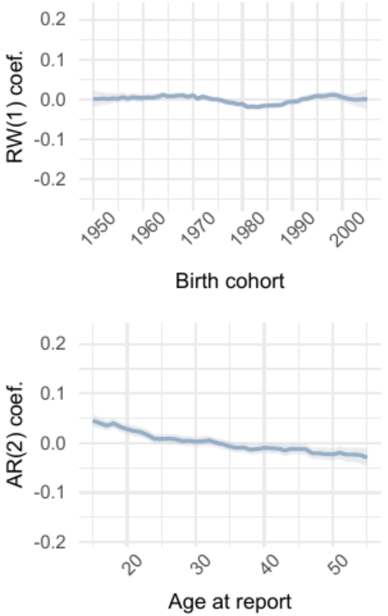


| | Parameter | Estimate |
|---|---|---|
| 1 | intercept | 0.02 [0.01 – 0.03] |
| 2 | skew | 1.17 [1.12 – 1.22] |
| 3 | shape | 9.27 [9.12 – 9.43] |

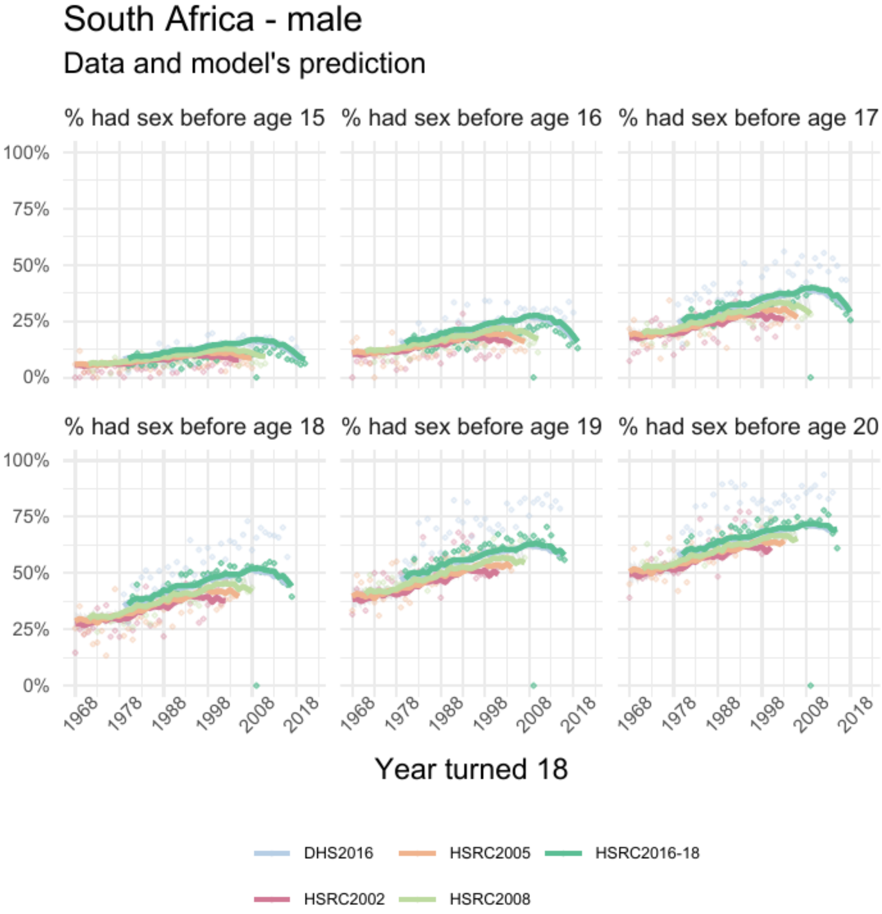


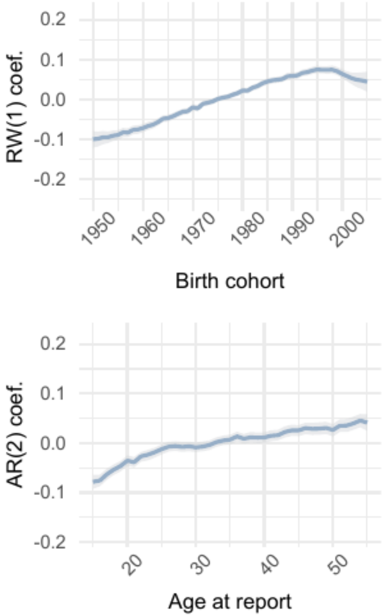


| | Parameter | Estimate |
|---|---|---|
| 1 | intercept | 0.08 [0.07 – 0.09] |
| 2 | skew | 1.85 [1.78 – 1.92] |
| 3 | shape | 7.37 [7.27 – 7.47] |

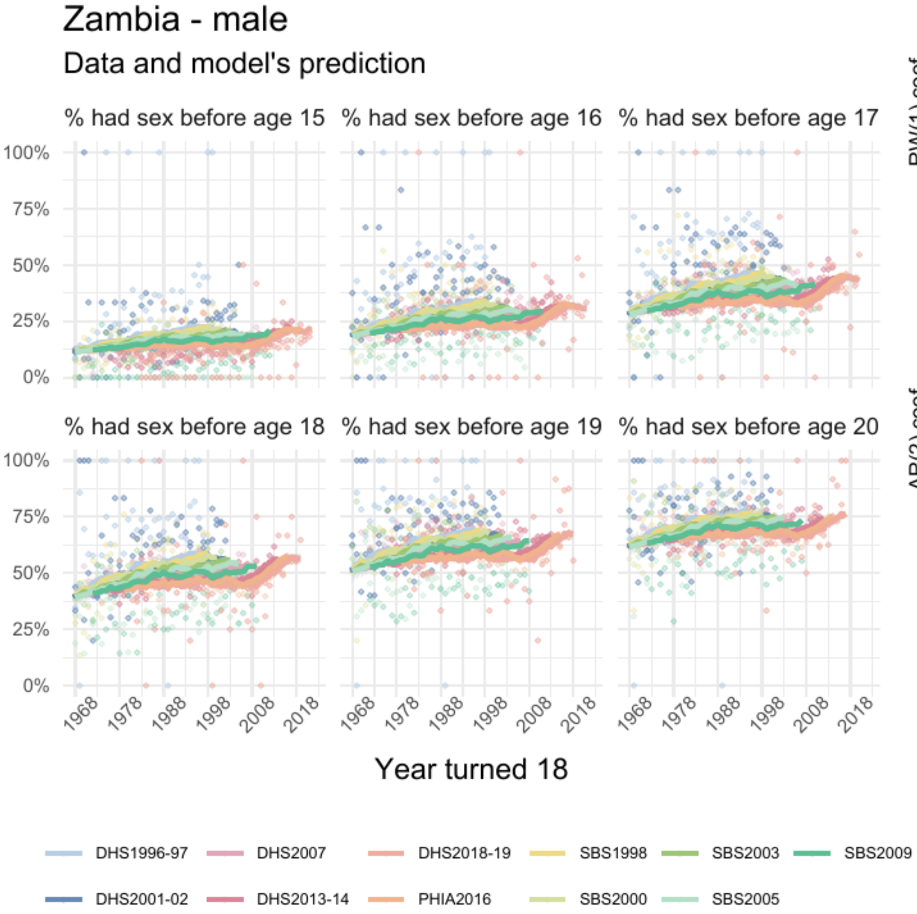


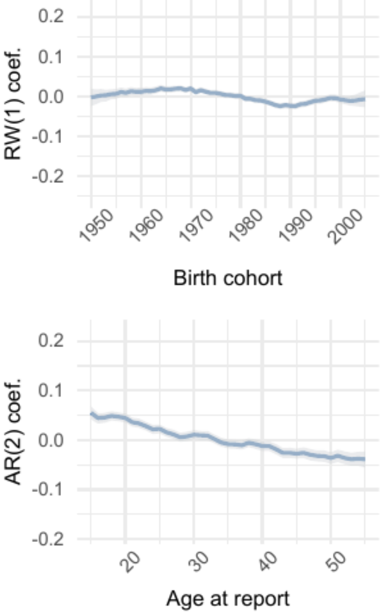


| | Parameter | Estimate |
|---|---|---|
| 1 | intercept | 0.02 [0.01 – 0.03] |
| 2 | skew | 1.09 [1.05 – 1.12] |
| 3 | shape | 8.46 [8.33 – 8.58] |

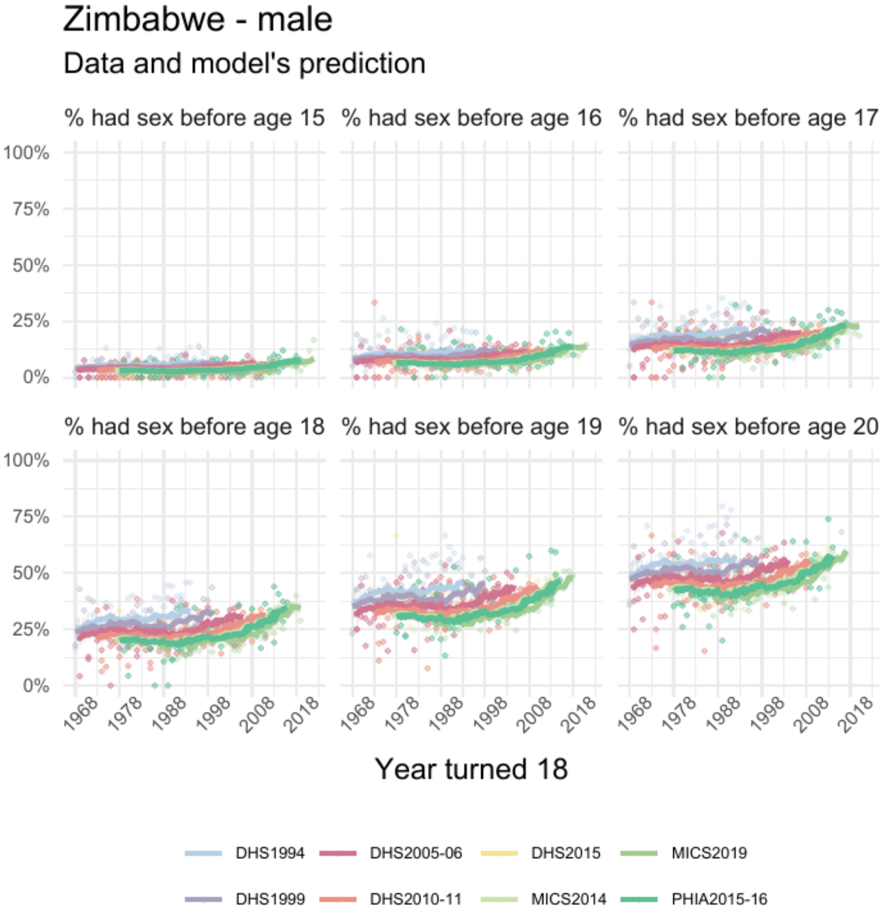


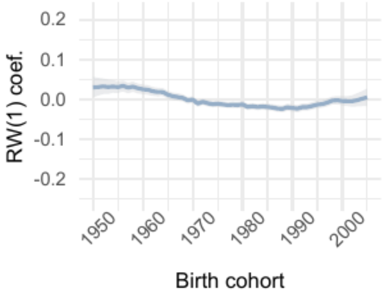


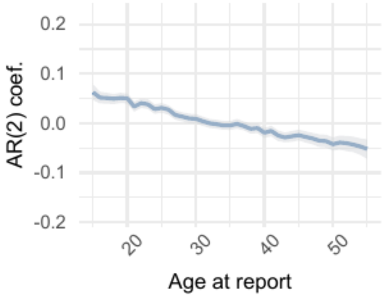


| | Parameter | Estimate |
|---|---|---|
| *1* | intercept | -0.03 [-0.04 – -0.03] |
| *2* | skew | 1.51 [1.45 – 1.57] |
| *3* | shape | 8.61 [8.48 – 8.74] |
